# Effectiveness of Covid-19 vaccines against SARS-CoV-2 Omicron variant (B.1.1.529): A systematic review with meta-analysis and meta-regression

**DOI:** 10.1101/2022.04.29.22274454

**Authors:** Nando Reza Pratama, Ifan Ali Wafa, David Setyo Budi, Henry Sutanto, Tri Pudy Asmarawati, Citrawati Dyah Kencono Wungu

**Affiliations:** Faculty of Medicine, Universitas Airlangga, Indonesia; Department of Physiology and Pharmacology, State University of New York (SUNY) Downstate Health Sciences University, New York, USA; Department of Internal Medicine, Universitas Airlangga Hospital, Universitas Airlangga, Indonesia; Institute of Tropical Disease, Universitas Airlangga, Indonesia; Department of Physiology and Medical Biochemistry, Faculty of Medicine, Universitas Airlangga, Indonesia

**Keywords:** COVID-19, SARS-CoV-2, Omicron variant, vaccine effectiveness, booster vaccination

## Abstract

**Background:** There is a need for evaluation regarding vaccine effectiveness (VE) and the urgency of booster vaccination against Covid-19 B.1.1.529 (Omicron) variant.

**Methods:** Systematic search was conducted on April 6th, 2022, on databases (PubMed, ScienceDirect, CENTRAL, Web of Science, Scopus). VE difference (VED) estimates were assessed using random-effects model and DerSimonian-Laird tau estimators. Two models result, i.e., within 3 months and within 3 months or more, are compared. VE versus time meta-regression analysis was evaluated using mixed-effects model with Restricted-Maximum Likelihood tau estimators and Hartung-Knapp adjustments.

**Findings:** Ad26.COV2.S, BNT162b2, ChAdOx1 nCov-19, and mRNA-1273 vaccines were included in the analyses. Compared to full dose, booster dose of overall vaccines provided better protection against any (VED=22% (95%CI 15%-29%), p<0.001), severe (VED=20% (95%CI 8%-32%), p=0.001) and symptomatic (VED=22% (95%CI 11%-34%), p<0.001) Omicron infections within 3 months, as well as within 3 months or more (VED=30% (95%CI 24%-37%), p<0.001 for any, VED=18% (95%CI 13%-23%), p<0.001 for severe and VED=37% (95%CI 29%-46%), p<0.001 for symptomatic infections). The meta-regression analysis of overall vaccines revealed that the full dose VE against any and symptomatic Omicron infections were significantly reduced each month by 3.0% (95%CI 0.9%-4.8%, p=0.004) and 5.2% (95%CI 3.3%-7.1%, p=0.006), respectively; whereas booster dose effectiveness against severe and symptomatic Omicron infections were decreased by 3.7% (95%CI 5.1%-12.6%, p=0.030) and 3.9% (95%CI 1.2%-6.5%, p=0.006), respectively.

**Interpretation:** Compared to full dose only, a booster dose addition provides better protection against B.1.1.529 infection. Although the VE estimates of Ad26.COV2.S, BNT162b2, ChAdOx1 nCov-19, and mRNA-1273 vaccines against B.1.1.529 infection after both full and booster doses are generally moderate, and the booster dose provides excellent protection against severe infection, it is important to note that the VE estimates decline over time, suggesting the need for a regular Covid-19 booster injection after certain period of time to maintain VE.

## INTRODUCTION

As of April 2022, there were at least two circulating SARS-CoV-2 variants of concern: the B.1.617.2 (Delta) and B.1.1.529 (Omicron) variants.^1^ In November 2021, the Omicron variant was first identified in South Africa and was immediately declared a variant of concern by the World Health Organization. Alongside the massive rise in the confirmed cases of SARS-CoV-2 infection in South Africa, the Omicron variant started to spread across the globe in no time. The identification of several concerning mutations in the SARS-CoV-2 Omicron variant and evidences of an enhanced immune escape ability contributed to the rapid spread of the Omicron variant worldwide.^2^ Compared to the ancestral variants (Wuhan-Hu-1 or Wuhan-1), the Omicron variant contains more mutations (i.e., 60 mutations), 32 of which occur in the spike gene which encodes the primary antigen target for a wide variety of Covid-19 vaccines.^3^ These mutations have been linked to increased transmissibility, a high rate of immune evasion following natural infection and vaccination, and the impairment of the efficacy of SARS-CoV-2 vaccines.^4^

Vaccine effectiveness (VE) is a measure of how well vaccines protect people from infections in the real world setting.^5^ It played a critical role in restricting the spreading of SARS-CoV-2 infections in the current Covid-19 pandemic.^6^ Earlier in the Covid-19 pandemic, studies predicted a 60-90% herd immunity threshold to limit the disease spreading, which could be achieved through several measures, including a mass vaccination campaign.^7,8^ Indeed, several Covid-19 vaccines have been shown to be promising by numerous large randomized-controlled trials (RCTs).^9–13^ Since then, many countries have extensively implemented Covid-19 vaccination programs. However, prior laboratory and clinical studies have indicated a reduction in VE against the Omicron variant as compared to the earlier variants^14–16^, potentially affecting the current Covid-19 vaccination strategy. Therefore, with the surge of new SARS-CoV-2 variants, booster vaccine doses were administered to confer stronger immunity, which hopefully could increase VE.^17,18^ However, due to global disparity in the availability, distribution of COVID-19 vaccines and the vaccination rates, developing countries were pushed to expedite booster vaccination with (limited) available resources to foster their booster vaccination rates.^19,20^

Since an equal distribution of booster vaccines remains a challenge and Omicron’s spike antigen landscape is heavily altered, there is a need for explorations regarding the effectivity of currently available vaccines and the urgency of booster vaccination against SARS-CoV-2 Omicron variant. Here, we performed systematic review and meta-analysis to unravel the effectiveness of full and booster vaccinations against the SARS-CoV-2 Omicron variant.

## METHODS

This systematic review conformed with the guidelines of Preferred Reporting Items for Systematic Review and Meta-Analysis (PRISMA) 2020^21^ and has been registered in the PROSPERO database (CRD42022302267).

### Eligibility Criteria

This review included any study designs, including RCT, cohort, case-control, and cross-sectional studies. Studies were selected according to the following criteria: (1) administration of Covid-19 vaccine during the Omicron variant’s wave as the study of interest; (2) eligible studies reporting at least one of our outcomes of interest; and (3) English language. Our outcomes included VE difference (VED) between the booster and full dose, the correlation of booster dose VE with time, and the correlation of full dose VE with time. We excluded review articles, nonhuman studies, irrelevant articles, and duplications.

### Search Strategy and Selection of Studies

Two authors (I.A.W and D.S.B) conducted a keyword search for articles published in databases (PubMed, ScienceDirect, Cochrane Central Register of Controlled Trials [CENTRAL], Web of Science, and Scopus) up to April 6^th^, 2022. Extended manual search (e.g., in medRxiv, bioRxiv) and bibliographical search were also conducted to obtain additional potential articles. The following keywords were used: “((SARS-CoV-2) OR (COVID-19)) AND ((Omicron) OR (B.1.1.529)) AND ((Vaccine) OR (Vaccination)) AND ((Vaccine efficacy) OR (Vaccine effectiveness))” Detailed search strategies are available in ***Supplementary Materials***. We exported all studies retrieved from the electronic search into the Mendeley reference manager for duplication removal and independent screening. Any disagreements between these two authors were resolved by discussion with all authors until consensus was reached. The number of excluded studies were specified in the PRISMA flow diagram alongside their reasons for exclusion **(Figure 1)**.

**Figure 1.**
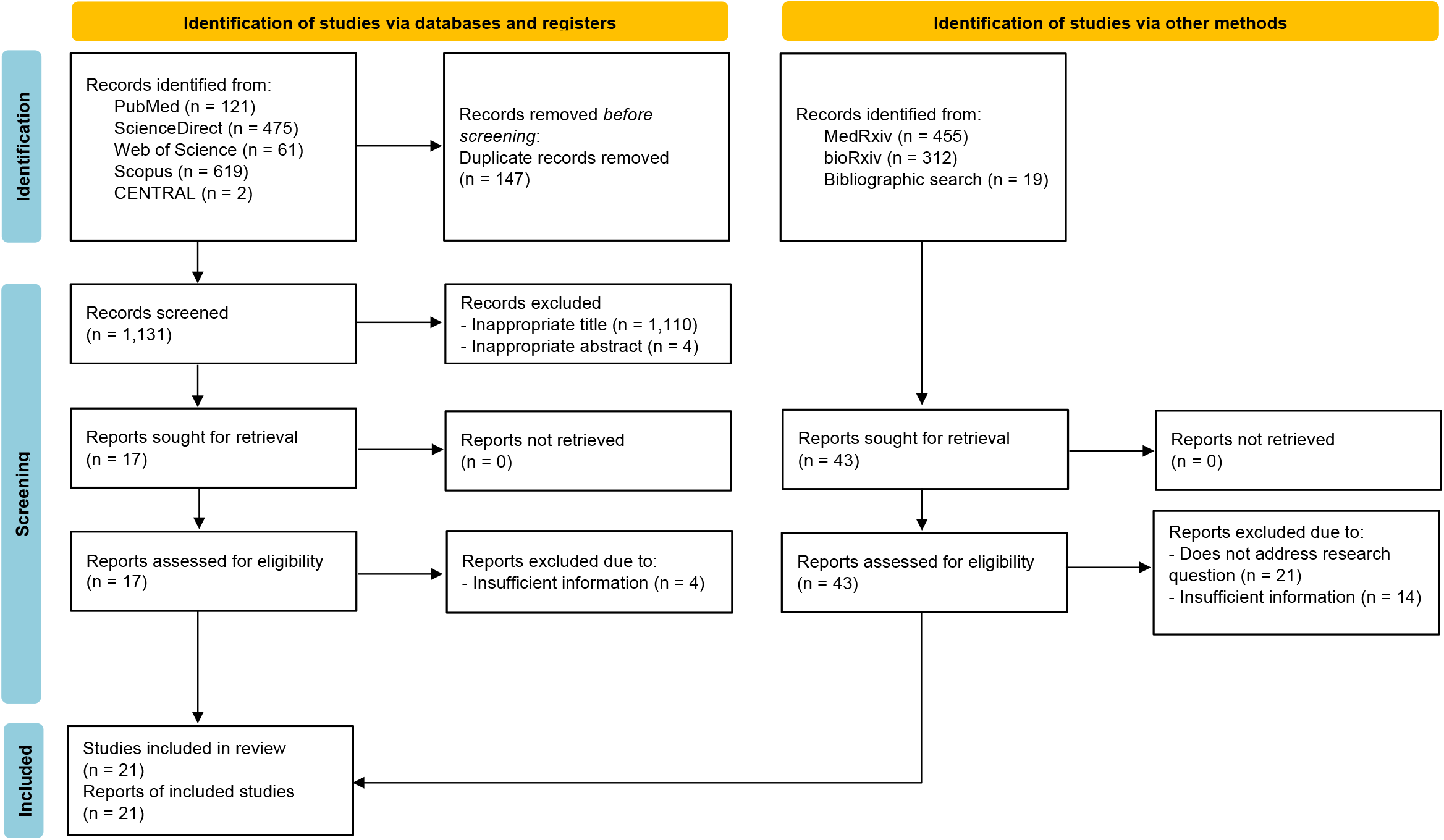
Preferred Reporting Items for Systematic Review and Meta-Analysis (PRISMA) flow diagram of study selection process^21^

### Data Extraction

Two review authors (N.R.P and D.S.B) independently extracted relevant data from each selected study using a structured and standardized form. For each included study, the following relevant data were collected: first authors’ names and publication year, study design, country of origin, sample size, patient age, Omicron strain confirmation method, follow-up duration, dose, types and administration interval of Covid-19 vaccines, endpoints, and VE.

### Quality Assessment

The methodological quality of each study was assessed independently by two authors (I.A.W and D.S.B) using the original Newcastle-Ottawa Scale (NOS) for case-control and cohort studies.^22^ The tool evaluates the quality of observational studies from the following 3 domains: (1) sample selection; (2) study comparability; and (3) study outcome. The NOS contains 8 items with scores ranging from 0 to 9. The total score of 0–3, 4–6, and 7–9 indicated low-, moderate-, and good-quality studies, respectively. Any discrepancies were resolved by discussion until consensus was reached.

### Statistical Analysis

Primary analyses were carried out using R version 4.0.5 with meta and dmetar package. VE was defined as (1–OR)x100%, (1–RR)x100% or (1–HR)x100%. VED was defined as the difference of VE between the booster and full dose vaccines, i.e., VE of booster dose – VE of full dose. We used the *I*^*2*^ test to quantify the heterogeneity between studies, with values *I*^*2*^>50% representing moderate-to-high heterogeneity. Random effects were used with the inverse variance method for pooling the results and DerSimonian-Laird for estimating τ^2^. Egger’s test was performed for the evaluation whether publication bias analyses were indicated. All statistical analyses with a p-value<0.05 was considered statistically significant. Leave-one-out sensitivity analysis was conducted to find the source of statistical heterogeneity and demonstrate how each study influenced the overall result. Meta-regression analysis was also performed using inverse-variance and Restricted-Maximum Likelihood with Hartung-Knapp adjustment. VE reduction per month was approximated by multiplying the VE reduction per day—the slope of VE versus (vs) time—by 30.

## RESULTS

### Study Selection and Quality Assessment

From databases and manual search, 1,278 and 786 records were retrieved, respectively. A total of 147 duplicates were subsequently removed. Following the screening of titles and abstracts, 60 potential articles were selected for review. After a full-text review, 21 observational studies, consisting of 5 cohorts and 16 test-negative case-control studies, were included in the systematic review, meta-analysis, and meta-regression. The overall screening process of this systematic review and meta-analysis is summarized in the PRISMA flow diagram (**Figure 1**). The quality assessment of each study using the NOS critical appraisal checklist is listed in **Table S2 and S3**. All included studies were considered good-quality studies according to the quality assessment.

### Study Characteristics

We included 21 studies, consisting of 5 cohorts and 16 test-negative case-control studies with a total of 20,691,164 participants. Four types of Covid-19 vaccine (i.e., Ad26.COV2.S, BNT162b2, ChAdOx1 nCov-19, and mRNA-1273) were included in the VE analyses of the full and booster doses. Full dose vaccination represents 2 doses of BNT162b2, ChAdOx1 nCov-19 and mRNA-1273 vaccines, or 1 dose of Ad26.COV2.S vaccines, whereas booster dose vaccination was defined as the administration of an extra dose of Covid-19 vaccine on top of the full dose vaccination. Two studies^31,37^ did not specify the type of vaccines being used; therefore, in this study, we defined it as a mixture. However, it does not necessarily mean that the vaccines are heterologous or mix-and-matched. Four studies ^29,32,34,35^ used mRNA vaccines but did not further specify the manufacturers. In this case, we defined them as a mixture of mRNA vaccines. Two studies^38,43^ evaluated the effectiveness of Covid-19 vaccines only among children and adolescents, while the rest of the studies included adults as their participants. Geographically, the included studies were originated from 8 countries/locations: 12 studies were conducted in the United States, 4 studies in Europe, 2 studies in South Africa, 2 studies in Qatar, and 1 study in Canada. The follow-up time intervals varied among studies. Between booster and full doses, only 3 studies ^32,39,41^ had a similar median or range from the date of dose receipt to the date of endpoint events.^32,39,41^ The VE was calculated as (1-OR)x100% among all case-control studies, while VE among cohort studies was calculated as (1-OR)x100%, (1-RR)x100%, or (1-HR)x100% **(Table 1)**. The VE from each study with its 95% clinical interval (CI) for full and booster doses are summarized in **Table S4** and **Table S5**, respectively. The results of VED calculation are summarized in **Table S6**.

**Table 1.** Characteristics of the included studies

| Reference | Study Design | County of Origin | Sample Sizes | Age /years | Omicron strain Confirmation Method | Follow up duration & (median (IQR)) or Range)<sup>+</sup> /days |  | Type of Vaccines | Endpoints | Vaccine effectiveness<sup>§</sup> /100% |
| --- | --- | --- | --- | --- | --- | --- | --- | --- | --- | --- |
|  |  |  |  |  |  | Dose 2 | Dose 3 |  |  |  |
| Buchan et al., 2021<sup>23</sup> | Case-control<sup>^</sup> | Canada | 134,435 | ≥ 18 | Viral whole genome or S-gene sequencing | NR | NR | BNT162b2, mRNA-1273 | Symptomatic infection and severe infection | 1-OR |
| Gray et al., 2021<sup>24</sup> | Case-control<sup>^</sup> | South Africa | 52,468 | ≥ 18 | Omicron period | N/A | 0-13 days group<br>8 (5-11) days<br>14-27 days group<br>20 (17-24) days<br>1-2 months group<br>32 (29-34) days | Ad26.COV2 | Positive Covid-19 and severe infection | 1-OR |
| Accorsi et al., 2022<sup>25</sup> | Case-control<sup>^</sup> | United States | 70,155 | ≥ 18 | Viral ORFlab, S, and N gene sequencing | 8.0 (1.0) months | 1.0 (1.0) month | BNT162b2, mRNA-1273 | Symptomatic infection | 1-OR |
| Andrews et al., 2022<sup>26</sup> | Case-control<sup>^</sup> | United Kingdom | 2,663,549 | ≥ 18 | Viral whole genome and S-gene sequencing | NR | 39 (range, 14-118) | ChAdOx1, BNT162b2, mRNA-1273 | Symptomatic infection | 1-OR |
| Chemaitelly et al., 2022<sup>27</sup> | Case-control<sup>^</sup> | Qatar | 133,327 | No restriction | Viral whole genome sequencing | NR | NR | BNT162b2, mRNA-1273 | Symptomatic infection | 1-OR |
| Collie et al., 2022<sup>28</sup> | Case-control<sup>^</sup> | South Africa | 211,610 | ≥ 18 | Viral S-gene sequencing | NR | NR | BNT162b2 | Symptomatic infection and severe infection | 1-OR |
| Ferdinand et al., 2022<sup>29</sup> | Case-control<sup>^</sup> | United States | 93,408 | ≥ 18 | Omicron-predominance period | 214 (164-259) | 49 (30-73) | Mixture of mRNA vaccines*** | Symptomatic infection* | 1-OR |
| Klein et al., 2022<sup>30</sup> | Case-control<sup>^</sup> | United States | 39,217 | ≥ 18 | Omicron-predominance period | 5 to 11 y.:<br>14-67<br>12-15 y.:<br>NR<br>16-17 y.:<br>NR | NR | BNT162b2 | Symptomatic infection* | 1-OR |
| Lauring et al., 2022<sup>31</sup> | Case-control<sup>^</sup> | United States | 11,690 | $\geq 18$ | Viral whole genome sequencing | NR | 69.5 (41.5-97) | Mixture<sup>**</sup> | Severe infection | 1-OR |
| Natarajan et al., 2022<sup>32</sup> | Case-control<sup>^</sup> | United States | 80,287 | $\geq 18$ | Omicron-predominance period | Ad26.COV 2.S: 52 (33-71)<br>Mixture: 48 (32-71) | Median (IQR) 59 (38-79) | Mixture of mRNA vaccines<sup>***</sup> | Symptomatic infection* and Hospitalization | 1-OR |
| Tartof et al., 2022<sup>33</sup> | Case-control<sup>^</sup> | United States | 14,137 | $\geq 18$ | Viral whole genome and S-gene sequencing | NR | NR | BNT162b2 | Symptomatic infection and severe infection | 1-OR |
| Tenforde et al., 2022<sup>34</sup> | Case-control<sup>^</sup> | United States | 7544 | $\geq 18$ | Omicron-predominance period | 256 | 60 | Mixture of mRNA | Severe infection<sup>#</sup> | 1-OR |
| Thompson et al., 2022<sup>35</sup> | Case-control<sup>^</sup> | United States | 222,772 | $\geq 18$ | Omicron-predominant period | < 180 days group: 137<br>$\geq 180$ days group: 223 | Median interval: 41-44 | Mixture of mRNA vaccines<sup>***</sup> | Positive Covid-19 and severe infection | 1-OR |
| Tseng et al., 2022<sup>36</sup> | Case-control<sup>^</sup> | United States | 136,345 | $\geq 18$ | Viral whole genome and S-gene sequencing | 14-365 days | NR | mRNA-1273 | Positive Covid-19 and severe infection | 1-OR |
| Young-Xu et al., 2022<sup>37</sup> | Case-control<sup>^</sup> | United States | 6,9215 | $\geq 18$ | Omicron-predominant period | NR | NR | Mixture<sup>**</sup> | Positive Covid-19 | 1-OR |
| Zambrano et al., 2022<sup>38</sup> | Case-control<sup>^</sup> | United States | 283 | 12-18 years | Viral genome sequencing | MIS-C: 63 (48-89) | N/A | BNT162b2 | Severe infection<sup>&</sup> | 1-OR |
| Abu-Raddad et al., 2022<sup>39</sup> | Retrospective Cohort | Qatar | 2,239,193 | No restriction | Viral genome sequencing | 21 (11-38) | 22 (12-38) | BNT162b2, mRNA-1273 | Symptomatic infection and severe infection | 1-HR |
| Hansen et al., 2021<sup>40</sup> | Retrospective Cohort | Denmark | 5,767 | $\geq 12$ and $\geq 60$ | Sequencing of viral whole genome or a novel variant specific targeting the 452L mutation | 1-150 | 1-30 | BNT162b2, mRNA-1273 | Positive Covid-19 | 1-HR |
| Monge et al., 2022<sup>41</sup> | Retrospective Cohort | Spain | 6,222,318 | $\geq 40$ | Omicron-predominant period | 0-34 | 0-34 | ChAdOx1-S, Ad26.COV2.S, mRNA-1273, BNT162b2 | Positive Covid-19 | 1-RR |
| Šmíd et al., 2022<sup>42</sup> | Retrospective Cohort | Czech Republic | 8,282,080 | No restriction | Viral genome sequencing | NR | NR | BNT162b2, mRNA-1273 | Symptomatic infection and Hospitalization | 1-HR |
| Fowlkes et al., 2022<sup>43</sup> | Prospective cohort | United States | 1,364 | 5 to 18 | Viral genome sequencing | 5 to 11 y.: 14-82<br>12 to 15 y.: NR | N/A | BNT162b2 | Any infection | 1-HR |

Data obtained from the emergency department and urgent care.
†Data from IMV or hospital-related death.
‡Multisystem Inflammatory Syndrome in children (MIS-C).
†The number shows a median (IQR) or range between dose receipt date and endpoint date. The purpose is to depict how data distribution between booster and full dose differ.

### Outcomes Measure

We evaluated three outcomes, i.e., VED between the booster and full dose, correlation of booster dose VE with time, and correlation of full dose VE with time. We further evaluated each of these outcomes for three different endpoints: any, symptomatic and severe Omicron infections. The ‘any infection’ outcome included positively-tested Covid-19, symptomatic Covid-19, and severe Covid-19. Meanwhile, the ‘symptomatic infection’ endpoint comprised any individuals who had been tested positive and showed Covid-19 symptoms. Individuals who require hospital visits without hospitalization were also included. Those who were hospitalized due to Covid-19, regardless of the received treatment, were deemed as having a severe infection. For VED between the full and booster doses, we analyzed the results using 2 models: the ‘within 3 months’ and the ‘within 3 months or more’ models. The ‘within 3 months’ model included data in the first 3 months reported by each study and the ‘within 3 months or more’ model included data in the first 3 months or more reported by each study.

### VED Estimates Between Booster and Full Dose

#### Overall analysis

Results from two meta-analysis models, i.e. ‘within 3 months’ (**Figure 2A**) and ‘within 3 months or more’ (**Figure 2B**) models, were compared. In the ‘within 3 months’ model, there were 22 separate analysis data involving BNT162b2 (k = 8), ChAdOx1 nCov-19 (k = 1), Mixture (k = 2), Mixture of mRNA vaccines (k = 7), and mRNA-1273 (k = 4),. Meanwhile, in the ‘within 3 months or more’ model, we obtained 28 separate analysis data that involved BNT162b2 (k = 9), ChAdOx1 nCov-19 (k = 1), Mixture (k = 2), Mixture of mRNA vaccines (k = 10), and mRNA-1273 (k = 6). As a comparison between these two models, pooled results of the ‘within 3 months or more’ model generally had a higher VED, but for severe infection, the pooled results yielded somewhat similar VED.

**Figure 2.**
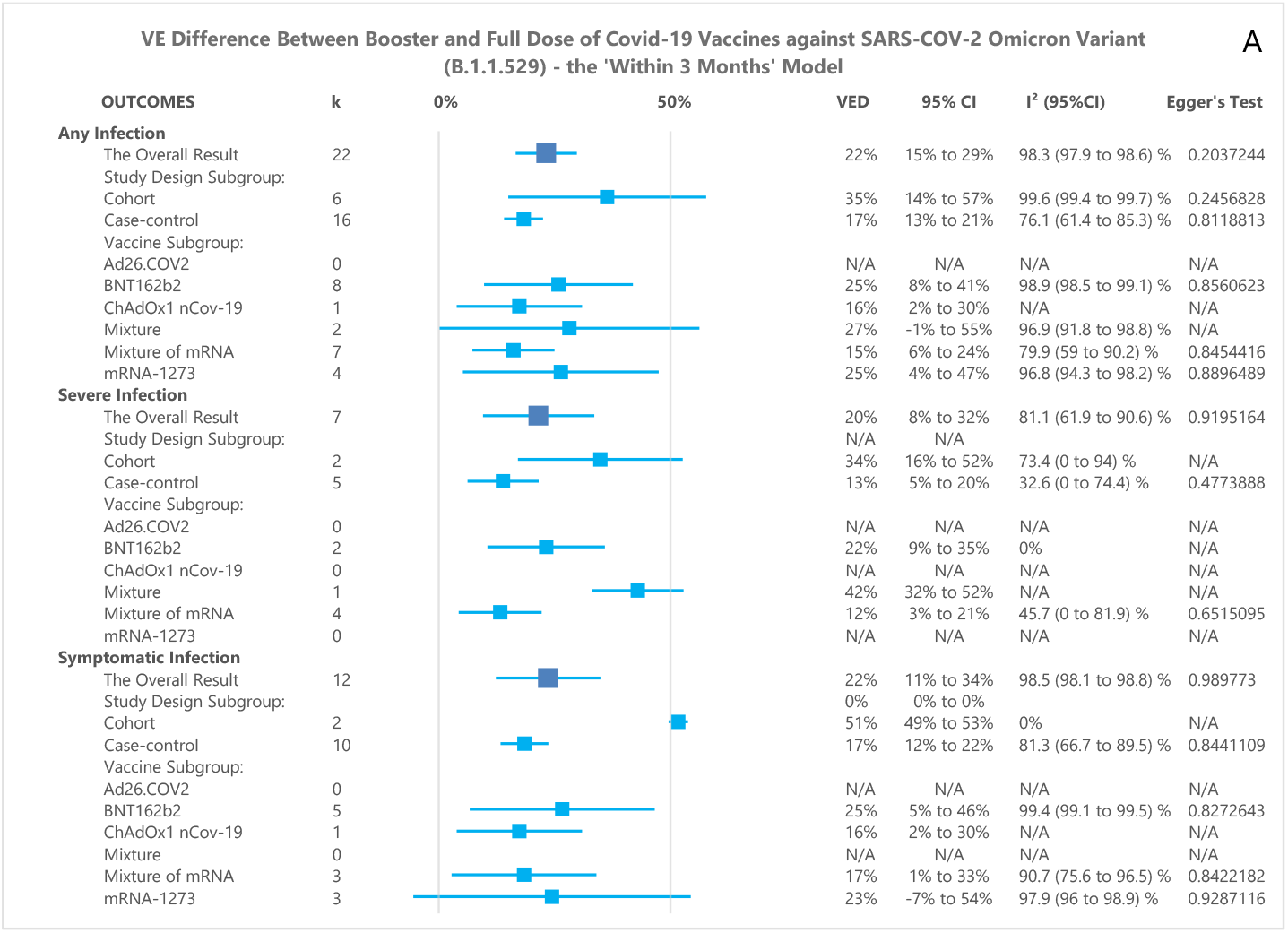
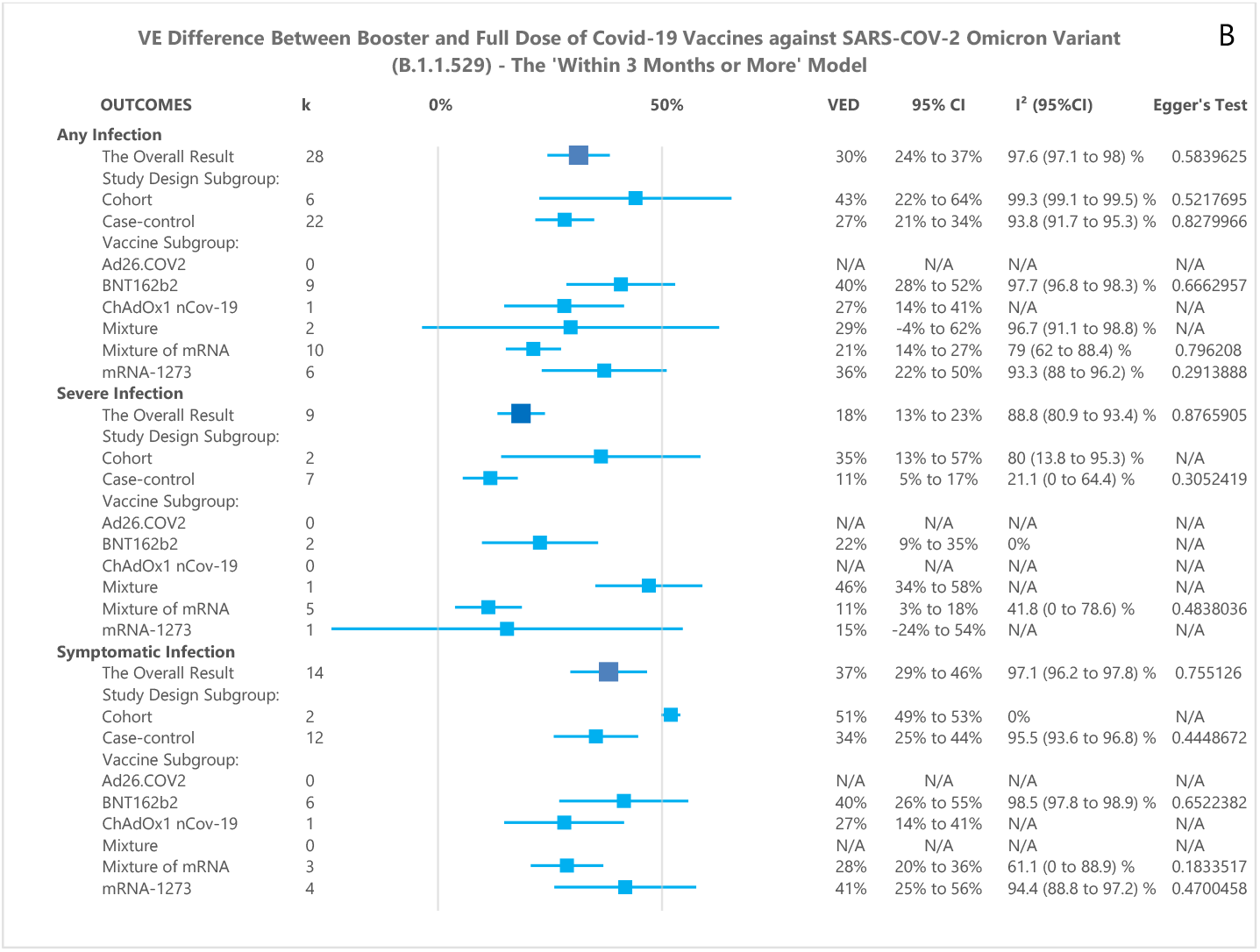
Forest Plot summary representing VED between the booster and a full dose of Covid-19 vaccine against SARS-CoV-2 infections. Panel A and B show subgroup summary of VED ‘within 3 months’ and ‘within 3 months or more’ models, respectively.

In the ‘within 3 months’ model, the pooled results for preventing any, severe and symptomatic infections showed that booster dose had a significantly higher VE than that of the full dose (VED of 22% (95%CI 15% to 29%), 20% (95%CI 8% to 32%), and 22% (95%CI 11% to 34%), respectively). In the ‘within 3 months or more’ model, the pooled results also showed a better VE on booster dose for any, severe and symptomatic infections (VED of 30% (95%CI 24% to 37%), 18% (95%CI 13% to 23%), and 37% (95%CI 29% to 46%), respectively).

The heterogeneity of overall and subgroup analyses is summarized in **Table S15** and **Table S16**. Leave-one-out sensitivity analyses performed on the overall analysis yielded similar results in terms of effect estimates or statistical heterogeneity if each study was omitted from the pooled results calculation (**Figure S1-S6**).

#### Subgroup analysis of ‘within 3 months’ model

For any infection, the cohort subgroup had a VED of 35% (95%CI 14% to 57%), but results obtained from the case-control subgroup showed a lower VED, with a VED of 17% (95%CI 13% to 21%). A further subgroup analysis with respect to vaccine type showed that BNT162b2, ChAdOx1 nCov-19, Mixture, Mixture of mRNA vaccines, and mRNA-1273 had VEDs of 25% (95%CI 8% to 41%), 16% (95%CI 2% to 30%), 27% (95%CI -1% to 55%), 15% (95%CI 6% to 24%) and 25% (95%CI 4% to 47%), respectively. These results were statistically significant except that of the Mixture subgroup.

For severe infection, cohort and case-control subgroups showed significant results (VEDs of 34% (95%CI 16% to 52%) and 13% (95%CI 5% to 20%), respectively). A further subgroup analysis showed that the VED of BNT162b2, Mixture, and Mixture of mRNA vaccines were 22% (95%CI 9% to 35%), 42% (95CI 32% to 52%), and 12% (95%CI 3% to 21%), respectively. All of these results were also statistically significant.

For symptomatic infection, cohort and case-control subgroups also showed VEDs of 51% (95%CI 49% to 53%) and 17% (95%CI 12% to 22%), respectively. In the BNT162b2, ChAdOx1 nCov-19, and Mixture of mRNA vaccines subgroups, the booster dose had a better VE than that of the full dose (VED of 25% (95%CI 5% to 46%, 16% (95%CI 2% to 30%), and 17% (95%CI 1% to 33%), respectively). These results were statistically significant, except for the mRNA-1273 vaccine subgroup, the result did not reach statistical significance (VED of 23% (95%CI -7% to 54%)).

#### Subgroup analysis of ‘within 3 months or more’ model

For any infection, the cohort subgroup had a VED of 43% (95%CI 22% to 64%), but results obtained from the case-control subgroup showed a lower VED, with a VED of 27% (95%CI 21% to 34%). A further subgroup analysis with respect to vaccine type showed that, BNT162b2, ChAdOx1 nCov-19, mixture of mRNA vaccines, and mRNA-1273 had VEDs of 40% (95%CI 28% to 52%), 27% (95%CI 14% to 41%), 21% (95%CI 14% to 27%), and 36% (95%CI 22% to 50%), respectively. All these results were statistically significant, except that of Mixture subgroup with the VED of 29% (95%CI -4% to 62%).

For severe infection, cohort and case-control subgroups showed significant results (VEDs of 35% (95%CI 13% to 57%) and 11% (95%CI 5% to 17%), respectively). A further subgroup analysis showed that the VED of BNT162b2 vaccine was 22% (95%CI 9% to 35%), while the VEDs of Mixture and Mixture of mRNA vaccines were 46% (95%CI 34% to 58%), 11% (95%CI 3% to 18%), respectively. All these results were statistically significant, except that of mRNA-1273 subgroup with the VED of 15% (95%CI -24% to 54%).

For symptomatic infection, cohort and case-control subgroups also showed significant results with VEDs of 51% (95%CI 49% to 53%) and 34% (95%CI 25% to 44%), respectively. In the BNT162b2, ChAdOx1 nCov-19, mixture of mRNA vaccines, and mRNA-1273 subgroups, the booster dose also presented a better VE than that of the full dose (VEDs of 40% (95%CI 26% to 55%), 27% (95%CI 14% to 41%), 28% (95%CI 20% to 36%), and 41% (95%CI 25% to 56%), respectively). All of these results were statistically significant.

### VE Estimates and VE reduction for Booster Dose and Full Dose

In the meta-regression analysis, the VE of full vaccination dose against any (**Figure 3A)** and symptomatic (**Figure 3C**) infections were significantly correlated with time (p = 0.0036, and p < 0.001, respectively), whereas the VE of full vaccination dose against severe infection was not (0.705). The VE of booster vaccination dose against symptomatic (**Figure 3E**) and severe (**Figure 3F**) infections were significantly correlated with time (p = 0.030 and p = 0.006, respectively), while the VE of booster vaccination dose against any infection was not (p = 0.1051). The correlation of VE (%) with time (day) was 50.12 - 0.1 per day (**Figure. 3A**); 68.28 - 0.018 per day (**Figure. 3B**); 58.21 - 0.173 per day (**Figure. 3C**); 66.04 - 0.077 per day (**Figure. 3D**); 93.96 - 0.054 per day (**Figure. 3E**); 65.03 - 0.129 per day (**Figure. 3F**).

**Figure 3.**
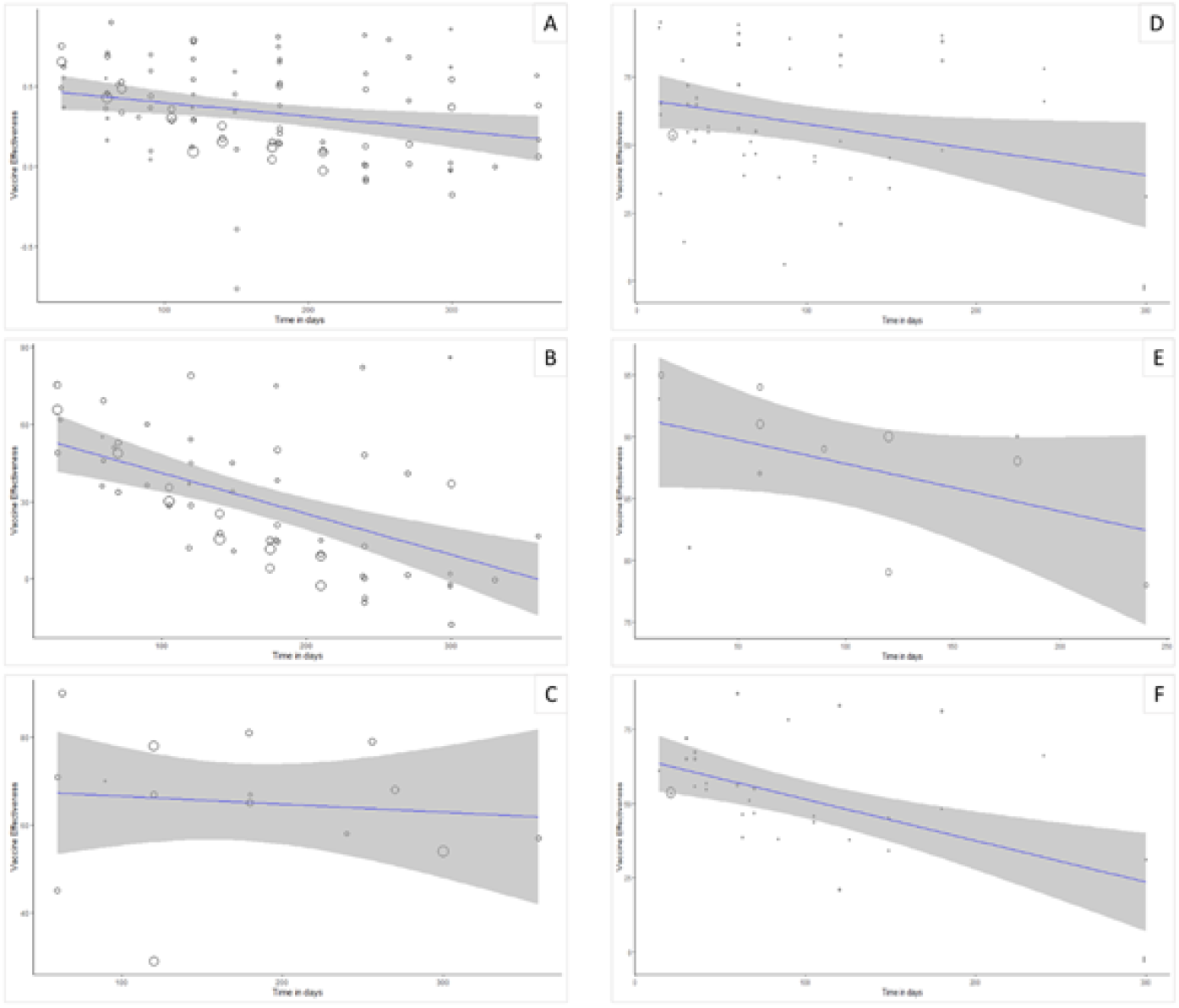
Meta-regression plot for VE vs Time in days. VE on panel A, B, and C represent VE of full dose against any infection, severe infection, and symptomatic infection, respectively. VE on panel D, E, and F represent VE of booster dose against any infection, severe infection, and symptomatic infection, respectively. VE estimate (%) was 50.12 (95%CI 37.78 to 62.46), p<0.001 (A); 68.28 (95%CI 48.71 to 87.85), p<0.001 (B); 58.21 (95%CI 46.26 to 70.15), p<0.001 (C); 66.04 (95%CI 55.32 to 76.75), p<0.001 (D); 93.96 (95%CI 88.3 to 99.63), p<0.001 (E); 65.03 (95%CI 54.59 to 75.48), p<0.001 (F).

For the full dose vaccination, our meta-regression model estimated that the VE of full dose against any and symptomatic infections were decreased in each month approximately by 3.0% (95%CI 0.9% to 4.8%) and 5.2% (95%CI 3.3% to 7.1%), respectively. Meanwhile, the VE of booster dose against severe and symptomatic infections were decreased in each month by 3.7% (95%CI 5.1% to 12.6%) and 3.9% (95%CI 1.2% to 6.5%), respectively. The detailed results displaying the correlation of VE with time for each vaccine were presented in **Table 2**.

**Table 2.**
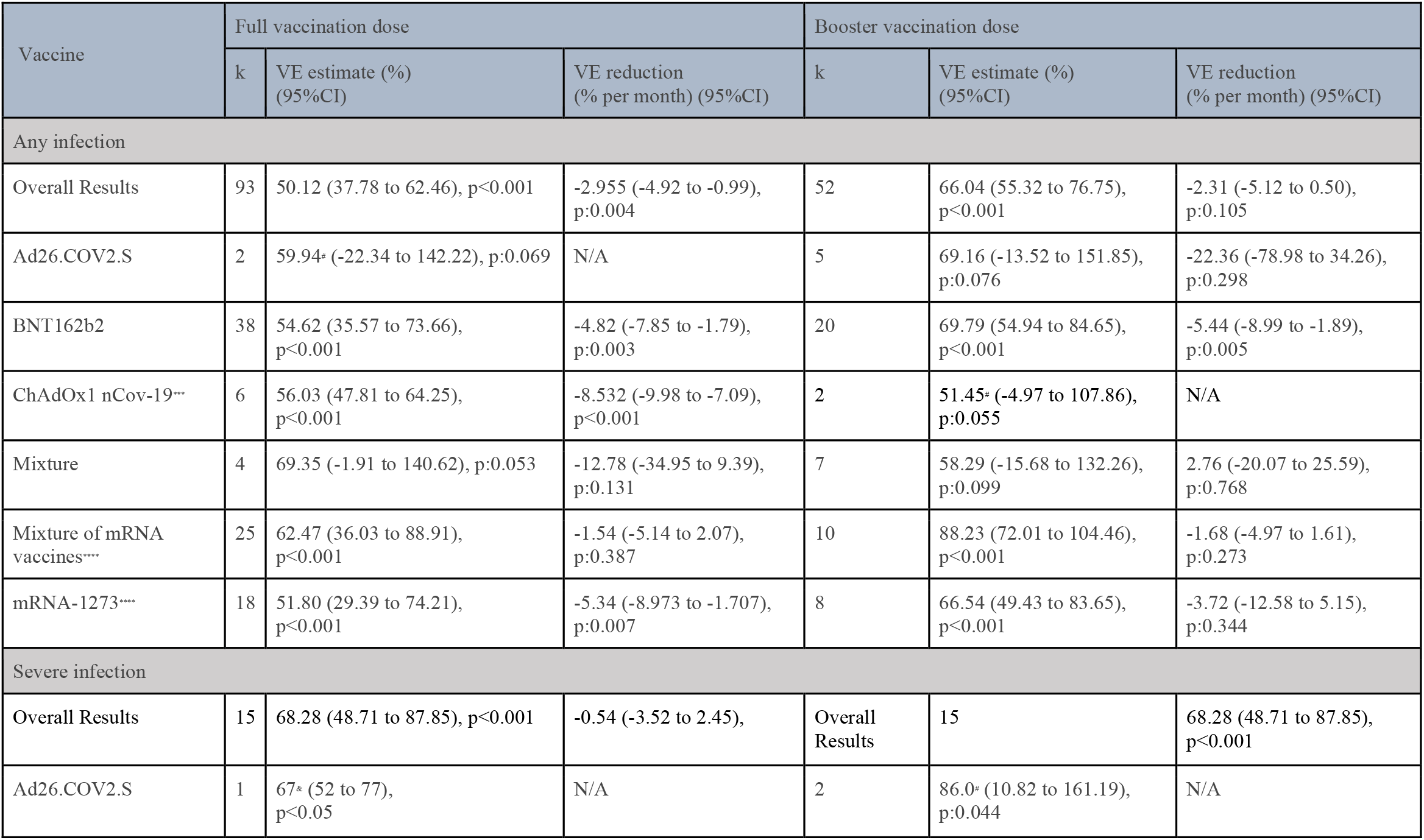
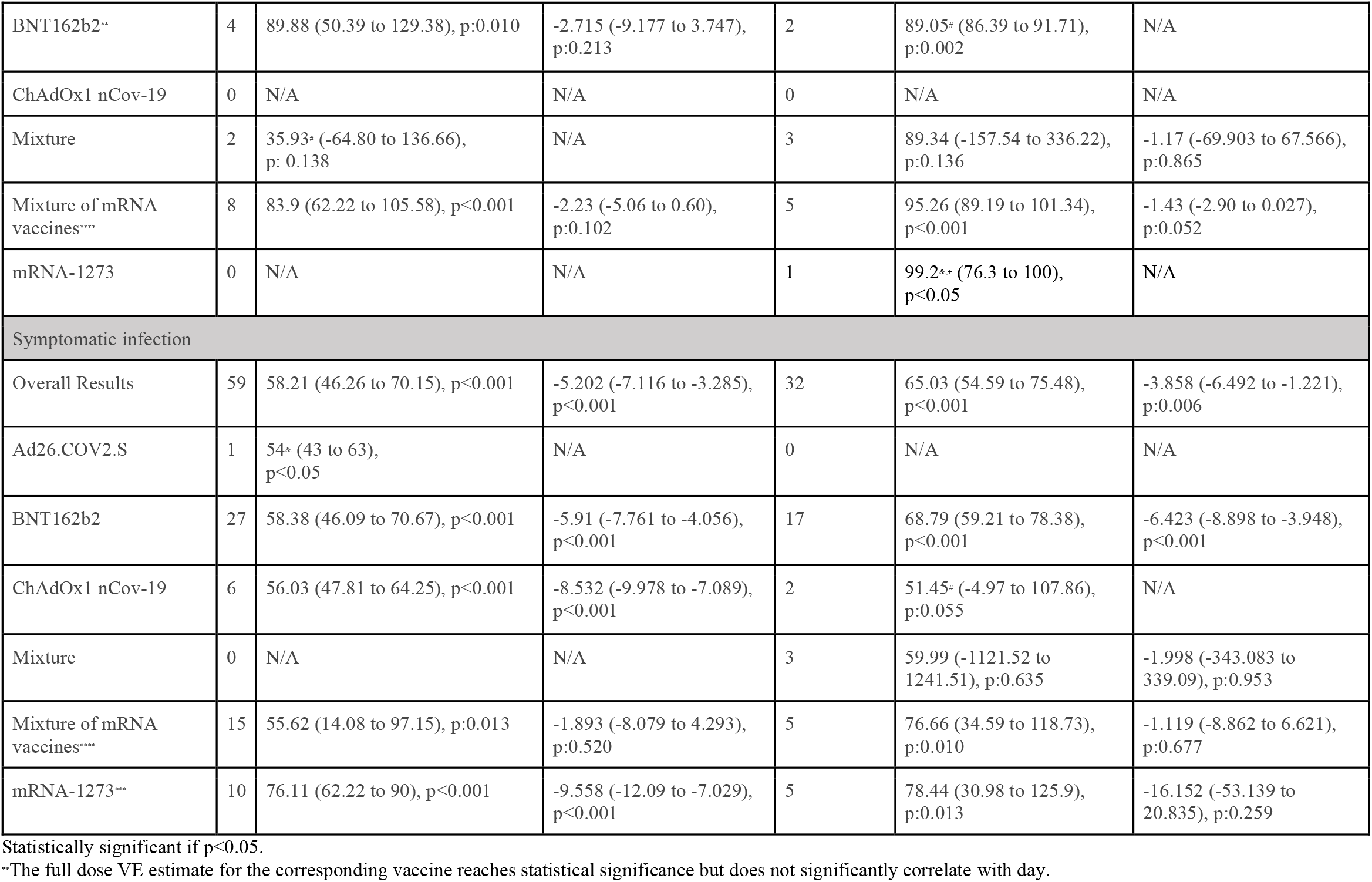
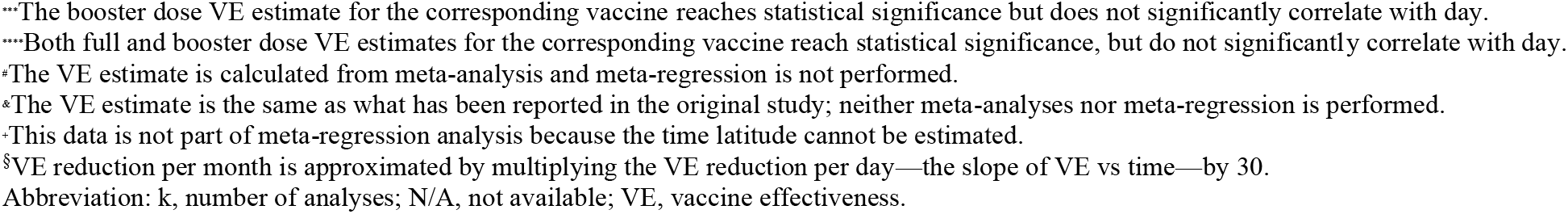
VE estimates and VE reduction for booster dose and full dose of each vaccine.

## DISCUSSION

This study aimed to evaluate the full dose and booster VE, and their correlation with time to evaluate waning immunity. We used two models for VED evaluation based on time period, i.e., ‘within 3 months’ and ‘within 3 months or more’. Our models demonstrated that there was a significant VED between the booster and full dose in terms of preventing any, symptomatic and severe infections of the SARS-CoV-2 Omicron variant. Although these results had a generally high heterogeneity, subgroup analyses showed that study design and types of vaccine did not seem to contribute to this heterogeneity. However, VED at a more prolonged interval model, i.e. ‘within 3 months or more’, was more elevated. Most analyses at a longer interval contained more full dose vaccine data since the follow-up interval for the full vaccination dose was considerably longer than that of the booster dose.

On the other hand, our meta-regression analysis showed that the full dose VE against any and severe infections were estimated to be reduced by 3.0% and 5.2% each month, respectively. Meanwhile, the booster dose VE against severe and symptomatic infections were estimated to be reduced by 3.7% and 3.9% each month, respectively. Additionally, VE estimates of booster dose were generally higher than that of full dose, in line with VED meta-analysis results. VE estimates of booster doses were generally at more than 60% for all endpoints. Booster doses of mRNA vaccines showed excellent protection against severe infection with a VE of 95.26%, compared to full dose with a VE of 83.9%. This result was in agreement with the prediction by Khoury *et al*.^44^ In that study, it was predicted that a booster dose could raise VE from 81.1%, by full dose, to 98.2% for mRNA vaccines. Also, in that study, BNT162b2 and mRNA-1273 were predicted to have more than 80% VE against severe infection, and VE against severe infection was generally higher than that of symptomatic infection.^44^

Vaccination or natural infection induces some immune cell subsets to turn into memory cells through clonal expansions.^45^ These previously primed cells could deliver a more robust immune response in the secondary response, which is protective against severe disease. Meanwhile, neutralizing antibodies can provide sterilizing immunity to prevent infection.^45–47^ Neutralizing antibodies produced by plasma B cells decay over time, but long-lived plasma B memory cells continuously secrete neutralizing antibodies even after the infection ends, maintaining their level.^48^ Moreover, a robust immune response and multiple infections or vaccination can elicit strong immunoglobulin G (IgG)-binding affinity as a result of an affinity maturation process.^49,50^ Consequently, compared to full vaccination doses, an additional booster vaccination dose would elicit a stronger immune response,^51^ as evidenced by some studies that reported a higher binding affinity and titers of neutralizing antibodies among individuals who received three vaccination doses than that of two doses.^51–53^ For instance, with regard to the Omicron variant, the antibody titer induced by a booster dose of BNT162b2 at 1 month were 23-fold higher than that of full dose recipients.^54^

Some studies showed that neutralizing antibody titers in Covid-19 were predictive of immune protection.^55,56^ Neutralization titers continuously declined and appeared to be short-lived,^57,58^ and immune escape was observed in several SARS-CoV-2 variants of concern,^59–62^ leading to reduced VE to some extent.^57^ The immune escape may cause a reduced VE among variants of concern, regardless of how long the last vaccination is given before the infection. A meta-analysis with modeling study has previously demonstrated the correlation between neutralization titers with VE.^57^

The magnitude of VE reduction depends on the initial efficacy. The VE against symptomatic infection could drop, by day 250, to 77% or 33% if initial efficacy was 95% or 70%, respectively.^63^ Thus, the efficacy of a vaccine may vary across types of vaccine, doses, and variants. The study by Khoury *et al*.^44^ estimated that neutralizing antibody levels needed to protect from severe infection were 6-fold lower than symptomatic infection, which could explain the reason why the VE against severe infection in our study remained high, despite the low VE against symptomatic infection.^63^ Since short-lived or substantial decay of neutralizing antibody titers means an increased vulnerability towards symptomatic infection, a persistent cellular immune memory enables a faster and stronger secondary immune response.^64,65^ An appropriate secondary immune response, especially T-cell response, is protective against severe infection.^66–68^

Our analyses showed a moderate VE reduction against the Omicron variant. Nonetheless, the Omicron variant did not display an increased severity despite the increased transmissibility.^69–71^ Our results showed that VE estimates against severe infection still exhibit a high efficacy for both the full and booster doses. However, we should be aware that new variants of concern may emerge anytime, and always need to be anticipated. Maintaining the Covid-19 pandemic to a low endemic level is seemingly a reasonable target before the eradication of Covid-19 could be achieved.

We acknowledge that this study has some limitations. First, most results had high heterogeneity. In the meta-analysis of VED outcome, we attempted to perform subgroup analyses based on the study design and types of the vaccine, but the heterogeneity remained high. Since there was a considerable discrepancy in the follow-up time between the booster and full doses, we suspected that the high heterogeneity was due to covariate time, as we have demonstrated in the other outcomes. Secondly, all included studies were observational studies. In observational studies, some confounding factors are difficult to measure, thereby cannot be controlled. For example, significant differences in the follow-up time would result in different exposure received between the two groups. Moreover, the fact that VE declines over time should be considered because cumulative comparison would lead to a bias. As the result, we attempted to limit the time interval in one model to only include data within three months to minimize this bias. Third, some included studies were obtained from preprint servers, which had not been preceded by a peer-review process and the presented data may differ from the final published, peer-reviewed, version.

## CONCLUSION

A lower initial VE supports the evidence that the SARS-CoV-2 Omicron variant has an increased immune escape ability, and the decline of VE over time indicates the presence of waned immunity in the SARS-CoV-2 Omicron variant infection. The VE of the booster dose was generally higher than that of the full dose. Although VE remained high in both the full and booster doses for severe infections, a booster vaccination dose is still recommended to confer utmost protection against the SARS-CoV-2 Omicron variant infection. Moreover, the emergence of other variants of concern should always be anticipated. Nevertheless, these meta-analyses and meta-regression were constructed upon observational studies, in which several confounders adjustment could be difficult to perform. Therefore, future RCTs might be able to address several limitations of this study.

## Data Availability

All data produced in the present study are available upon reasonable request to the authors

## Contributors

NRP, IAW, DSB developed the study design, data collection, data analysis, and writing (Original Draft Preparation). HS contributed to the writing (Review and Editing), manuscript validation, and supervision. TPA developed the study design, methodology, manuscript validation, and supervision. CDKW developed the study design, data analysis, methodology, manuscript validation, supervision. All authors reviewed and approved the final manuscript.

## Data Sharing

The protocol for the systematic review is available at: https://www.crd.york.ac.uk/prospero/display_record.php?ID=CRD42022302267. Detailed methods, results, and additional data are available in the manuscript and the associated appendix.

