## Supplementary Materials for "Effectiveness of Covid-19 vaccines against SARS-CoV-2 Omicron variant (B.1.1.529): A systematic review with meta-analysis and meta-regression"

### S.1. Literature Search

#### The Keywords

#1 (SARS-CoV-2) OR (COVID-19)

#2 (Omicron) OR (B.1.1.529)

#3 (Vaccine) OR (Vaccination)

#4 (Vaccine efficacy) OR (Vaccine effectiveness)

#### The Search Results

Table S1. Results of systematic searches.

| **Database** | **Keywords** | **Search Result** | **Search-time** |
|---|---|---|---|
| PubMed | #1 AND #2 AND #3 AND #4 | 121 | April 6$^{th}$, 2022 |
| ScienceDirect | #1 AND #2 AND #3 AND #4 | 475 | April 6$^{th}$, 2022 |
| CENTRAL | #1 AND #2 AND #3 AND #4 | 2 | April 6$^{th}$, 2022 |
| Web of Science | #1 AND #2 AND #3 AND #4 | 61 | April 6$^{th}$, 2022 |
| Scopus | #1 AND #2 AND #3 AND #4 | 619 | April 6$^{th}$, 2022 |
| medRxiv | #1 AND #2 AND #3 AND #4 | 455 | April 6$^{th}$, 2022 |
| bioRxiv | #1 AND #2 AND #3 AND #4 | 312 | April 6$^{th}$, 2022 |
| Bibliographic search | - | 19 | April 6$^{th}$, 2022 |

#### S.2. Assessment of Quality of Study

Table S2. The assessment of quality of study for cohort studies using Newcastle-Ottawa Scale (NOS)

| NOS of Cohort Study | | | | | |
|---|---|---|---|---|---|
| **Components** | Hansen et al. (2021) | Abu-Raddad et al. (2022) | Monge et al., (2022) | Šmíd et al. (2022) | Fowlkes et al. (2022) |
| **Selection** | | | | | |
| Representativeness of the exposed cohort | * | * | * | * | * |
| Selection of the non-exposed cohort | * | * | * | * | * |
| Ascertainment of exposure | * | * | * | * | * |
| Demonstration that outcome of interest was not present at start of the study | * | * | - | - | - |
| **Comparability** | | | | | |
| Comparability of cohorts on the basis of design or analysis | * | ** | * | * | * |
| **Exposure** | | | | | |
| Assessment of outcome | * | * | * | * | * |
| Enough follow-up time length for outcome to occur | * | * | * | * | * |
| Adequacy of follow-up of cohorts | * | * | * | * | * |
| **Study Quality** | | | | | |
| **Total Score** | 8 | 9 | 7 | 7 | 7 |
| **Interpretation** | Good | Good | Good | Good | Good |

Table S3. The assessment of quality of study for case-control studies using Newcastle-Ottawa Scale (NOS)

| NOS of Case-Control Study | | | | | | | | | | |
|---|---|---|---|---|---|---|---|---|---|---|
| | **Selection** | | | | **Comparability** | **Exposure** | | | **Study Quality** | |
| **Components** | Is the case definition adequate? | Representative-ness of the cases | Selection of controls | Definition of controls | Comparability of cases and controls on the basis of the design or analysis | Ascertainment of exposure | Same method of ascertainment for cases and controls | Non-response rate | **Total Score** | **Interpretation** |
| Buchan et al. (2021) | * | * | * | * | * | - | * | * | 7 | Good |
| Gray et al. (2021) | * | * | * | * | * | * | * | * | 8 | Good |
| Accorsi et al. (2022) | * | * | * | * | * | - | * | * | 7 | Good |
| Andrews et al. (2022) | * | * | * | * | * | * | * | * | 8 | Good |
| Chemaitelly et al. (2022) | * | * | * | * | * | * | * | * | 8 | Good |
| Collie et al. (2022) | * | * | * | * | * | * | * | * | 8 | Good |
| Ferdinand et al. (2022) | * | * | * | * | * | * | * | * | 8 | Good |
| Klein et al. (2022) | * | * | * | * | * | * | * | * | 8 | Good |
| Lauring et al. (2022) | * | * | * | * | ** | * | * | * | 9 | Good |
| Natarajan et al. (2022) | * | * | * | * | ** | * | * | * | 9 | Good |
| Tartof et al. (2022) | * | * | * | * | * | * | * | * | 8 | Good |
| Tenforde et al. (2022) | * | * | * | * | * | - | * | * | 7 | Good |

| | | | | | | | | | | |
|---|---|---|---|---|---|---|---|---|---|---|
| Thompson et al. (2022) | * | * | * | * | * | * | * | * | 8 | Good |
| Tseng et al. (2022) | * | * | * | * | ** | * | * | * | 9 | Good |
| Young-Xu et al. (2022) | * | * | * | * | * | * | * | * | 8 | Good |
| Zambrano et al. (2022) | * | * | * | * | * | * | * | * | 8 | Good |

#### S.3. Summary of outcomes for full dose

##### Table S4. Summary of outcomes for full dose

| Study | Endpoints | Vaccine | Study Design | Days Latitude | Follow-up Interval | SE | VE (95%CI) |
|---|---|---|---|---|---|---|---|
| Ferdinand et al., 2022 | Severe infection | Mixture of mRNA | Case-control | 60 | 0 to 2 months | 8.163 | 71% (51% to 83%) |
| | | Mixture of mRNA | Case-control | 180 | 2 to 3 months | 5.357 | 65% (53% to 74%) |
| | | Mixture of mRNA | Case-control | 240 | 3 to 4 months | 8.418 | 58% (38% to 71%) |
| | | Mixture of mRNA | Case-control | 300 | 5+ months | 2.806 | 54% (48% to 59%) |
| | Symptomatic Infection | Mixture of mRNA | Case-control | 60 | 0 to 2 months | 3.316 | 69% (62% to 75%) |
| | | Mixture of mRNA | Case-control | 180 | 2 to 3 months | 2.551 | 50% (45% to 55%) |
| | | Mixture of mRNA | Case-control | 240 | 3 to 4 months | 3.316 | 48% (41% to 54%) |
| | | Mixture of mRNA | Case-control | 300 | 5+ months | 1.531 | 37% (34% to 40%) |
| Klein et al., 2022 | Symptomatic infection (Age 16-17 years) | BNT162b2 | Case-control | 149 | 14 to 149 days | 11.480 | 34% (8% to 53%) |
| | | BNT162b2 | Case-control | 299 | 150+ days | 12.245 | -3% (-30% to 18%) |
| | | BNT162b2 | Case-control | 149 | 14 to 149 days | 6.888 | 45% (30% to 57%) |
| | | BNT162b2 | Case-control | 299 | 150+ days | 10.714 | -2% (-25% to 17%) |
| | | BNT162b2 | Case-control | 67 | 14 to 67 days | 8.929 | 51% (30% to 65%) |
| Natarajan et al., 2022 | Severe infection | AD26.COV2S | Case-control | 120 | 7 to 120 days | 6.378 | 67% (52% to 77%) |
| | | Mixture of mRNA | Case-control | 120 | 7 to 120 days | 3.571 | 78% (70% to 84%) |
| | Symptomatic Infection | AD26.COV2S | Case-control | 120 | 7 to 120 days | 5.102 | 54% (43% to 63%) |
| | | Mixture of mRNA | Case-control | 120 | 7 to 120 days | 2.041 | 79% (74% to 82%) |
| Tenforde et al., 2022 | Severe infection | Mixture of mRNA | Case-control | 256 | 256 days (median) | 5.357 | 79% (66% to 87%) |
| Zambrano et al., 2022 | Severe Infection | BNT162b2 | Case-control | 63 | 63 days (median) | 5.357 | 90% (75% to 96%) |
| Accorsi et al., 2022* | Symptomatic Infection | BNT162b2 | Case-control | 240 | 14+ days | 2.653 | 0% (-5% to 5%) |
| | | mRNA-1273 | Case-control | 240 | 14+ days | 2.679 | 13% (7% to 18%) |
| Andrews et al., 2022 | Symptomatic Infection | ChAdOx1 nCov-19 | Case-control | 28 | 2 to 4 weeks | 4.566 | 49% (39% to 57%) |

| | | | | | | | |
|---|---|---|---|---|---|---|---|
| | | ChAdOx1 nCov-19 | Case-control | 70 | 5 to 9 weeks | 4.209 | 34% (25% to 42%) |
| | | ChAdOx1 nCov-19 | Case-control | 105 | 10 to 14 weeks | 3.750 | 29% (21% to 36%) |
| | | ChAdOx1 nCov-19 | Case-control | 140 | 15 to 19 weeks | 2.168 | 18% (13% to 22%) |
| | | ChAdOx1 nCov-19 | Case-control | 175 | 20 to 24 weeks | 1.071 | 4% (2% to 6%) |
| | | ChAdOx1 nCov-19 | Case-control | 210 | 25+ weeks | 0.765 | -3% (-4% to -1%) |
| | | BNT162b2 | Case-control | 28 | 2 to 4 weeks | 0.791 | 66% (64% to 67%) |
| | | BNT162b2 | Case-control | 70 | 5 to 9 weeks | 0.791 | 49% (47% to 50%) |
| | | BNT162b2 | Case-control | 105 | 10 to 14 weeks | 0.714 | 30% (29% to 32%) |
| | | BNT162b2 | Case-control | 140 | 15 to 19 weeks | 0.612 | 15% (14% to 17%) |
| | | BNT162b2 | Case-control | 175 | 20 to 24 weeks | 0.714 | 12% (10% to 13%) |
| | | BNT162b2 | Case-control | 210 | 25+ weeks | 0.893 | 9% (7% to 11%) |
| | | mRNA-1273 | Case-control | 28 | 2 to 4 weeks | 2.015 | 75% (71% to 79%) |
| | | mRNA-1273 | Case-control | 70 | 5 to 9 weeks | 2.270 | 53% (48% to 57%) |
| | | mRNA-1273 | Case-control | 105 | 10 to 14 weeks | 1.454 | 36% (33% to 38%) |
| | | mRNA-1273 | Case-control | 140 | 15 to 19 weeks | 1.071 | 25% (23% to 27%) |
| | | mRNA-1273 | Case-control | 175 | 20 to 24 weeks | 1.684 | 15% (12% to 18%) |
| | | mRNA-1273 | Case-control | 210 | 25+ weeks | 5.306 | 15% (4% to 25%) |
| Buchan et al., 2021 | Symptomatic Infection | Mixture of mRNA | Case-control | 59 | 7 to 59 days | 5.357 | 36% (24% to 45%) |
| | | Mixture of mRNA | Case-control | 119 | 60 to 119 days | 4.592 | 12% (3% to 21%) |
| | | Mixture of mRNA | Case-control | 179 | 120 to 179 days | 3.571 | 15% (8% to 22%) |
| | | Mixture of mRNA | Case-control | 239 | 180 to 239 days | 4.592 | 1% (-8% to 10%) |
| | | Mixture of mRNA | Case-control | 299 | 240+ days | 8.673 | 2% (-17% to 17%) |
| | | Mixture of mRNA | Case-control | 59 | 7 to 59 days | 50.000 | 55% (-106% to 90%) |
| | | Mixture of mRNA | Case-control | 119 | 60 to 119 days | 37.755 | 37% (-71% to 77%) |
| | | Mixture of mRNA | Case-control | 179 | 120 to 179 days | 9.184 | 75% (51% to 87%) |
| | | Mixture of mRNA | Case-control | 239 | 180 to 239 days | 7.398 | 82% (62% to 91%) |

| | | | | | | | |
|---|---|---|---|---|---|---|---|
| | | Mixture of mRNA | Case-control | 299 | 240+ days | 28.061 | 86% (-12% to 98%) |
| Chemaitelly et al., 2022 | Symptomatic Infection | BNT162b2 | Case-control | 30 | 0 to 1 month | 5.408 | 62% (50% to 71%) |
| | | BNT162b2 | Case-control | 60 | 1 to 2 months | 5.612 | 46% (34% to 56%) |
| | | BNT162b2 | Case-control | 90 | 2 to 3 months | 5.281 | 36% (25% to 46%) |
| | | BNT162b2 | Case-control | 120 | 3 to 4 months | 5.051 | 29% (18% to 38%) |
| | | BNT162b2 | Case-control | 150 | 4 to 5 months | 6.173 | 11% (-2% to 22%) |
| | | BNT162b2 | Case-control | 180 | 5 to 6 months | 3.980 | 14% (6% to 22%) |
| | | BNT162b2 | Case-control | 210 | 6 to 7 months | 3.546 | 10% (2% to 16%) |
| | | BNT162b2 | Case-control | 240 | 7 to 8 months | 3.852 | -8% (-15% to 0%) |
| | | BNT162b2 | Case-control | 270 | 8 to 9 months | 3.801 | 2% (-6% to 9%) |
| | | BNT162b2 | Case-control | 300 | 9 to 10 months | 3.903 | -18% (-26% to -10%) |
| | | BNT162b2 | Case-control | 330 | 10 to 11 months | 4.796 | 0% (-10% to 9%) |
| | | BNT162b2 | Case-control | 360 | 11+ months | 6.378 | 17% (3% to 28%) |
| | | mRNA-1273 | Case-control | 120 | 1 to 3 months | 12.194 | 45% (16% to 64%) |
| | | mRNA-1273 | Case-control | 180 | 4 to 6 months | 3.495 | 21% (14% to 27%) |
| | | mRNA-1273 | Case-control | 240 | 7+ months | 3.444 | -9% (-16% to -3%) |
| Collie et al., 2022 | Severe Infection | BNT162b2 | Case-control | NR | 14+ days | 6.888 | 50% (35% to 62%) |
| | Symptomatic Infection | BNT162b2 | Case-control | NR | 0+ days | 4.847 | 70% (59% to 78%) |
| Lauring et al., 2022 | Severe Infection | Mixture of mRNA | Case-control | NR | 7+ days | 7.143 | 66% (49% to 77%) |
| Tartof et al., 2022 | | BNT162b2 | Case-control | 90 | 0 to 3 months | 10.969 | 70% (41% to 84%) |
| | | BNT162b2 | Case-control | 180 | 3 to 6 months | 9.184 | 67% (44% to 80%) |
| | | BNT162b2 | Case-control | 270 | 6+ months | 5.102 | 68% (56% to 76%) |
| | Symptomatic Infection | BNT162b2 | Case-control | 90 | 0 to 3 months | 7.398 | 60% (43% to 72%) |
| | | BNT162b2 | Case-control | 180 | 3 to 6 months | 7.653 | 38% (21% to 51%) |
| | | BNT162b2 | Case-control | 270 | 6+ months | 4.592 | 41% (32% to 50%) |
| Thompson et al., 2022 | Positive Covid-19 | Mixture of mRNA | Case-control | 180 | 0 to 180 days | 3.061 | 52% (46% to 58%) |
| | | Mixture of mRNA | Case-control | 360 | 180+ days | 2.806 | 38% (32% to 43%) |
| | | Mixture of mRNA | Case-control | 179 | 0 to 179 days | 6.378 | 81% (65% to 90%) |

| | | | | | | | |
|---|---|---|---|---|---|---|---|
| | Severe Infection | Mixture of mRNA | Case-control | 359 | 180+ days | 7.908 | 57% (39% to 70%) |
| Tseng et al., 2022 | Positive Covid-19 | mRNA-1273 | Case-control | 90 | 14 to 90 days | 4.209 | 44% (35% to 52%) |
| | | mRNA-1273 | Case-control | 180 | 91 to 180 days | 3.469 | 24% (16% to 30%) |
| | | mRNA-1273 | Case-control | 270 | 181 to 270 days | 1.811 | 14% (10% to 17%) |
| | | mRNA-1273 | Case-control | 360 | 270+ days | 2.704 | 6% (0% to 11%) |
| | Severe Infection | mRNA-1273 | Case-control | NR | 14+ days | 18.852 | 85% (23% to 97%) |
| Yong-Xu, 2022 | Positive Covid-19 | Mixture of mRNA | Case-control | NR | NR | 2.551 | 25% (20% to 30%) |
| Fowlkes et al., 2022 | Positive Covid-19 | BNT162b2 | Cohort | 82 | 14 to 82 days | 9.949 | 31% (9% to 48%) |
| | | BNT162b2 | Cohort | 149 | 14 to 149 days | 14.541 | 59% (22% to 79%) |
| | | BNT162b2 | Cohort | 299 | 150+ days | 29.847 | 62% (-28% to 89%) |
| Hansen et al., 2021 | Positive Covid-19 | BNT162b2 | Cohort | 30 | 27 to 28 days | 12.806 | 55% (24% to 74%) |
| | | BNT162b2 | Cohort | 60 | 31 to 60 days | 15.944 | 16% (-21% to 42%) |
| | | BNT162b2 | Cohort | 90 | 61 to 90 days | 9.209 | 10% (-10% to 26%) |
| | | BNT162b2 | Cohort | 150 | 91 to 150 days | 9.133 | -77% (-95% to -60%) |
| | | mRNA-1273 | Cohort | 30 | 1 to 30 days | 37.321 | 37% (-70% to 76%) |
| | | mRNA-1273 | Cohort | 60 | 31 to 60 days | 27.219 | 30% (-41% to 65%) |
| | | mRNA-1273 | Cohort | 90 | 61 to 90 days | 15.459 | 4% (-31% to 30%) |
| | | mRNA-1273 | Cohort | 150 | 91 to 150 days | 10.612 | -39% (-62% to -20%) |
| Šmíd et al., 2022** | Positive Covid-19 | Mixture | Cohort | 60 | 0 to 2 months | 0.510 | 43% (42% to 44%) |
| | | Mixture | Cohort | 120 | 2+ months | 0.510 | 9% (8% to 10%) |
| | Severe Infection | Mixture | Cohort | 60 | 0 to 2 months | 7.143 | 45% (29% to 57%) |
| | | Mixture | Cohort | 120 | 2+ months | 4.082 | 29% (21% to 37%) |

*Data of full dose in Accorsi et al., 2022 calculated from crude data of number of infected individuals

**Smid et al., 2022 reports two full dose datasets, the one that exclude previous infection and with previous infection. We included the data that exclude previous infection for the analysis

*Studies whose data of Days Latitude are only day 7 or below and NR are excluded from the meta-regression analysis.

#### S.4. Summary of outcomes for booster dose

##### Table S5. Summary of outcomes for booster dose

| Authors | Endpoints | Vaccine | Design | Days Latitude | Follow-up Interval | SE of VE | VE (95%CI) |
|---|---|---|---|---|---|---|---|
| Accorsi et al., 2022 | Symptomatic Infection | BNT162b2 | Case-control | 30 | 7+ days | 1.531 | 65% (62% to 68%) |
| | | mRNA-1273 | Case-control | 30 | 7+ days | 1.276 | 72% (69% to 74%) |
| Andrews et al., 2022 | Symptomatic Infection | BNT162b2 | Case-control | 7 | 1 week | 0.383 | 67% (66% to 68%) |
| | | BNT162b2 | Case-control | 35 | 2 to 4 weeks | 0.332 | 67% (67% to 68%) |
| | | BNT162b2 | Case-control | 70 | 5 to 9 weeks | 0.408 | 55% (54% to 56%) |
| | | BNT162b2 | Case-control | 105 | 10+ week | 5.867 | 46% (45% to 68%) |
| | | ChAdOx1 nCov-19 | Case-control | 7 | 1 week | 8.597 | 58% (38% to 71%) |
| | | ChAdOx1 nCov-19 | Case-control | 35 | 2 to 4 weeks | 5.230 | 56% (44% to 65%) |
| | | ChAdOx1 nCov-19 | Case-control | 70 | 5 to 9 weeks | 5.714 | 47% (34% to 57%) |
| | | mRNA-1273 | Case-control | 7 | 1 week | 1.301 | 64% (62% to 67%) |
| | | mRNA-1273 | Case-control | 35 | 2 to 4 weeks | 1.276 | 65% (62% to 67%) |
| Buchan et al., 2021 | Severe infection | Mixture of mRNA | Case-control | 6 | 0 to 6 days | 6.633 | 91% (71% to 97%) |
| | | Mixture of mRNA | Case-control | 14 | 7+ days | 2.806 | 95% (87% to 98%) |
| | Symptomatic Infection | Mixture of mRNA | Case-control | 6 | 0 to 6 days | 3.571 | 36% (29% to 43%) |
| | | Mixture of mRNA | Case-control | 14 | 7+ days | 2.296 | 61% (56% to 65%) |
| Chemaitelly et al., 2022 | Symptomatic Infection | BNT162b2 | Case-control | 7 | 1 week | 7.015 | 16% (1% to 28%) |
| | | BNT162b2 | Case-control | 21 | 2 to 3 weeks | 6.097 | 53% (41% to 65%) |
| | | BNT162b2 | Case-control | 42 | 4 to 5 weeks | 2.781 | 57% (51% to 62%) |
| | | BNT162b2 | Case-control | 63 | 6 to 7 weeks | 3.138 | 46% (40% to 52%) |
| | | BNT162b2 | Case-control | 84 | 8 to 9 weeks | 4.694 | 38% (28% to 47%) |
| | | BNT162b2 | Case-control | 105 | 10 to 11 weeks | 5.051 | 44% (33% to 53%) |
| | | BNT162b2 | Case-control | 126 | 12+ weeks | 4.235 | 38% (29% to 45%) |
| | | mRNA-1273 | Case-control | 7 | 1 week | 15.332 | 4% (-31% to 29%) |

| | | | | | | | |
|---|---|---|---|---|---|---|---|
| | | mRNA-1273 | Case-control | 21 | 2 to 3 weeks | 5.638 | 53% (41% to 63%) |
| | | mRNA-1273 | Case-control | 42 | 4 to 5 weeks | 6.097 | 55% (41% to 65%) |
| | | mRNA-1273 | Case-control | 63 | 6+ weeks | 8.597 | 39% (19% to 53%) |
| Ferdinand et al., 2022 | Symptomatic Infection | Mixture of mRNA | Case-control | 60 | 0 to 2 months | 0.765 | 87% (85% to 88%) |
| | | Mixture of mRNA | Case-control | 180 | 2 to 3 months | 0.765 | 81% (79% to 82%) |
| | | Mixture of mRNA | Case-control | 240 | 3 to 4 months | 3.061 | 66% (59% to 71%) |
| | | Mixture of mRNA | Case-control | 300 | 5+ months | 30.102 | 31% (-50% to 68%) |
| | Severe infection | Mixture of mRNA | Case-control | 60 | 0 to 2 months | 1.276 | 91% (88% to 93%) |
| | | Mixture of mRNA | Case-control | 180 | 2 to 3 months | 1.276 | 88% (85% to 90%) |
| | | Mixture of mRNA | Case-control | 240 | 4+ months | 4.592 | 78% (67% to 85%) |
| Gray et al., 2021 | Positive Covid-19 | Ad26.COV2 | Case-control | 14 | 0 to 13 days | 3.567 | 32% (25% to 39%) |
| | | Ad26.COV2 | Case-control | 28 | 14 to 27 days | 3.674 | 14% (7% to 22%) |
| | | Ad26.COV2 | Case-control | 87 | 27 to 28 days | 6.857 | 6% (-6% to 20%) |
| | Severe Infection | Ad26.COV2 | Case-control | 13 | 0 to 13 days | 13.265 | 93% (47% to 99%) |
| | | Ad26.COV2 | Case-control | 27 | 14 to 27 days | 11.224 | 81% (49% to 93%) |
| Klein et al., 2022 | Symptomatic Infection (Age 5-11 years) | BNT162b2 | Case-control | 67 | 14 to 67 days | 8.929 | 51% (30% to 65%) |
| | Symptomatic Infection (Age 12-15 years) | BNT162b2 | Case-control | 149 | 14 to 149 days | 6.888 | 45% (30% to 57%) |
| | | BNT162b2 | Case-control | 299 | 150+ days | 10.714 | -2% (-25% to 17%) |
| | Symptomatic infection (Age 16-17 years) | BNT162b2 | Case-control | 149 | 14 to 149 days | 11.480 | 34% (8% to 53%) |
| | | BNT162b2 | Case-control | 299 | 150+ days | 12.245 | -3% (-30% to 18%) |
| Lauring et al., 2022 | Severe infection | Mixture of mRNA | Case-control | N/A | 7+ days | 4.337 | 86% (75% to 92%) |
| Monge et al., 2022 | Positive Covid-19 | Mixture | Case-control | 34 | 7 to 34 days | 0.561 | 51% (50% to 52%) |
| Natarajan et al., 2022 | Symptomatic Infection | Mixture | Case-control | 120 | 7 to 120 days | 0.510 | 83% (82% to 84%) |

| | | | | | | | |
|---|---|---|---|---|---|---|---|
| | Severe infection | Mixture | Case-control | 120 | 7 to 120 days | 0.765 | 90% (88% to 91%) |
| Tartof et al., 2022 | Severe infection | BNT162b2 | Case-control | 90 | 0 to 3 months | 2.296 | 89% (83% to 92%) |
| | | BNT162b2 | Case-control | 180 | 3 to 5 months | 10.459 | 90% (57% to 98%) |
| | Symptomatic Infection | BNT162b2 | Case-control | 90 | 6+ months | 2.296 | 78% (73% to 82%) |
| | | BNT162b2 | Case-control | 180 | 270+ days | 14.286 | 48% (13% to 69%) |
| Tenforde et al., 2022 | Severe infection | Mixture of mRNA | Case-control | 60 | 60 days (median) | 2.296 | 94% (88% to 97%) |
| Thompson et al., 2022 | Symptomatic Infection | Mixture of mRNA | Case-control | N/A | 14+ days | 1.276 | 82% (79% to 84%) |
| | Severe infection | Mixture of mRNA | Case-control | N/A | 14+ days | 3.571 | 90% (80% to 94%) |
| Tseng et al., 2022 | Severe infection | mRNA-1273 | Case-control | N/A | 14+ days | 6.046 | 99% (76% to 100%) |
| | Positive Covid-19 | mRNA-1273 | Case-control | 60 | 14 to 60 days | 0.944 | 72% (70% to 74%) |
| | | mRNA-1273 | Case-control | 120 | 60+ days | 3.342 | 51% (44% to 57%) |
| Yong-Xu, 2022 | Severe infection | Mixture of mRNA | Case-control | N/A | NR | 2.296 | 91% (85% to 94%) |
| Hansen et al., 2021 | Positive Covid-19 | BNT162b2 | Cohort | 30 | 1 to 30 days | 10.179 | 55% (31% to 70%) |
| Šmíd et al., 2022 | Symptomatic Infection | Mixture | Cohort | 60 | 60 to 120 days | 0.255 | 56% (55% to 56%) |
| | | Mixture | Cohort | 120 | 120+ days | 1.020 | 21% (19% to 23%) |
| | Severe infection | Mixture | Cohort | 60 | 60 to 120 days | 5.357 | 87% (73% to 94%) |
| | | Mixture | Cohort | 120 | 120+ days | 2.041 | 79% (75% to 83%) |

*Studies whose data of Days Latitude are N/A or less than 7 days are excluded from the analysis.

#### S.5. Summary of outcomes for booster vs full dose of the within 3 months or more model

##### Table S6. Summary of outcomes for booster vs full dose of the within 3 months or more model

| Authors | Outcomes | Design | Vaccine | VED (95%CI) | SE of VED |
|---|---|---|---|---|---|
| Abu-Raddad et al., 2022 | Symptomatic Infection | Cohort | BNT162b2 | 50.6% (50.18% to 51.02%) | 0.213 |
| Abu-Raddad et al., 2022 | Symptomatic Infection | Cohort | mRNA-1273 | 52.7% (51.53% to 53.87%) | 0.597 |
| Abu-Raddad et al., 2022 | Severe infection | Cohort | BNT162b2 | 23.5% (20.57% to 26.43%) | 1.497 |
| Accorsi et al., 2022 | Symptomatic Infection | Case-control | BNT162b2 | 64.9% (64.34% to 65.46%) | 0.284 |
| Accorsi et al., 2022 | Symptomatic Infection | Case-control | mRNA-1273 | 59.5% (58.81% to 60.19%) | 0.352 |
| Andrews et al., 2022 | Symptomatic Infection | Cohort | ChAdOx1 nCov-19 | 27.29% (16.16% to 38.43%) | 5.68 |
| Andrews et al., 2022 | Symptomatic Infection | Cohort | BNT162b2 | 28.15% (26.5% to 29.79%) | 0.841 |
| Andrews et al., 2022 | Symptomatic Infection | Cohort | mRNA-1273 | 33.96% (28.34% to 39.59%) | 2.869 |
| Buchan et al., 2021 | Symptomatic Infection | Case-control | mixture of mRNA | 35.85% (24.81% to 46.89%) | 5.633 |
| Buchan et al., 2021 | Severe infection | Case-control | mixture of mRNA | 24.21% (-33.37% to 81.79%) | 29.377 |
| Chemaitelly et al., 2022 | Symptomatic Infection | Case-control | mRNA-1273 | 6% (-14.52% to 26.51%) | 10.466 |
| Chemaitelly et al., 2022 | Symptomatic Infection | Case-control | BNT162b2 | 9.01% (-23.82% to 41.83%) | 16.748 |
| Ferdinand et al., 2022 | symptomatic Infection | Case-control | mixture of mRNA | 22.33% (16.9% to 27.77%) | 2.774 |

| | | | | | |
|---|---|---|---|---|---|
| Hansen et al., 2022 | Positive Covid-19 | Cohort | BNT162b2 | 78.54% (53.26% to 103.81%) | 12.895 |
| Klein et al., 2022 | symptomatic Infection | Case-control | BNT162b2 | 41.7% (21.43% to 61.98%) | 10.345 |
| Lauring et al., 2022 | Severe infection | Case-control | mixture of mRNA | 2% (-4.88% to 8.88%) | 3.512 |
| Natarajan et al., 2022 | Symptomatic Infection | Case-control | mixture of mRNA | 29% (26.41% to 31.59%) | 1.321 |
| Natarajan et al., 2022 | Severe infection | Case-control | mixture of mRNA | 17.5% (14.87% to 20.13%) | 1.342 |
| Šmíd et al., 2022 | Severe infection | Case-control | Mixture | 46% (35.35% to 56.65%) | 5.436 |
| Šmíd et al., 2022 | Positive Covid-19 | Case-control | Mixture | 1.5% (-3.94% to 6.94%) | 2.774 |
| Tartof et al., 2022 | Symptomatic Infection | Case-control | BNT162b2 | 33% (24.98% to 41.02%) | 4.09 |
| Tartof et al., 2022 | Severe infection | Case-control | BNT162b2 | 19% (-0.18% to 38.18%) | 9.788 |
| Tenforde et al., 2022 | Severe infection | Case-control | mixture of mRNA | 15% (12.88% to 17.12%) | 1.082 |
| Thompson et al., 2022 | Severe infection | Case-control | mixture of mRNA | 9% (-1.9% to 19.9%) | 5.563 |
| Thompson et al., 2022 | Positive Covid-19 | Case-control | mixture of mRNA | 30% (25.07% to 34.93%) | 2.516 |
| Tseng et al., 2022 | severe infection | Case-control | mRNA-1273 | 14.7% (7.49% to 21.91%) | 3.676 |
| Tseng et al., 2022 | Positive Covid-19 | Case-control | mRNA-1273 | 27.6% (20.4% to 34.8%) | 3.674 |
| Yong-Xu et al., 2022 | Positive Covid-19 | Case-control | mixture of mRNA | 21% (20.14% to 21.86%) | 0.437 |

#### S.6. Summary of outcomes for booster vs full dose of the within 3 months model

##### Table S7. Summary of outcomes for booster vs full dose of the within 3 months model

| Authors | Outcomes | Design | Vaccine | VED (95% CI) | SE_diff |
|---|---|---|---|---|---|
| Andrews et al., 2022 | Symptomatic Infection | Cohort | ChAdOx1 nCov-19 | 16.11% (2.41 to 29.82%) | 6.99 |
| Andrews et al., 2022 | Symptomatic Infection | Cohort | BNT162b2 | 17.64% (16.24 to 19.05%) | 0.717 |
| Andrews et al., 2022 | Symptomatic Infection | Cohort | mRNA-1273 | 16.52% (12.45 to 20.59%) | 2.075 |
| Buchan et al., 2021 | Symptomatic Infection | Case-control | mixture of mRNA | 27.25% (-5.06 to 59.56%) | 16.485 |
| Chemaitelly et al., 2022 | Symptomatic Infection | Case-control | BNT162b2 | 0% (-6.08 to 6.08%) | 3.103 |
| Chemaitelly et al., 2022 | Symptomatic Infection | Case-control | mRNA-1273 | 33% (-21.38 to 87.38%) | 27.744 |
| Tartof et al., 2022 | Symptomatic Infection | Case-control | BNT162b2 | -5.72% (-19.59 to 8.15%) | 7.077 |
| Abu-Raddad et al., 2022 | Symptomatic Infection | Cohort | BNT162b2 | -7.7% (-38.69 to 23.29%) | 15.812 |
| Abu-Raddad et al., 2022 | Symptomatic Infection | Cohort | mRNA-1273 | 2% (-8.26 to 12.26%) | 5.234 |
| Natarajan et al., 2022 | Symptomatic Infection | Case-control | mixture of mRNA | 27.6% (19.15 to 36.06%) | 4.314 |
| Klein et al., 2022 | symptomatic Infection | Case-control | BNT162b2 | 33% (23.82 to 42.18%) | 4.683 |
| Ferdinand et al., 2022 | symptomatic Infection | Case-control | mixture of mRNA | 19% (-2.97 to 40.97%) | 11.207 |
| Buchan et al., 2021 | Severe infection | Case-control | mixture of mRNA | -3.1% (-6.29 to 0.09%) | 1.625 |

| | | | | | |
|---|---|---|---|---|---|
| Lauring et al., 2022 | Severe infection | Case-control | mixture of mRNA | -3.5% (-6.45 to -0.55%) | 1.507 |
| Tartof et al., 2022 | Severe infection | Case-control | BNT162b2 | 42% (32 to 52%) | 5.102 |
| Smid et al., 2022 | Severe infection | Case-control | Mixture | 3.5% (0.17 to 6.83%) | 1.697 |
| Abu-Raddad et al., 2022 | Severe infection | Cohort | BNT162b2 | 50.6% (48.35 to 52.85%) | 1.148 |
| Natarajan et al., 2022 | Severe infection | Case-control | mixture of mRNA | 23.5% (7.7 to 39.3%) | 8.061 |
| Tenforde et al., 2022 | Severe infection | Case-control | mixture of mRNA | 52.7% (46.4 to 59%) | 3.214 |
| Hansen et al., 2022 | Positive Covid-19 | Cohort | BNT162b2 | 29% (18.95 to 39.05%) | 5.127 |
| Tseng et al., 2022 | Positive Covid-19 | Case-control | mRNA-1273 | 17.5% (7.26 to 27.74%) | 5.225 |
| Monge et al., 2022 | Positive Covid-19 | Case-control | mRNA-1273 | 15% (3.58 to 26.42%) | 5.828 |
| Monge et al., 2022 | Positive Covid-19 | Design | BNT162b2 | 30% (12.93 to 47.07%) | 8.71 |
| Smid et al., 2022 | Positive Covid-19 | Cohort | Mixture | 22.33% (16.11 to 28.56%) | 3.176 |

#### S.8. Detailed Results of VE Estimates and VE Reduction of Full Dose

Table S9. Detailed results of VE estimates and VE reduction of full dose for any infection outcome

| Variable | Overall | Report |
|---|---|---|
| Results | Mixed-Effects Model (k = 93; tau^2 estimator: REML)<br><br>logLik deviance AIC BIC AICc<br>-434.8900 869.7800 875.7800 883.3125 876.0558<br><br>tau^2 (estimated amount of residual heterogeneity): 741.3064 (SE = 118.5740)<br>tau (square root of estimated tau^2 value): 27.2269<br>I^2 (residual heterogeneity / unaccounted variability): 99.50%<br>H^2 (unaccounted variability / sampling variability): 199.94<br>R^2 (amount of heterogeneity accounted for): 9.91%<br><br>Test for Residual Heterogeneity:<br>QE(df = 91) = 7954.7133, p-val < .0001<br><br>Test of Moderators (coefficient 2):<br>F(df1 = 1, df2 = 91) = 8.9517, p-val = 0.0036<br><br>Model Results:<br><br>estimate se tval df pval ci.lb ci.ub<br>intrcpt 50.1200 6.2132 8.0667 91 <.0001 37.7783 62.4618 ***<br>Days -0.0985 0.0329 -2.9919 91 0.0036 -0.1640 -0.0331 ** | R^2 = 9.91%<br>VE estimate (%):<br>50.12 (37.78 to 62.46), p:<.0001<br>VE reduction estimate (% per month):<br>-2.955 (-4.92 to -0.993), p:0.0036<br><br>VE (%) = 50.12 - 0.0985 day |

| | | |
|---|---|---|
| Reg Plot | 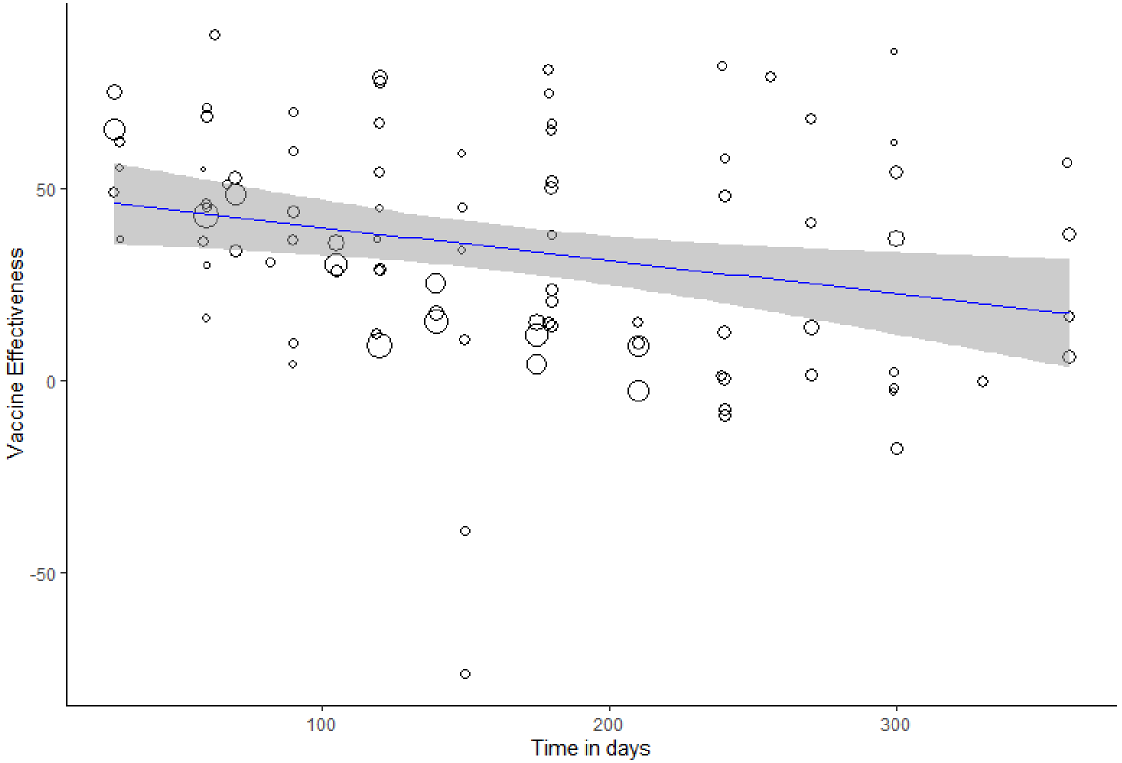 | |
| Variable | Ad26.COV2.S | |
| Results | Number of studies combined: k = 2<br><br>VE 95%-CI t p-value<br>Random effects model 59.94 [-22.34; 142.22] 9.26 0.0685<br><br>Quantifying heterogeneity:<br>tau^2 = 0.0051; tau = 0.0715<br>I^2 = 60.5% [0.0%; 90.8%]; H = 1.59 [1.00; 3.30] | I^2 = 60.5%<br>VE estimate (%):<br>59.94 (-22.34 to 142.22), p:0.0685<br>VE reduction estimate (% per month):<br>N/A |

| | | |
|---|---|---|
| | Test of heterogeneity:<br>Q d.f. p-value<br>2.53 1 0.1114 | |
| Reg Plot | N/A | |
| Variable | BNT162b2 | |
| Results | Mixed-Effects Model (k = 38; tau^2 estimator: REML)<br><br>logLik deviance AIC BIC AICc<br>-171.6450 343.2899 349.2899 354.0405 350.0399<br><br>tau^2 (estimated amount of residual heterogeneity): 717.3130 (SE = 183.0239)<br>tau (square root of estimated tau^2 value): 26.7827<br>I^2 (residual heterogeneity / unaccounted variability): 99.45%<br>H^2 (unaccounted variability / sampling variability): 181.89<br>R^2 (amount of heterogeneity accounted for): 24.08%<br><br>Test for Residual Heterogeneity:<br>QE(df = 36) = 1269.8612, p-val < .0001<br><br>Test of Moderators (coefficient 2):<br>F(df1 = 1, df2 = 36) = 10.4159, p-val = 0.0027<br><br>Model Results:<br><br>estimate se tval df pval ci.lb ci.ub<br>intrcpt 54.6182 9.3902 5.8165 36 <.0001 35.5739 73.6624 ***<br>Days -0.1607 0.0498 -3.2274 36 0.0027 -0.2617 -0.0597 ** | R^2 = 24.08%<br>VE estimate (%):<br>54.62 (35.57 to 73.66), p:<.0001<br>VE reduction estimate (% per month):<br>-4.821 (-7.851 to -1.791), p:0.0027<br><br>VE (%) = 54.6182 - 0.1607 day |

| | | |
|---|---|---|
| Reg Plot | 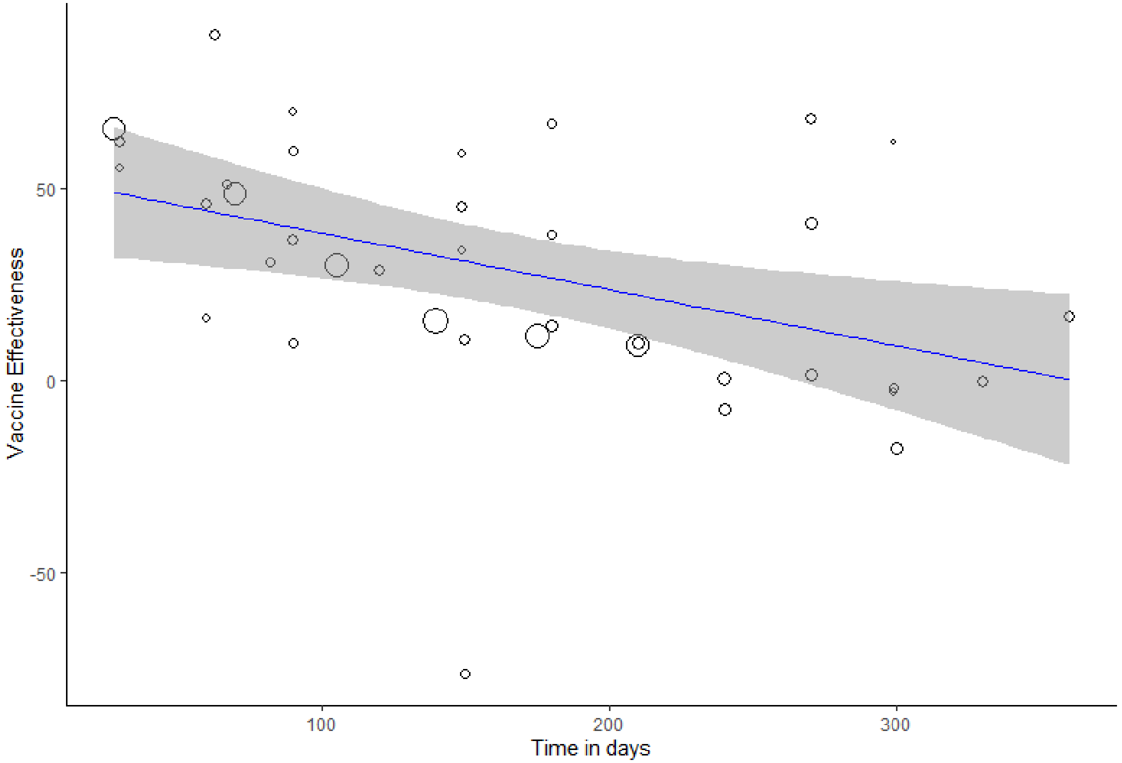 | |
| Variable | ChAdOx1 nCov-19 | |
| Results | Mixed-Effects Model (k = 6; tau^2 estimator: REML)<br><br>logLik deviance AIC BIC AICc<br>-9.6120 19.2241 25.2241 23.3830 49.2241<br><br>tau^2 (estimated amount of residual heterogeneity): 2.8159 (SE = 4.7880)<br>tau (square root of estimated tau^2 value): 1.6781<br>I^2 (residual heterogeneity / unaccounted variability): 46.23%<br>H^2 (unaccounted variability / sampling variability): 1.86<br>R^2 (amount of heterogeneity accounted for): 99.22% | R^2 = 99.22%<br>VE estimate (%):<br>56.03 (47.81 to 64.25), p:<.0001<br>VE reduction estimate (% per month):<br>-8.532 (-9.978 to -7.089), p:<.0001<br><br>VE (%) = 56.0307 - 0.2844 day |

| | |
|---|---|
| | Test for Residual Heterogeneity:<br>QE(df = 4) = 6.9972, p-val = 0.1360<br><br>Test of Moderators (coefficient 2):<br>F(df1 = 1, df2 = 4) = 269.3835, p-val < .0001<br><br>Model Results:<br><br>estimate se tval df pval ci.lb ci.ub<br>intrcpt 56.0307 2.9616 18.9192 4 <.0001 47.8081 64.2534 ***<br>Days -0.2844 0.0173 -16.4129 4 <.0001 -0.3326 -0.2363 *** |
| Reg Plot | 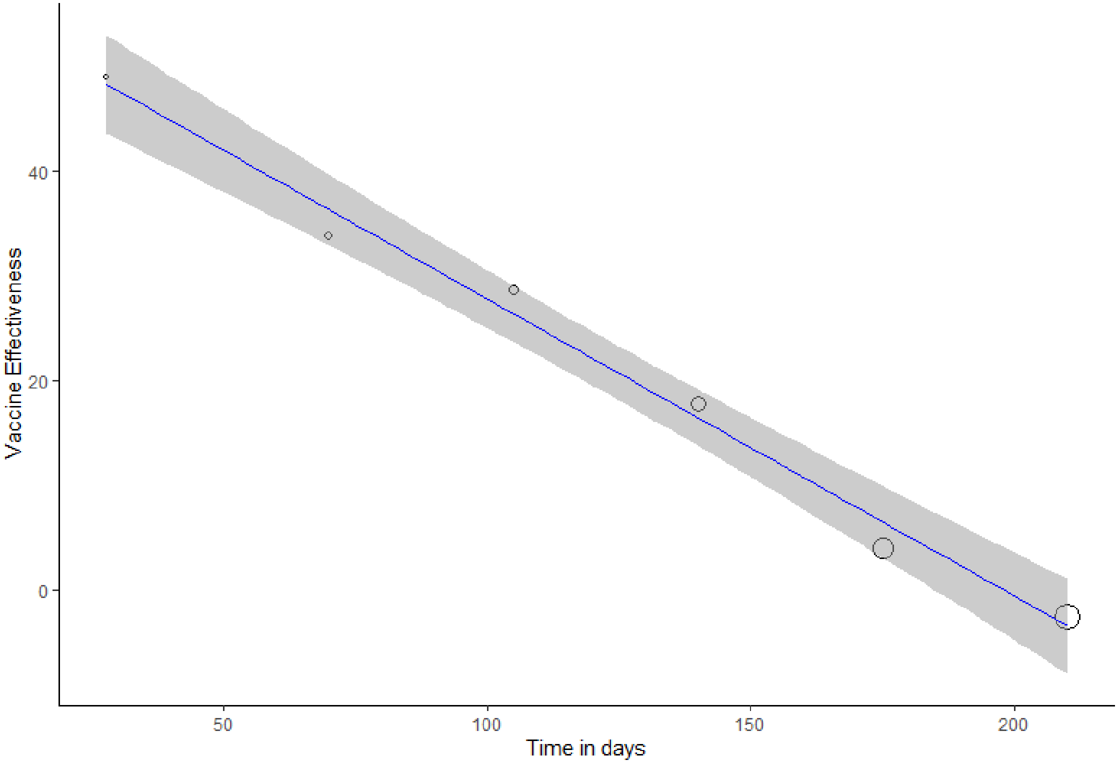 |
| Variable | Mixture |

| | | |
|---|---|---|
| Results | Mixed-Effects Model (k = 4; tau^2 estimator: REML)<br><br>logLik deviance AIC BIC AICc<br>-7.5277 15.0555 21.0555 17.1349 45.0555<br><br>tau^2 (estimated amount of residual heterogeneity): 99.6973 (SE = 115.8024)<br>tau (square root of estimated tau^2 value): 9.9849<br>I^2 (residual heterogeneity / unaccounted variability): 88.68%<br>H^2 (unaccounted variability / sampling variability): 8.84<br>R^2 (amount of heterogeneity accounted for): 63.29%<br><br>Test for Residual Heterogeneity:<br>QE(df = 2) = 23.7186, p-val < .0001<br><br>Test of Moderators (coefficient 2):<br>F(df1 = 1, df2 = 2) = 6.1503, p-val = 0.1313<br><br>Model Results:<br><br>estimate se tval df pval ci.lb ci.ub<br>intrcpt 69.3531 16.5628 4.1873 2 0.0526 -1.9107 140.6170 .<br>Days -0.4259 0.1717 -2.4800 2 0.1313 -1.1649 0.3130 | R^2 = 63.29%<br>VE estimate (%):<br>69.35 (-1.91 to 140.62), p:0.0526<br>VE reduction estimate (% per month):<br>-12.777 (-34.947 to 9.39), p:0.1313<br><br>VE (%) = 69.3531 - 0.4259 day |

| Reg Plot | 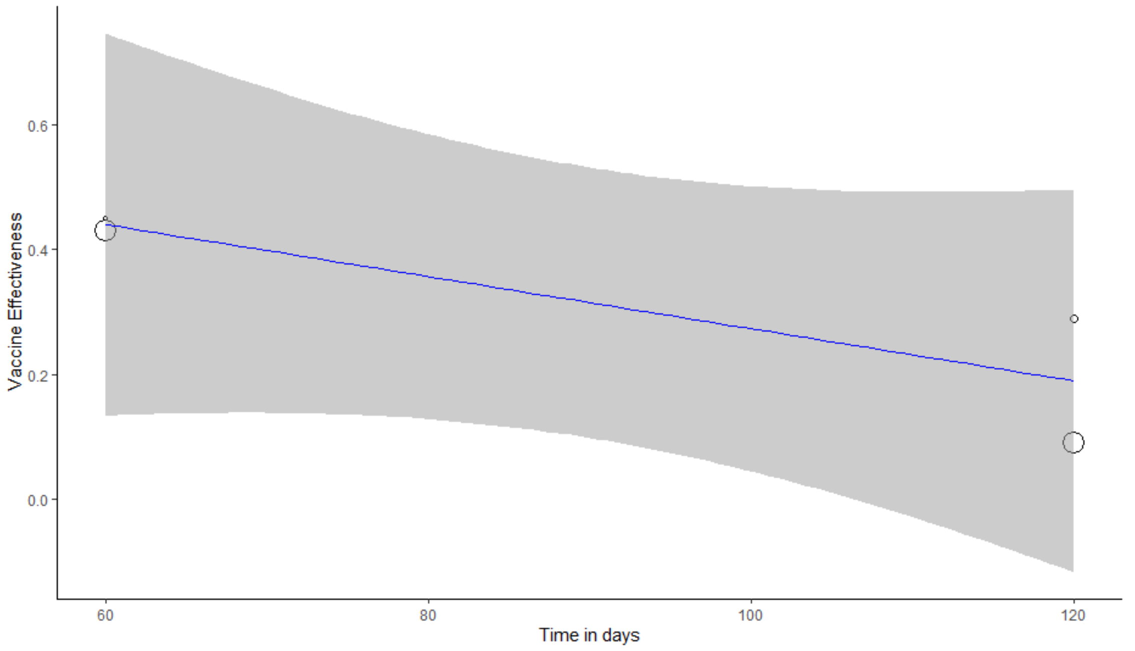 | |
|---|---|---|
| Variable | Mixture of mRNA vaccines | |
| Results | Mixed-Effects Model (k = 25; tau^2 estimator: REML)<br><br>logLik deviance AIC BIC AICc<br>-108.0914 216.1827 222.1827 225.5892 223.4459<br><br>tau^2 (estimated amount of residual heterogeneity): 613.3061 (SE = 201.1280)<br>tau (square root of estimated tau^2 value): 24.7650<br>I^2 (residual heterogeneity / unaccounted variability): 97.50%<br>H^2 (unaccounted variability / sampling variability): 40.05<br>R^2 (amount of heterogeneity accounted for): 0.00%<br><br>Test for Residual Heterogeneity:<br>QE(df = 23) = 628.4139, p-val < .0001 | R^2 = 0%<br>VE estimate (%):<br>62.47 (36.03 to 88.91), p:<.0001<br>VE reduction estimate (% per month):<br>-1.536 (-5.142 to 2.067), p:0.3868<br><br>VE (%) = 62.4733 - 0.0512 day |

| | | |
|---|---|---|
| | Test of Moderators (coefficient 2):<br>F(df1 = 1, df2 = 23) = 0.7783, p-val = 0.3868<br><br>Model Results:<br><br>estimate se tval df pval ci.lb ci.ub<br>intrcpt 62.4733 12.7813 4.8879 23 <.0001 36.0331 88.9135 ***<br>Days -0.0512 0.0581 -0.8822 23 0.3868 -0.1714 0.0689 | |
| Reg Plot | 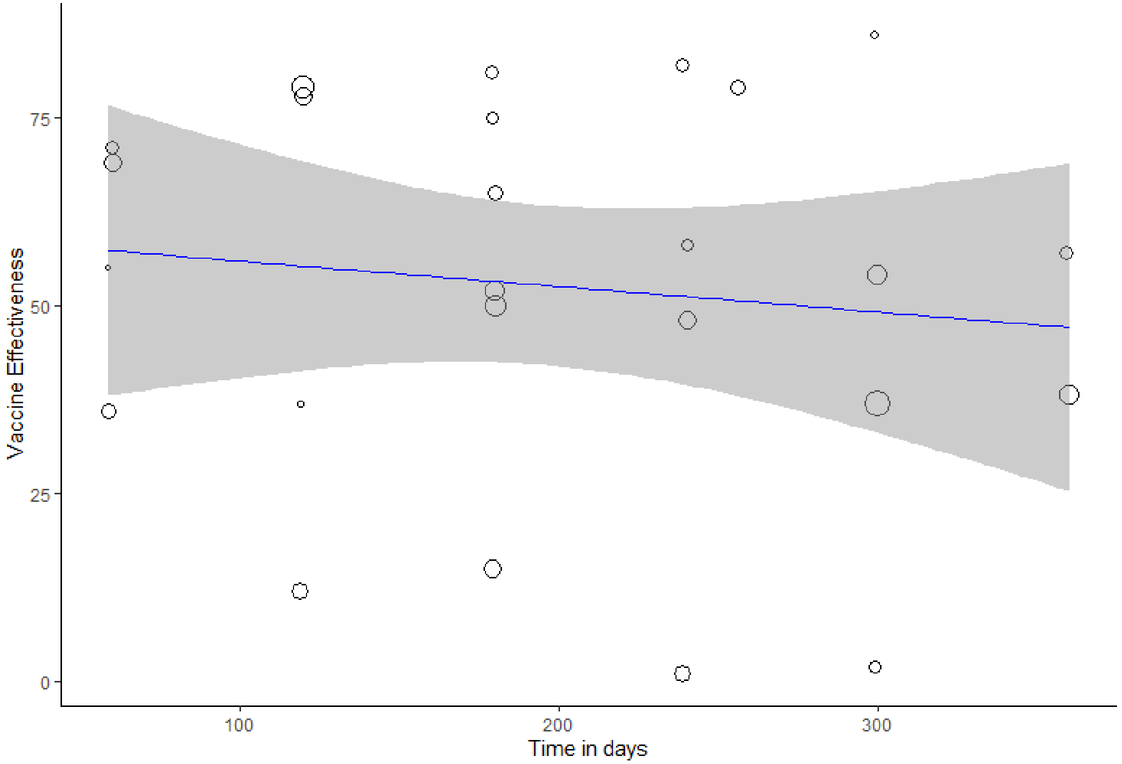 | |
| Variable | mRNA-1273 | |
| Results | Mixed-Effects Model (k = 18; tau^2 estimator: REML)<br><br>logLik deviance AIC BIC AICc | R^2 = 42.33%<br>VE estimate (%): |

| | | |
|---|---|---|
| | -71.7588 143.5177 149.5177 151.8354 151.5177<br><br>tau^2 (estimated amount of residual heterogeneity): 346.2541 (SE = 140.5909)<br>tau (square root of estimated tau^2 value): 18.6079<br>I^2 (residual heterogeneity / unaccounted variability): 98.12%<br>H^2 (unaccounted variability / sampling variability): 53.13<br>R^2 (amount of heterogeneity accounted for): 42.33%<br><br>Test for Residual Heterogeneity:<br>QE(df = 16) = 344.1080, p-val < .0001<br><br>Test of Moderators (coefficient 2):<br>F(df1 = 1, df2 = 16) = 9.7084, p-val = 0.0067<br><br>Model Results:<br><br>estimate se tval df pval ci.lb ci.ub<br>intrcpt 51.8048 10.5712 4.9006 16 0.0002 29.3949 74.2146 ***<br>Days -0.1780 0.0571 -3.1158 16 0.0067 -0.2991 -0.0569 ** | 51.8 (29.39 to 74.21), p:0.0002<br>VE reduction estimate (% per month):<br>-5.34 (-8.973 to -1.707), p:0.0067<br><br>VE (%) = 51.8048 - 0.178 day |

| | |
|---|---|
| Reg Plot | 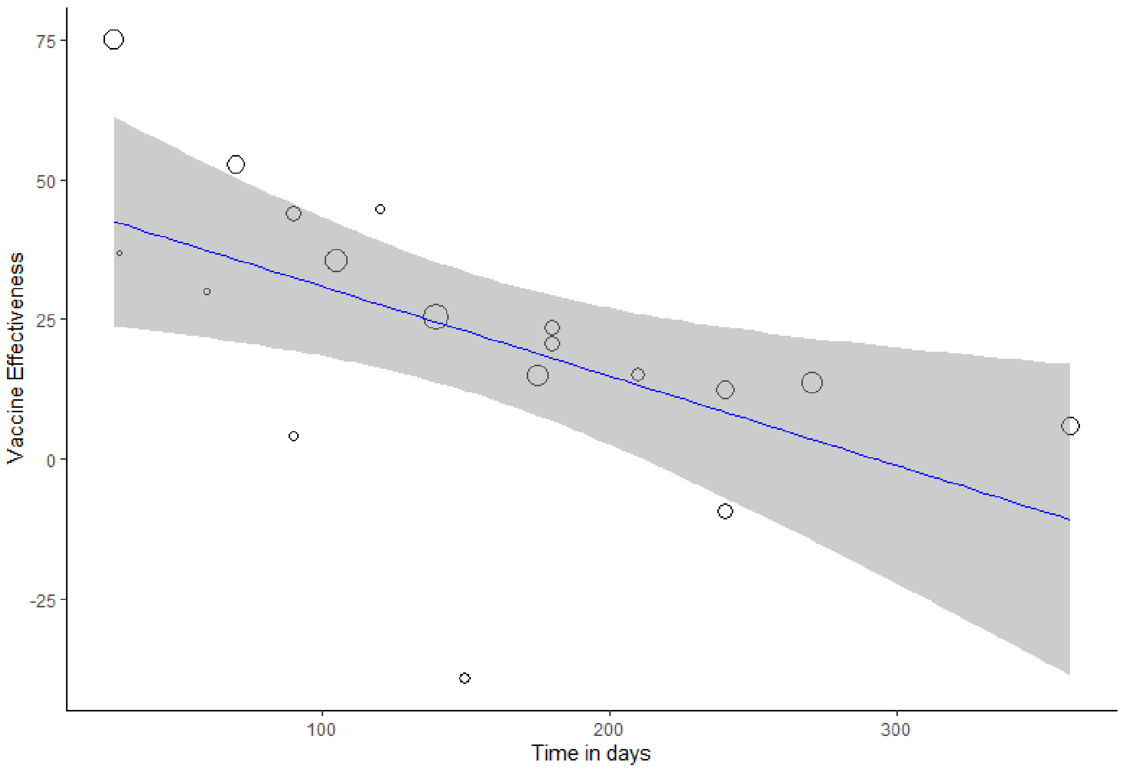 |

Cell is denoted as **green** if meta-regression is performed (i.e., if k > 3). Cell is denoted as **yellow** if the result is calculated from meta-analysis of two reports (k = 2).

Table S10. Detailed results of VE estimates and VE reduction of full dose for severe infection outcome

| Variable | overall | |
|---|---|---|
| Results | Mixed-Effects Model (k = 15; tau^2 estimator: REML)<br><br>logLik deviance AIC BIC AICc<br>-54.6842 109.3685 115.3685 117.0633 118.0351<br><br>tau^2 (estimated amount of residual heterogeneity): 235.8090 (SE = 108.9539)<br>tau (square root of estimated tau^2 value): 15.3561<br>I^2 (residual heterogeneity / unaccounted variability): 88.26%<br>H^2 (unaccounted variability / sampling variability): 8.52<br>R^2 (amount of heterogeneity accounted for): 0.00%<br><br>Test for Residual Heterogeneity:<br>QE(df = 13) = 145.4933, p-val < .0001<br><br>Test of Moderators (coefficient 2):<br>F(df1 = 1, df2 = 13) = 0.1494, p-val = 0.7053<br><br>Model Results:<br><br>estimate se tval df pval ci.lb ci.ub<br>intrcpt 68.2800 9.0585 7.5377 13 <.0001 48.7104 87.8497 ***<br>Days -0.0178 0.0460 -0.3865 13 0.7053 -0.1172 0.0817 | R^2 = 0%<br>VE estimate (%):<br>68.28 (48.71 to 87.85), p:<.0001<br>VE reduction estimate (% per month):<br>-0.534 (-3.516 to 2.451), p:0.7053<br><br>VE (%) = 68.28 - 0.0178 day |

| | | |
|---|---|---|
| Reg Plot | 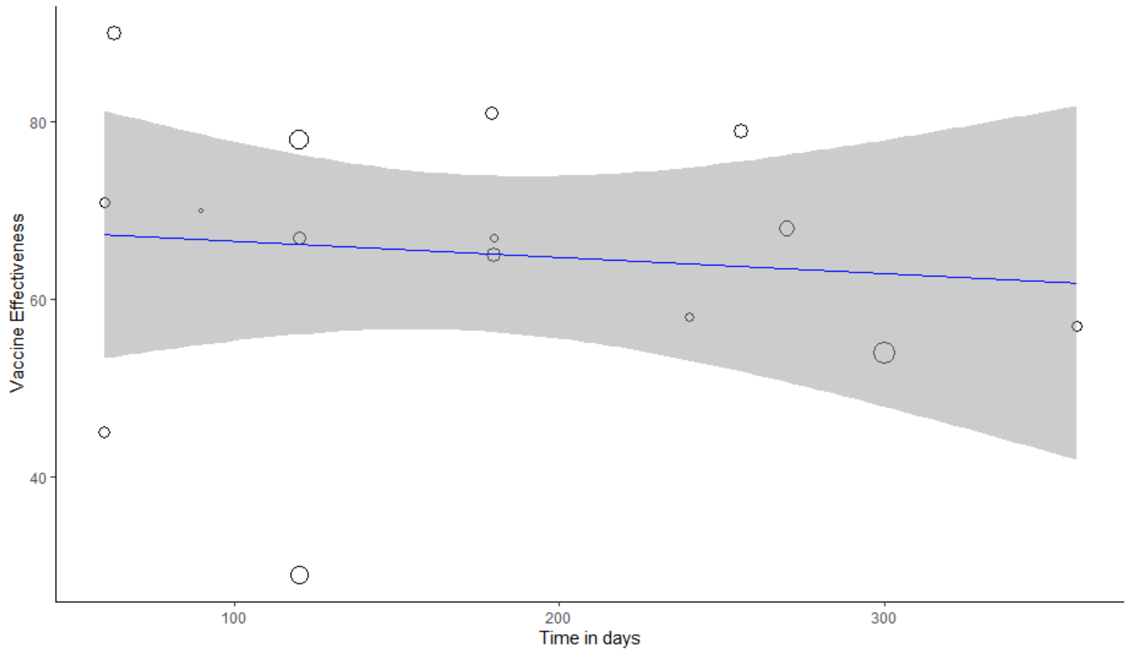 | |
| Variable | Ad26.COV2.S | |
| Results | VE estimate (%): 67 (52 to 77) | VE estimate (%): 67 (52 to 77)<br>VE reduction estimate (% per month): N/A |
| Reg Plot | N/A | |
| Variable | BNT162b2 | |
| Results | Mixed-Effects Model (k = 4; tau^2 estimator: REML)<br><br>tau^2 (estimated amount of residual heterogeneity): 37.0869 (SE = 110.2998)<br>tau (square root of estimated tau^2 value): 6.0899<br>I^2 (residual heterogeneity / unaccounted variability): 33.62%<br>H^2 (unaccounted variability / sampling variability): 1.51<br>R^2 (amount of heterogeneity accounted for): 63.40% | R^2 = 63.4%<br>VE estimate (%):<br>89.88 (50.39 to 129.38), p:0.0103<br>VE reduction estimate (% per month):<br>-2.715 (-9.177 to 3.747), p:0.2125<br><br>VE (%) = 89.8824 - 0.0905 day |

| | |
|---|---|
| | Test for Residual Heterogeneity:<br>QE(df = 2) = 2.9009, p-val = 0.2345<br><br>Test of Moderators (coefficient 2):<br>F(df1 = 1, df2 = 2) = 3.2663, p-val = 0.2125<br><br>Model Results:<br><br>estimate se tval df pval ci.lb ci.ub<br>intrcpt 89.8824 9.1794 9.7918 2 0.0103 50.3867 129.3781 *<br>Days -0.0905 0.0501 -1.8073 2 0.2125 -0.3059 0.1249 |
| Reg Plot | 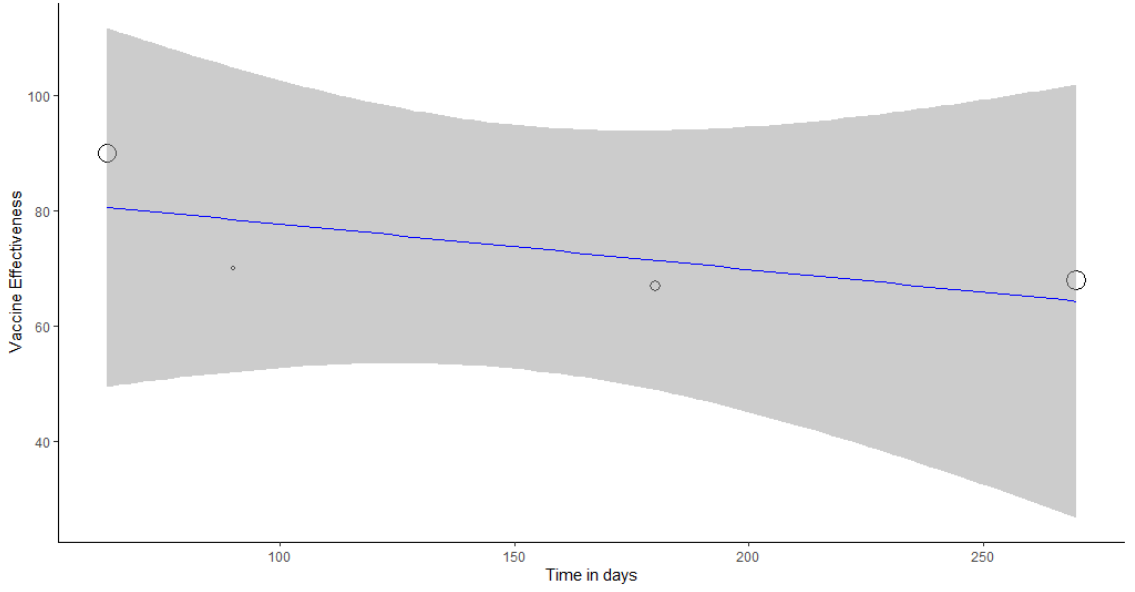 |
| Variable | ChAdOx1 nCov-19 |
| Results | N/A |
| Reg Plot | N/A |

| | | |
|---|---|---|
| Variable | mixture | |
| Results | Number of studies combined: k = 2<br><br>VE 95%-CI t p-value<br>Random effects model 35.9262 [-64.8036; 136.6561] 4.53 0.1383<br><br>Quantifying heterogeneity:<br>tau^2 = 94.1599; tau = 9.7036<br>I^2 = 73.6% [0.0%; 94.0%]; H = 1.94 [1.00; 4.10]<br><br>Test of heterogeneity:<br>Q d.f. p-value<br>3.78 1 0.0518 | I^2 = 73.6%<br>VE estimate (%):<br>35.9262 (-64.8036 to 136.6561), p: 0.1383<br>VE reduction estimate (% per month):<br>N/A |
| Reg Plot | N/A | |
| Variable | Mixture of mRNA | |
| Results | Mixed-Effects Model (k = 8; tau^2 estimator: REML)<br><br>logLik deviance AIC BIC AICc<br>-22.0809 44.1618 50.1618 49.5370 62.1618<br><br>tau^2 (estimated amount of residual heterogeneity): 59.2136 (SE = 53.6906)<br>tau (square root of estimated tau^2 value): 7.6950<br>I^2 (residual heterogeneity / unaccounted variability): 66.06%<br>H^2 (unaccounted variability / sampling variability): 2.95<br>R^2 (amount of heterogeneity accounted for): 38.23%<br><br>Test for Residual Heterogeneity:<br>QE(df = 6) = 18.3740, p-val = 0.0054<br><br>Test of Moderators (coefficient 2): | R^2 = 38.23%<br>VE estimate (%):<br>83.9 (62.22 to 105.58), p:<.0001<br>VE reduction estimate (% per month):<br>-2.229 (-5.061 to 0.603), p:0.1023<br><br>VE (%) = 83.8956 - 0.0743 day |

| | |
|---|---|
| | F(df1 = 1, df2 = 6) = 3.7116, p-val = 0.1023<br><br>Model Results:<br><br>estimate se tval df pval ci.lb ci.ub<br>intrcpt 83.8956 8.8601 9.4689 6 <.0001 62.2156 105.5755 ***<br>Days -0.0743 0.0386 -1.9266 6 0.1023 -0.1687 0.0201 |
| Reg Plot | 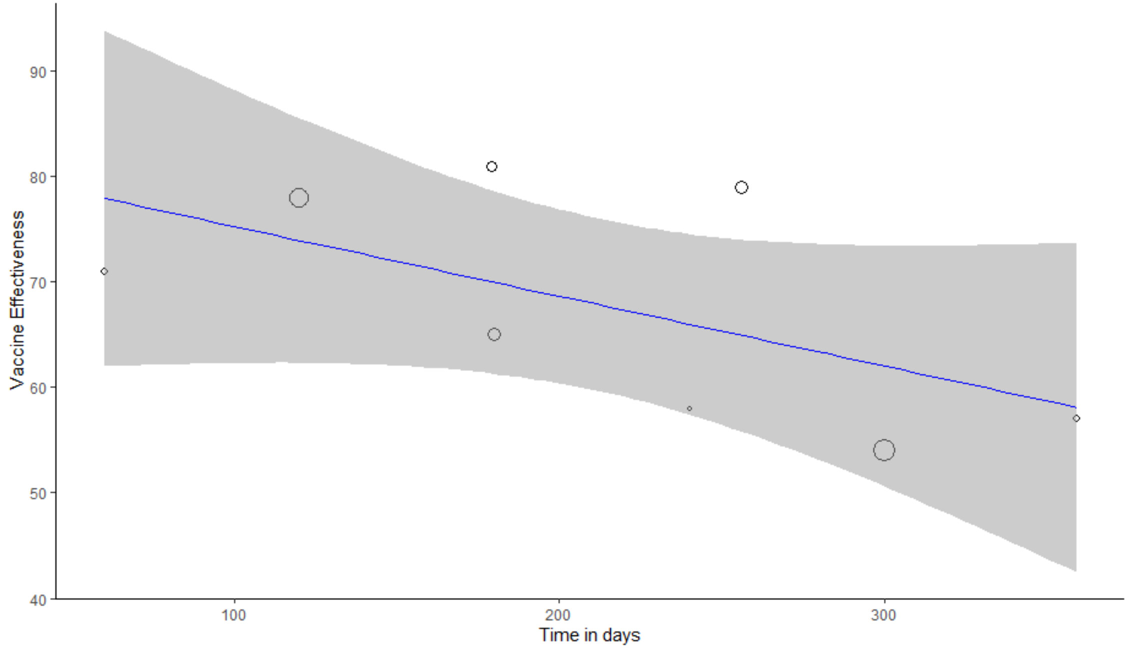 |
| Variable | mRNA-1273 |
| Results | N/A |
| Reg Plot | N/A |

Cell is denoted as **green** if meta-regression is performed (i.e., if k > 3). Cell is denoted as **yellow** if the result is calculated from meta-analysis of two report (k = 2). Cell is denoted as **orange** if the result is from a report (k = 1) which stays the same as what has been reported by the original study. Cell is denoted as **red** if the result has not been reported by any studies (k = 0).

Table S11. Detailed results of VE estimates and VE reduction of full dose for symptomatic infection outcome

| Variable | overall | |
|---|---|---|
| Results | Mixed-Effects Model (k = 59; tau^2 estimator: REML)<br><br>logLik deviance AIC BIC AICc<br>-255.0308 510.0617 516.0617 522.1908 516.5145<br><br>tau^2 (estimated amount of residual heterogeneity): 380.0117 (SE = 77.7305)<br>tau (square root of estimated tau^2 value): 19.4939<br>I^2 (residual heterogeneity / unaccounted variability): 99.08%<br>H^2 (unaccounted variability / sampling variability): 108.75<br>R^2 (amount of heterogeneity accounted for): 37.44%<br><br>Test for Residual Heterogeneity:<br>QE(df = 57) = 3743.4931, p-val < .0001<br><br>Test of Moderators (coefficient 2):<br>F(df1 = 1, df2 = 57) = 29.5771, p-val < .0001<br><br>Model Results:<br><br>estimate se tval df pval ci.lb ci.ub<br>intrcpt 58.2061 5.9668 9.7551 57 <.0001 46.2579 70.1543 ***<br>Days -0.1734 0.0319 -5.4385 57 <.0001 -0.2372 -0.1095 *** | R^2 = 37.44%<br>VE estimate (%):<br>58.21 (46.26 to 70.15), p:<.0001<br>VE reduction estimate (% per month):<br>-5.202 (-7.116 to -3.285), p:<.0001<br><br>VE (%) = 58.2061 - 0.1734 day |

| | | |
|---|---|---|
| Reg Plot | 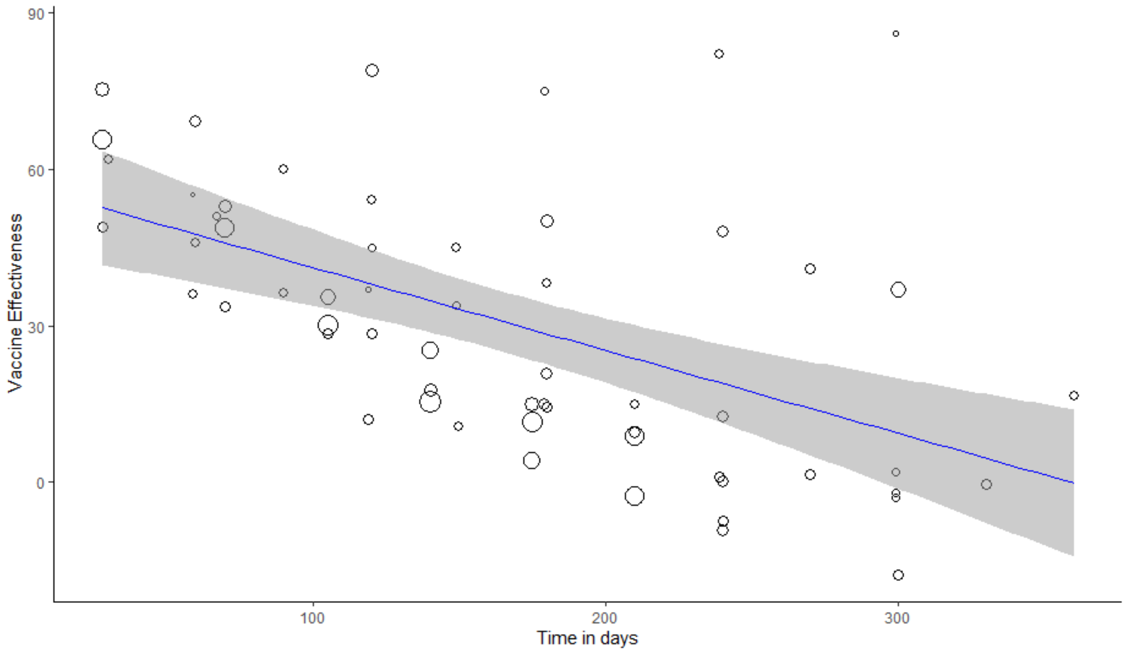 | |
| Variable | Ad26.COV2.S | |
| Results | VE estimate (%): 54 (43 to 63) | VE estimate (%): 54 (43 to 63)<br>VE reduction estimate (% per month): N/A |
| Reg Plot | N/A | |
| Variable | BNT162b2 | |
| Results | Mixed-Effects Model (k = 27; tau^2 estimator: REML)<br><br>logLik deviance AIC BIC AICc<br>-102.4504 204.9008 210.9008 214.5574 212.0437<br><br>tau^2 (estimated amount of residual heterogeneity): 185.0191 (SE = 60.1933)<br>tau (square root of estimated tau^2 value): 13.6022 | R^2 = 64.37%<br>VE estimate (%):<br>58.38 (46.09 to 70.67), p:<.0001<br>VE reduction estimate (% per month):<br>-5.91 (-7.761 to -4.056), p:<.0001<br><br>VE (%) = 58.3787 - 0.197 day |

| | |
|---|---|
| | I^2 (residual heterogeneity / unaccounted variability): 98.50%<br>H^2 (unaccounted variability / sampling variability): 66.78<br>R^2 (amount of heterogeneity accounted for): 64.37%<br><br>Test for Residual Heterogeneity:<br>QE(df = 25) = 760.6262, p-val < .0001<br><br>Test of Moderators (coefficient 2):<br>F(df1 = 1, df2 = 25) = 43.1650, p-val < .0001<br><br>Model Results:<br><br>estimate se tval df pval ci.lb ci.ub<br>intrcpt 58.3787 5.9674 9.7829 25 <.0001 46.0886 70.6687 ***<br>Days -0.1970 0.0300 -6.5700 25 <.0001 -0.2587 -0.1352 *** |

| | | |
|---|---|---|
| Reg Plot | 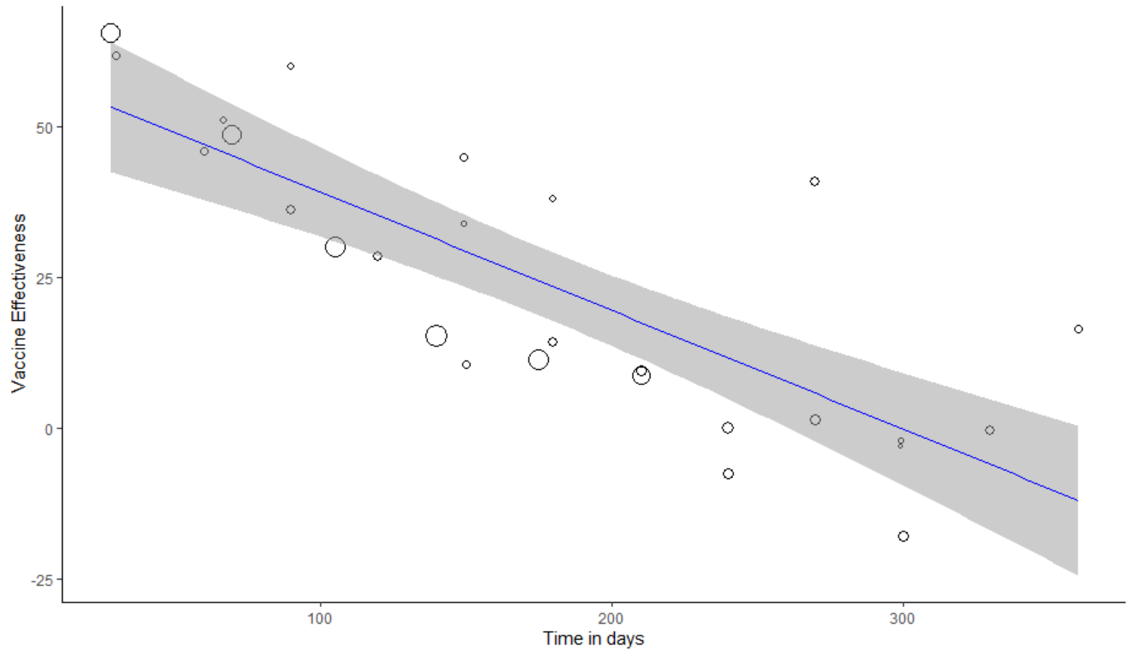<br> | |
| Variable | ChAdOx1 nCov-19 | |
| Results | Mixed-Effects Model (k = 6; tau^2 estimator: REML)<br><br>logLik deviance AIC BIC AICc<br>-9.6120 19.2241 25.2241 23.3830 49.2241<br><br>tau^2 (estimated amount of residual heterogeneity): 2.8159 (SE = 4.7880)<br>tau (square root of estimated tau^2 value): 1.6781<br>I^2 (residual heterogeneity / unaccounted variability): 46.23%<br>H^2 (unaccounted variability / sampling variability): 1.86<br>R^2 (amount of heterogeneity accounted for): 99.22%<br><br>Test for Residual Heterogeneity: | R^2 = 99.22%<br>VE estimate (%):<br>56.03 (47.81 to 64.25), p:<.0001<br>VE reduction estimate (% per month):<br>-8.532 (-9.978 to -7.089), p:<.0001<br><br>VE (%) = 56.0307 - 0.2844 day |

| | |
|---|---|
| | QE(df = 4) = 6.9972, p-val = 0.1360<br><br>Test of Moderators (coefficient 2):<br>F(df1 = 1, df2 = 4) = 269.3835, p-val < .0001<br><br>Model Results:<br><br>estimate se tval df pval ci.lb ci.ub<br>intrcpt 56.0307 2.9616 18.9192 4 <.0001 47.8081 64.2534 ***<br>Days -0.2844 0.0173 -16.4129 4 <.0001 -0.3326 -0.2363 *** |
| Reg Plot | 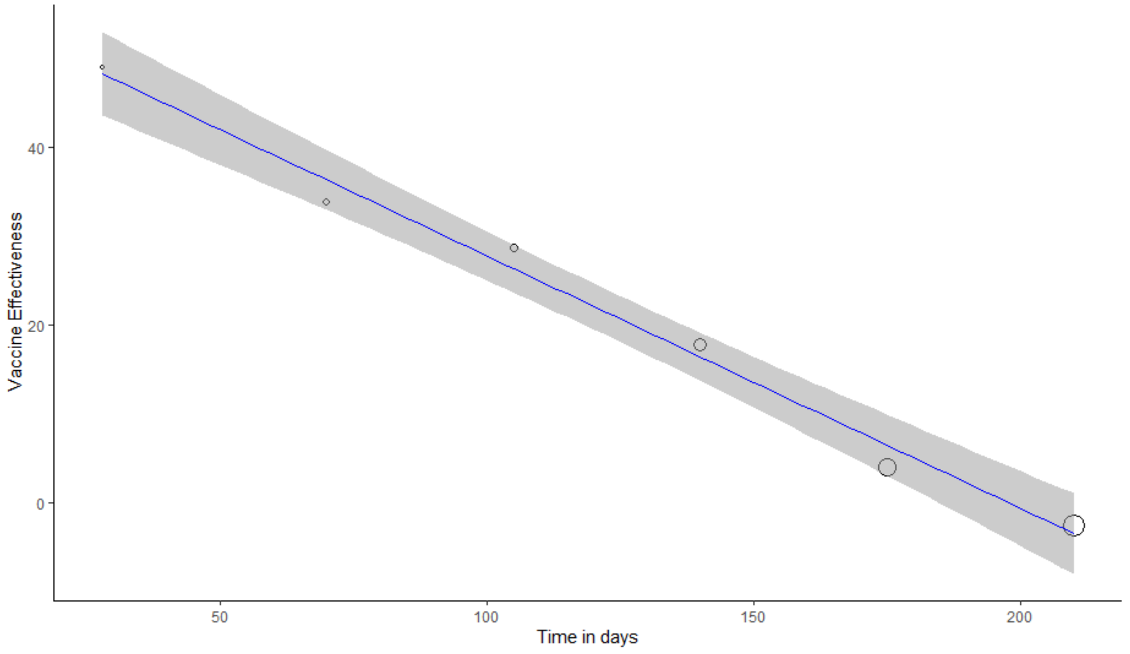 |
| Variable | mixture |
| Results | N/A |
| Reg Plot | N/A |

| Variable | Mixture of mRNA | |
|---|---|---|
| Results | Mixed-Effects Model (k = 15; tau^2 estimator: REML)<br><br>logLik deviance AIC BIC AICc<br>-63.2346 126.4692 132.4692 134.1641 135.1359<br><br>tau^2 (estimated amount of residual heterogeneity): 829.6853 (SE = 371.9081)<br>tau (square root of estimated tau^2 value): 28.8043<br>I^2 (residual heterogeneity / unaccounted variability): 97.99%<br>H^2 (unaccounted variability / sampling variability): 49.87<br>R^2 (amount of heterogeneity accounted for): 0.00%<br><br>Test for Residual Heterogeneity:<br>QE(df = 13) = 477.2669, p-val < .0001<br><br>Test of Moderators (coefficient 2):<br>F(df1 = 1, df2 = 13) = 0.4370, p-val = 0.5201<br><br>Model Results:<br><br>estimate se tval df pval ci.lb ci.ub<br>intrcpt 55.6158 19.2264 2.8927 13 0.0126 14.0797 97.1519 *<br>Days -0.0631 0.0954 -0.6611 13 0.5201 -0.2693 0.1431 | R^2 = 0%<br>VE estimate (%):<br>55.62 (14.08 to 97.15), p:0.0126<br>VE reduction estimate (% per month):<br>-1.893 (-8.079 to 4.293), p:0.5201<br><br>VE (%) = 55.6158 - 0.0631 day |

<table>
<tr><td>Reg Plot</td><td>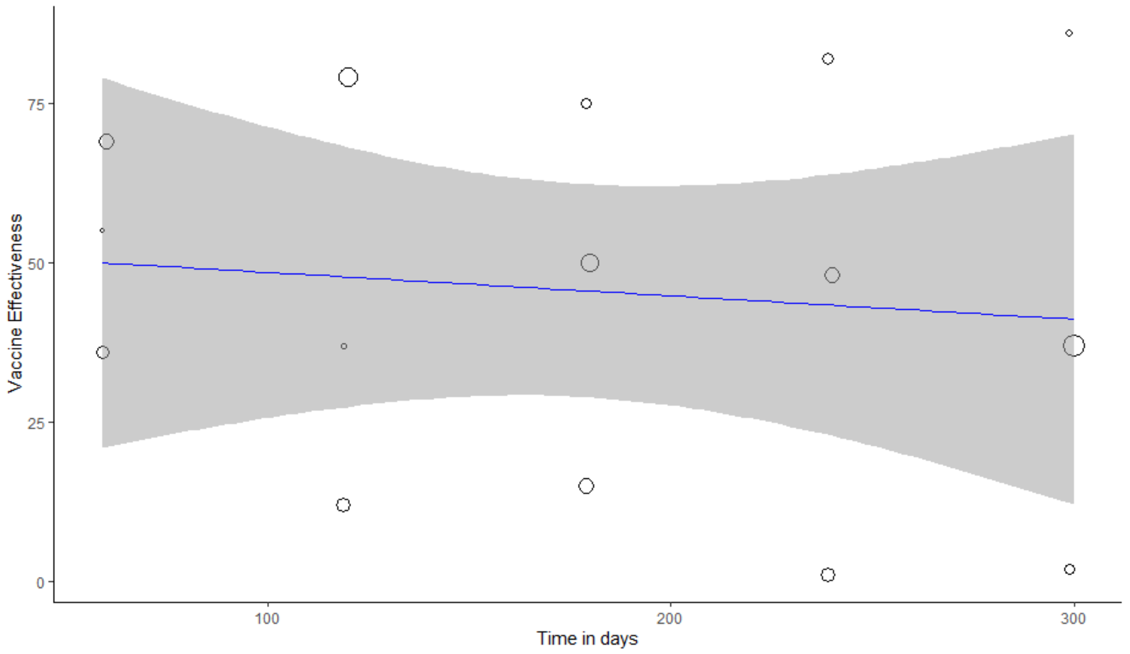
</td><td></td></tr>
<tr><td>Variable</td><td>mRNA-1273</td><td></td></tr>
<tr><td>Results</td><td>Mixed-Effects Model (k = 10; tau^2 estimator: REML)<br><br>logLik deviance AIC BIC AICc<br>-28.3074 56.6147 62.6147 62.8531 68.6147<br><br>tau^2 (estimated amount of residual heterogeneity): 57.4425 (SE = 34.3197)<br>tau (square root of estimated tau^2 value): 7.5791<br>I^2 (residual heterogeneity / unaccounted variability): 92.72%<br>H^2 (unaccounted variability / sampling variability): 13.75<br>R^2 (amount of heterogeneity accounted for): 89.96%<br><br>Test for Residual Heterogeneity:</td><td>R^2 = 89.96%<br>VE estimate (%):<br>76.11 (62.22 to 90), p:<.0001<br>VE reduction estimate (% per month):<br>-9.558 (-12.09 to -7.029), p:<.0001<br><br>VE (%) = 76.1092 - 0.3186 day</td></tr>
</table>

| | |
|---|---|
| | QE(df = 8) = 91.5273, p-val < .0001<br><br>Test of Moderators (coefficient 2):<br>F(df1 = 1, df2 = 8) = 75.8955, p-val < .0001<br><br>Model Results:<br><br>estimate se tval df pval ci.lb ci.ub<br>intrcpt 76.1092 6.0235 12.6354 8 <.0001 62.2191 89.9994 ***<br>Days -0.3186 0.0366 -8.7118 8 <.0001 -0.4030 -0.2343 *** |
| Reg Plot | 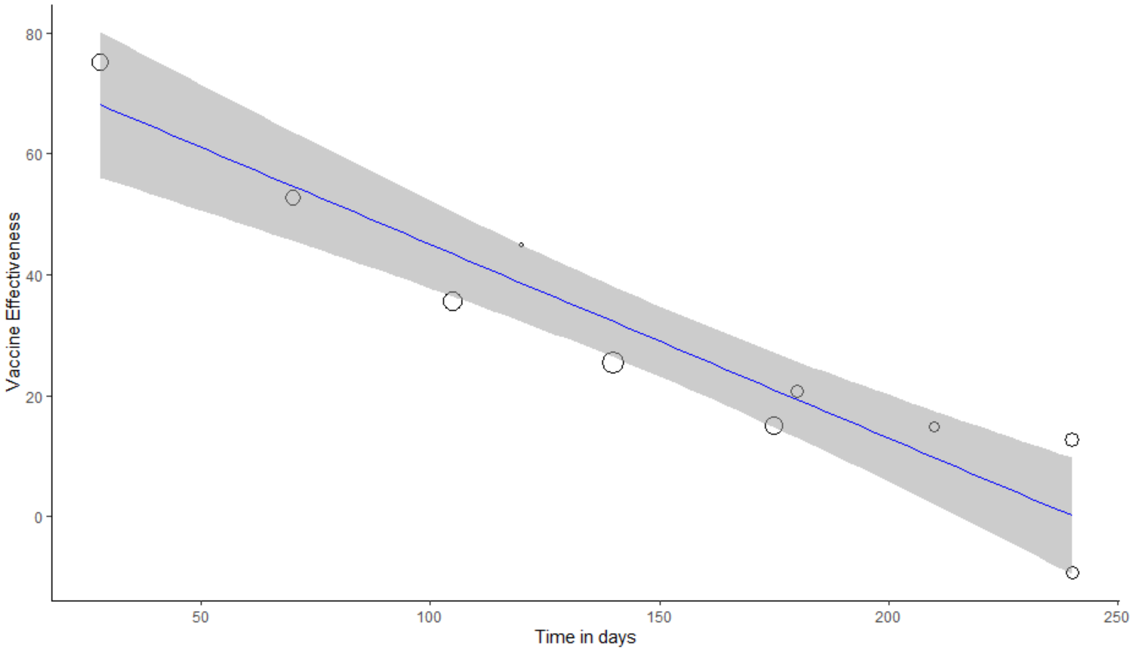 |

Cell is denoted as **green** if meta-regression is performed (i.e., if k > 3). Cell is denoted as **orange** if the result is from a report (k = 1) which stays the same as what has been reported by the original study. Cell is denoted as **red** if the result has not been reported by any studies (k = 0).

#### S.9. Detailed Results of VE Estimates and VE Reduction of Booster Dose

##### Table S12. Detailed results of VE estimates and VE reduction of booster dose for any infection outcome

| Variable | Overall | Report |
|---|---|---|
| Results | Mixed-Effects Model (k = 52; tau^2 estimator: REML)<br><br>logLik deviance AIC BIC AICc<br>-230.1926 460.3852 466.3852 472.1213 466.9070<br><br>tau^2 (estimated amount of residual heterogeneity): 542.6083 (SE = 115.2483)<br>tau (square root of estimated tau^2 value): 23.2940<br>I^2 (residual heterogeneity / unaccounted variability): 99.86%<br>H^2 (unaccounted variability / sampling variability): 710.84<br>R^2 (amount of heterogeneity accounted for): 1.18%<br><br>Test for Residual Heterogeneity:<br>QE(df = 50) = 8970.2627, p-val < .0001<br><br>Test of Moderators (coefficient 2):<br>F(df1 = 1, df2 = 50) = 2.7243, p-val = 0.1051<br><br>Model Results:<br><br>estimate se tval df pval ci.lb ci.ub<br>intrcpt 66.0370 5.3345 12.3792 50 <.0001 55.3223 76.7517 ***<br>Days -0.0769 0.0466 -1.6505 50 0.1051 -0.1706 0.0167 | R^2 = 0%<br>VE estimate (%):<br>66.04 (55.32 to 76.75), p:<.0001<br>VE reduction estimate (% per month):<br>-2.307 (-5.118 to 0.501), p:0.1051<br><br>VE (%) = 66.037 - 0.0769 day |

| | | |
|---|---|---|
| Reg Plot | 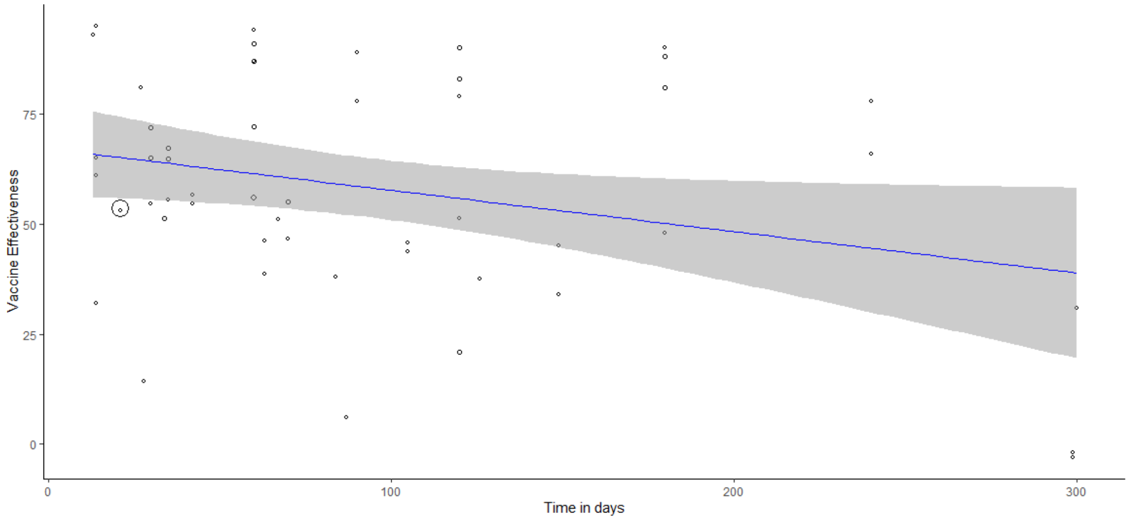<br> | |
| Variable | Ad26.COV2.S | |
| Results | Mixed-Effects Model (k = 5; tau^2 estimator: REML)<br><br>logLik deviance AIC BIC AICc<br>-15.0520 30.1039 36.1039 33.3997 60.1039<br><br>tau^2 (estimated amount of residual heterogeneity): 1254.1640 (SE = 1086.8963)<br>tau (square root of estimated tau^2 value): 35.4142<br>I^2 (residual heterogeneity / unaccounted variability): 97.44%<br>H^2 (unaccounted variability / sampling variability): 39.13<br>R^2 (amount of heterogeneity accounted for): 11.79%<br><br>Test for Residual Heterogeneity:<br>QE(df = 3) = 55.1021, p-val < .0001<br><br>Test of Moderators (coefficient 2):<br>F(df1 = 1, df2 = 3) = 1.5795, p-val = 0.2978 | R^2 = 11.79%<br>VE estimate (%):<br>69.16 (-13.52 to 151.85), p:0.0762<br>VE reduction estimate (% per month):<br>-22.359 (-78.981 to 34.26), p:0.2978<br><br>VE (%) = 69.1648 - 0.7453 day |

| | | |
|---|---|---|
| | Model Results:<br><br>estimate se tval df pval ci.lb ci.ub<br>intrcpt 69.1648 25.9815 2.6621 3 0.0762 -13.5200 151.8496 .<br>Days -0.7453 0.5931 -1.2568 3 0.2978 -2.6327 1.1420 | |
| Reg Plot | 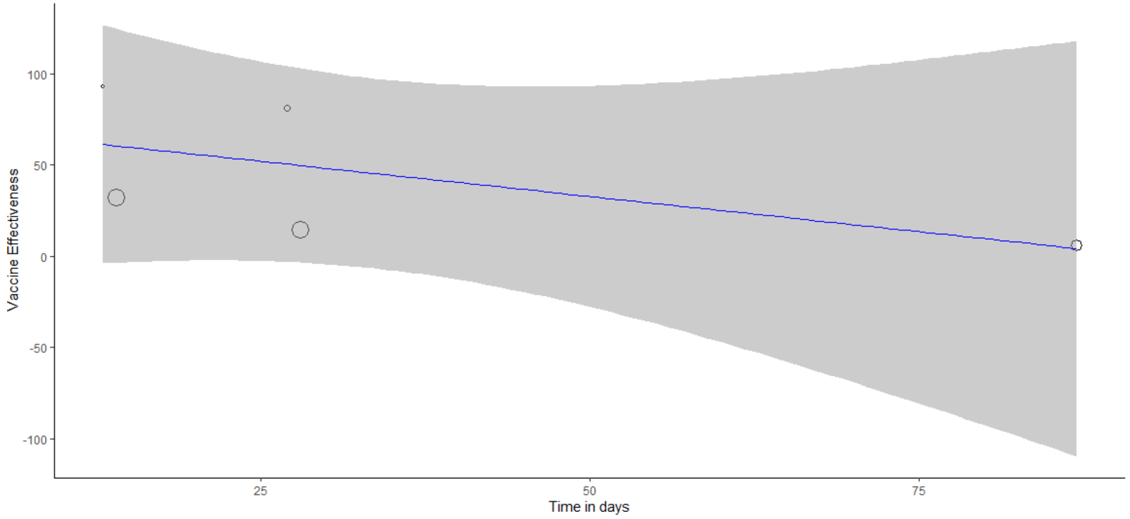 | |
| Variable | BNT162b2 | |
| Results | Mixed-Effects Model (k = 20; tau^2 estimator: REML)<br><br>logLik deviance AIC BIC AICc<br>-78.0335 156.0670 162.0670 164.7381 163.7812<br><br>tau^2 (estimated amount of residual heterogeneity): 282.9738 (SE = 107.4060)<br>tau (square root of estimated tau^2 value): 16.8218<br>I^2 (residual heterogeneity / unaccounted variability): 99.63%<br>H^2 (unaccounted variability / sampling variability): 270.53<br>R^2 (amount of heterogeneity accounted for): 36.53%<br><br>Test for Residual Heterogeneity: | R^2 = 36.53%<br>VE estimate (%):<br>69.79 (54.94 to 84.65), p:<.0001<br>VE reduction estimate (% per month):<br>-5.442 (-8.988 to -1.893), p:0.0047<br><br>VE (%) = 69.7933 - 0.1814 day |

| | | |
|---|---|---|
| | QE(df = 18) = 1907.8760, p-val < .0001<br><br>Test of Moderators (coefficient 2):<br>F(df1 = 1, df2 = 18) = 10.3822, p-val = 0.0047<br><br>Model Results:<br><br>estimate se tval df pval ci.lb ci.ub<br>intrcpt 69.7933 7.0722 9.8687 18 <.0001 54.9352 84.6513 ***<br>Days -0.1814 0.0563 -3.2221 18 0.0047 -0.2996 -0.0631 ** | |
| Reg Plot | 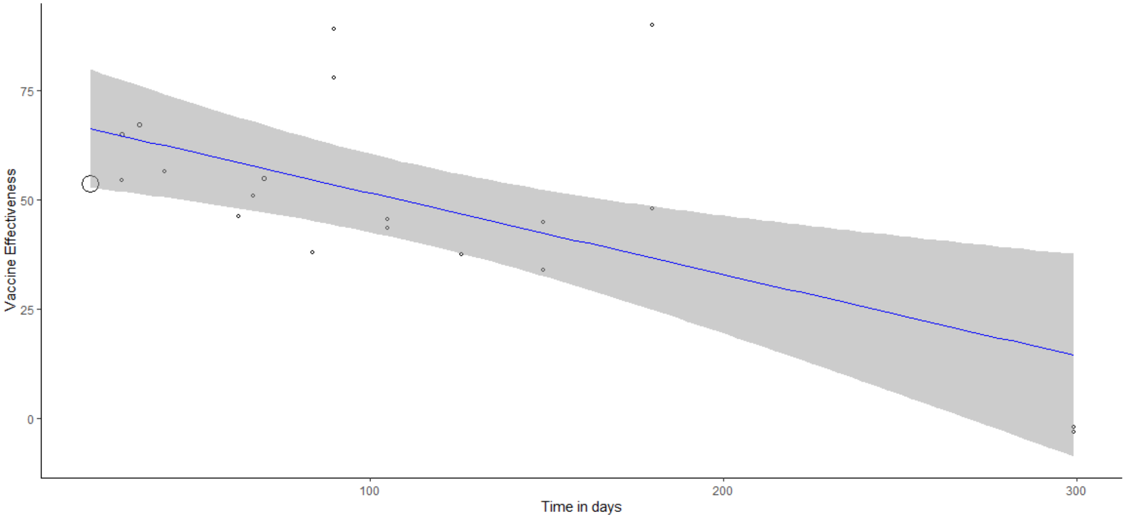 | |
| Variable | ChAdOx1 | |
| Results | Number of studies combined: k = 2<br><br>VE 95%-CI t p-value<br>Random effects model 51.4480 [-4.9677; 107.8637] 11.59 0.0548<br><br>Quantifying heterogeneity:<br>tau^2 = 9.6042; tau = 3.0991; I^2 = 24.2%; H = 1.15 | I^2 = 24.2%<br>VE estimate (%):<br>51.4480 (-4.9677 to 107.8637), p: 0.0548<br>VE reduction estimate (% per month): N/A |

| | | |
|---|---|---|
| | Test of heterogeneity:<br>Q d.f. p-value<br>1.32 1 0.2506 | |
| Reg Plot | N/A | |
| Variable | Mixture | |
| Results | Mixed-Effects Model (k = 7; tau^2 estimator: REML)<br><br>logLik deviance AIC BIC AICc<br>-23.6424 47.2849 53.2849 52.1132 77.2849<br><br>tau^2 (estimated amount of residual heterogeneity): 744.5094 (SE = 474.1033)<br>tau (square root of estimated tau^2 value): 27.2857<br>I^2 (residual heterogeneity / unaccounted variability): 99.94%<br>H^2 (unaccounted variability / sampling variability): 1581.11<br>R^2 (amount of heterogeneity accounted for): 0.00%<br><br>Test for Residual Heterogeneity:<br>QE(df = 5) = 3493.4528, p-val < .0001<br><br>Test of Moderators (coefficient 2):<br>F(df1 = 1, df2 = 5) = 0.0968, p-val = 0.7683<br><br>Model Results:<br><br>estimate se tval df pval ci.lb ci.ub<br>intrcpt 58.2918 28.7750 2.0258 5 0.0986 -15.6768 132.2603 .<br>Days 0.0921 0.2960 0.3111 5 0.7683 -0.6689 0.8531 | R^2 = 0%<br>VE estimate (%):<br>58.29 (-15.68 to 132.26), p:0.0986<br>VE reduction estimate (% per month):<br>2.763 (-20.067 to 25.593), p:0.7683<br><br>VE (%) = 58.2918 + 0.0921 day |

| | | |
|---|---|---|
| Reg Plot | 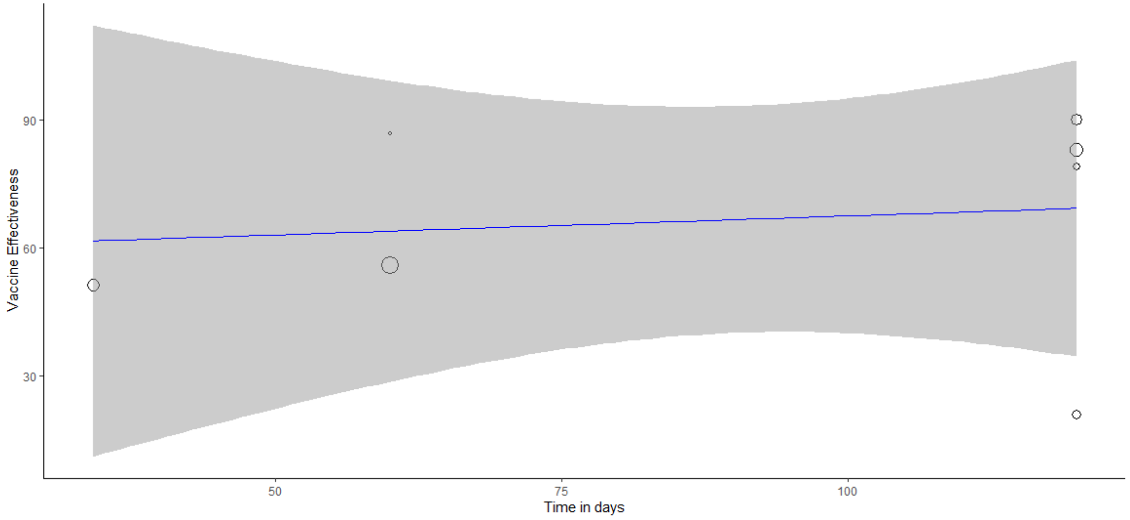 | |
| Variable | Mixture of mRNA | |
| Results | Mixed-Effects Model (k = 10; tau^2 estimator: REML)<br><br>logLik deviance AIC BIC AICc<br>-32.5356 65.0712 71.0712 71.3095 77.0712<br><br>tau^2 (estimated amount of residual heterogeneity): 148.4871 (SE = 81.8958)<br>tau (square root of estimated tau^2 value): 12.1855<br>I^2 (residual heterogeneity / unaccounted variability): 98.22%<br>H^2 (unaccounted variability / sampling variability): 56.20<br>R^2 (amount of heterogeneity accounted for): 0.00%<br><br>Test for Residual Heterogeneity:<br>QE(df = 8) = 215.9954, p-val < .0001<br><br>Test of Moderators (coefficient 2):<br>F(df1 = 1, df2 = 8) = 1.3832, p-val = 0.2734<br><br>Model Results: | R^2 = 0%<br>VE estimate (%):<br>88.23 (72.01 to 104.46), p:<.0001<br>VE reduction estimate (% per month):<br>-1.68 (-4.971 to 1.614), p:0.2734<br><br>VE (%) = 88.2341 - 0.056 day |

| | | |
|---|---|---|
| | estimate se tval df pval ci.lb ci.ub<br>intrcpt 88.2341 7.0370 12.5386 8 <.0001 72.0068 104.4614 ***<br>Days -0.0560 0.0476 -1.1761 8 0.2734 -0.1657 0.0538 | |
| Reg Plot | 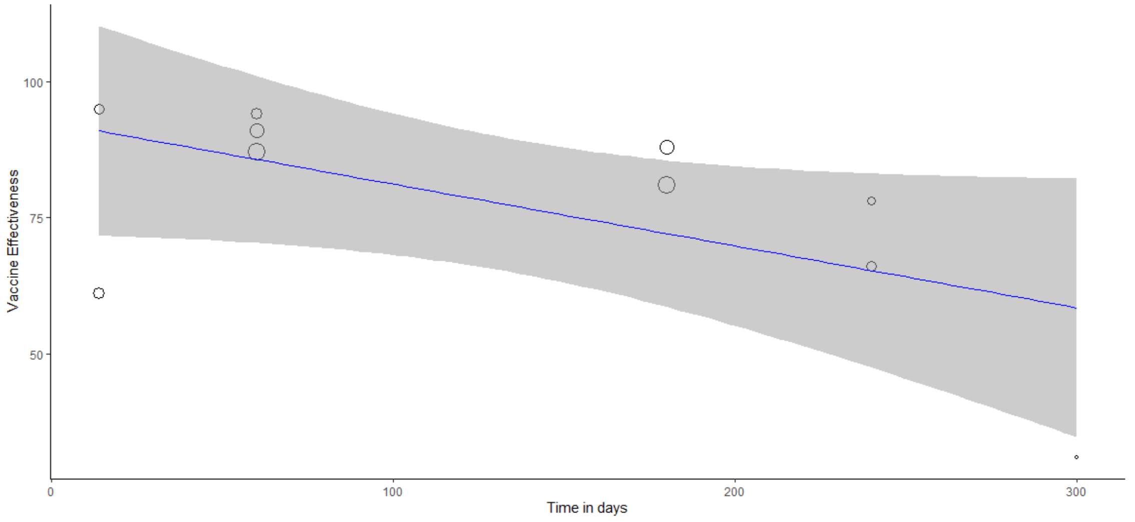 | |
| Variable | mRNA-1273 | |
| Results | Mixed-Effects Model (k = 8; tau^2 estimator: REML)<br><br>logLik deviance AIC BIC AICc<br>-22.9728 45.9455 51.9455 51.3208 63.9455<br><br>tau^2 (estimated amount of residual heterogeneity): 94.7994 (SE = 65.0811)<br>tau (square root of estimated tau^2 value): 9.7365<br>I^2 (residual heterogeneity / unaccounted variability): 95.56%<br>H^2 (unaccounted variability / sampling variability): 22.54<br>R^2 (amount of heterogeneity accounted for): 1.43%<br><br>Test for Residual Heterogeneity:<br>QE(df = 6) = 78.7042, p-val < .0001 | R^2 = 1.43%<br>VE estimate (%):<br>66.54 (49.43 to 83.65), p:<.0001<br>VE reduction estimate (% per month):<br>-3.717 (-12.579 to 5.145), p:0.3444<br><br>VE (%) = 66.5414 - 0.1239 day |

| | |
|---|---|
| | Test of Moderators (coefficient 2):<br>F(df1 = 1, df2 = 6) = 1.0529, p-val = 0.3444<br><br>Model Results:<br><br>estimate se tval df pval ci.lb ci.ub<br>intrcpt 66.5414 6.9927 9.5158 6 <.0001 49.4308 83.6519 ***<br>Days -0.1239 0.1207 -1.0261 6 0.3444 -0.4193 0.1715 |
| Reg Plot | 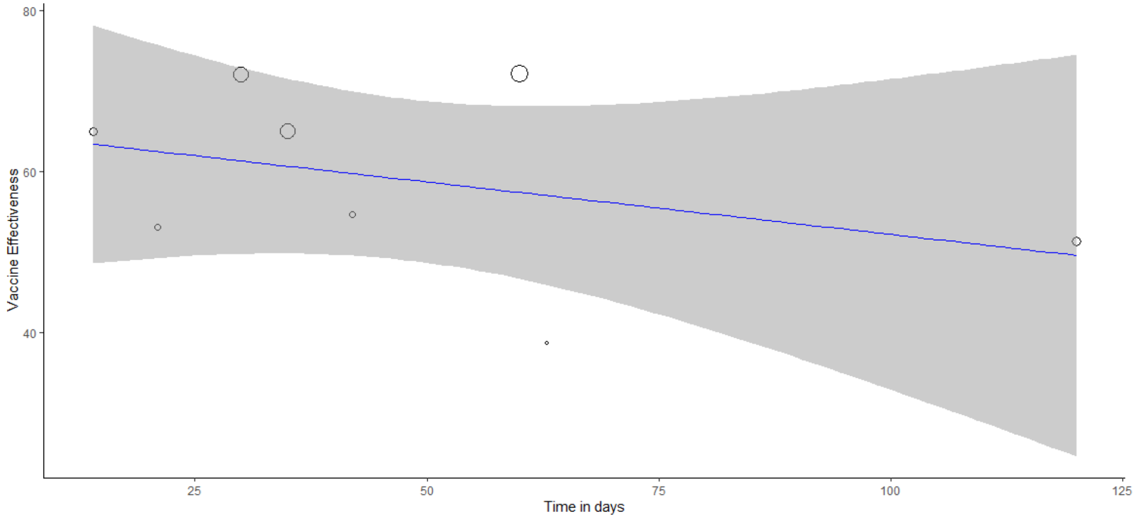 |

Cell is denoted as **green** if meta-regression is performed (i.e., if k > 3).

Table S13. Detailed results of VE estimates and VE reduction of booster dose for severe infection outcome

| Variable | overall | |
|---|---|---|
| Results | Mixed-Effects Model (k = 12; tau^2 estimator: REML)<br><br>logLik deviance AIC BIC AICc<br>-30.9074 61.8147 67.8147 68.7225 71.8147<br><br>tau^2 (estimated amount of residual heterogeneity): 13.4208 (SE = 9.8845)<br>tau (square root of estimated tau^2 value): 3.6634<br>I^2 (residual heterogeneity / unaccounted variability): 75.90%<br>H^2 (unaccounted variability / sampling variability): 4.15<br>R^2 (amount of heterogeneity accounted for): 38.93%<br><br>Test for Residual Heterogeneity:<br>QE(df = 10) = 32.6589, p-val = 0.0003<br><br>Test of Moderators (coefficient 2):<br>F(df1 = 1, df2 = 10) = 6.3464, p-val = 0.0304<br><br>Model Results:<br><br>estimate se tval df pval ci.lb ci.ub<br>intrcpt 93.9648 2.5418 36.9678 10 <.0001 88.3013 99.6283 ***<br>Days -0.0539 0.0214 -2.5192 10 0.0304 -0.1015 -0.0062 * | R^2 = 38.93%<br>VE estimate (%):<br>93.96 (88.3 to 99.63), p<.0001<br>Decrease (% per month):<br>-1.617 (-3.045 to -0.186), p: <.0001<br>VE (%) = 93.9648 - 0.0539 day |

<table>
<tr><td>Reg Plot</td><td>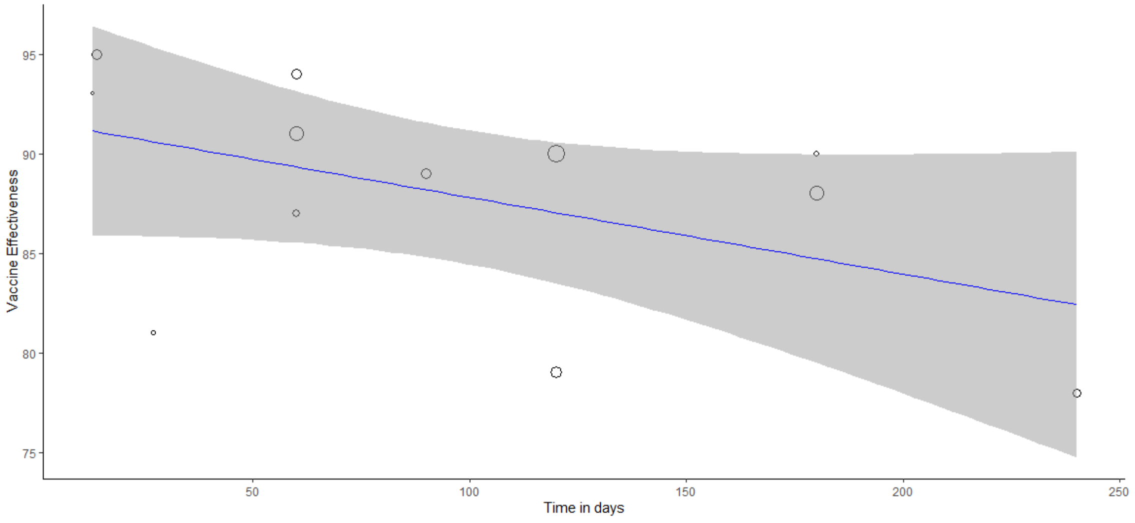
</td><td></td></tr>
<tr><td>Variable</td><td>Ad26.COV2.S</td><td></td></tr>
<tr><td>Results</td><td>Number of studies combined: k = 2<br><br>VE 95%-CI t p-value<br>Random effects model 86.0069 [10.8212; 161.1926] 14.53 0.0437<br><br>Quantifying heterogeneity:<br>tau^2 = 0; tau = 0; I^2 = 0.0%; H = 1.00<br><br>Test of heterogeneity:<br>Q d.f. p-value<br>0.48 1 0.4898</td><td>I^2 = 0%<br>VE estimate (%):<br>86.0069 (10.8212 to 161.1926), p: 0.0437<br>VE reduction estimate (% per month): N/A</td></tr>
<tr><td>Reg Plot</td><td>N/A</td><td></td></tr>
<tr><td>Variable</td><td>BNT162b2</td><td></td></tr>
<tr><td>Results</td><td>Number of studies combined: k = 2<br><br>VE 95%-CI t p-value</td><td>I^2 = 0%<br>VE estimate (%):</td></tr>
</table>

| | | |
|---|---|---|
| | Random effects model 89.0460 [86.3850; 91.7069] 425.20 0.0015<br><br>Quantifying heterogeneity:<br>tau^2 = 0; tau = 0; I^2 = 0.0%; H = 1.00<br><br>Test of heterogeneity:<br>Q d.f. p-value<br>0.01 1 0.9256 | 89.0460 (86.3850 to 91.7069), p:0.0015<br>VE reduction estimate (% per month): N/A |
| Reg Plot | N/A | |
| Variable | ChAdOx1 nCov-19 | |
| Results | N/A | |
| Reg Plot | N/A | |
| Variable | mixture | |
| Results | Mixed-Effects Model (k = 3; tau^2 estimator: REML)<br><br>logLik deviance AIC BIC AICc<br>-3.4703 6.9405 12.9405 6.9405 36.9405<br><br>tau^2 (estimated amount of residual heterogeneity): 58.1247 (SE = 85.5599)<br>tau (square root of estimated tau^2 value): 7.6240<br>I^2 (residual heterogeneity / unaccounted variability): 96.07%<br>H^2 (unaccounted variability / sampling variability): 25.47<br>R^2 (amount of heterogeneity accounted for): 0.00%<br><br>Test for Residual Heterogeneity:<br>QE(df = 1) = 25.4703, p-val < .0001<br><br>Test of Moderators (coefficient 2):<br>F(df1 = 1, df2 = 1) = 0.0467, p-val = 0.8645<br><br>Model Results:<br><br>estimate se tval df pval ci.lb ci.ub | R^2 = 0%<br>VE estimate (%):<br>89.34 (-157.54 to 336.22), p:0.1363<br>VE reduction estimate (% per month):<br>-1.17 (-69.903 to 67.566), p:0.8645<br><br>VE (%) = 89.3373 - 0.039 day |

| | | |
|---|---|---|
| | intrcpt 89.3373 19.4298 4.5979 1 0.1363 -157.5421 336.2167<br>Days -0.0390 0.1803 -0.2160 1 0.8645 -2.3301 2.2522 | |
| Reg Plot | 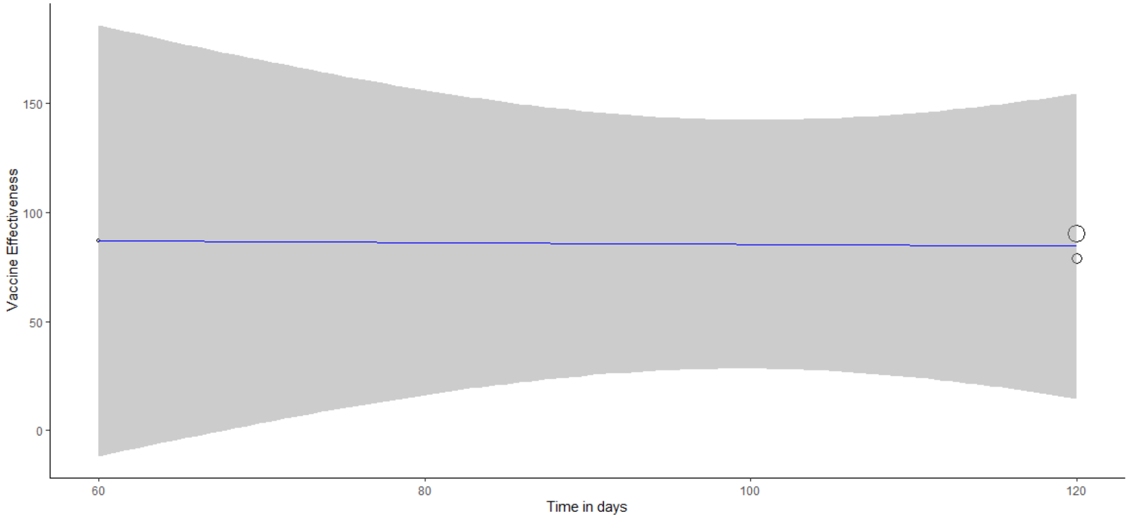<br> | |
| Variable | Mixture of mRNA | |
| Results | Mixed-Effects Model (k = 5; tau^2 estimator: REML)<br><br>logLik deviance AIC BIC AICc<br>-7.1849 14.3698 20.3698 17.6656 44.3698<br><br>tau^2 (estimated amount of residual heterogeneity): 1.7452 (SE = 5.1685)<br>tau (square root of estimated tau^2 value): 1.3210<br>I^2 (residual heterogeneity / unaccounted variability): 27.20%<br>H^2 (unaccounted variability / sampling variability): 1.37<br>R^2 (amount of heterogeneity accounted for): 91.43%<br><br>Test for Residual Heterogeneity:<br>QE(df = 3) = 4.1131, p-val = 0.2495<br><br>Test of Moderators (coefficient 2):<br>F(df1 = 1, df2 = 3) = 9.7430, p-val = 0.0524 | R^2 = 91.43%<br>VE estimate (%):<br>95.26 (89.19 to 101.34), p:<.0001<br>VE reduction estimate (% per month):<br>-1.434 (-2.895 to 0.027), p:0.0524<br><br>VE (%) = 95.2629 - 0.0478 day |

| | | |
|---|---|---|
| | Model Results:<br><br>estimate se tval df pval ci.lb ci.ub<br>intrcpt 95.2629 1.9087 49.9093 3 <.0001 89.1885 101.3373 ***<br>Days -0.0478 0.0153 -3.1214 3 0.0524 -0.0965 0.0009 . | |
| Reg Plot | 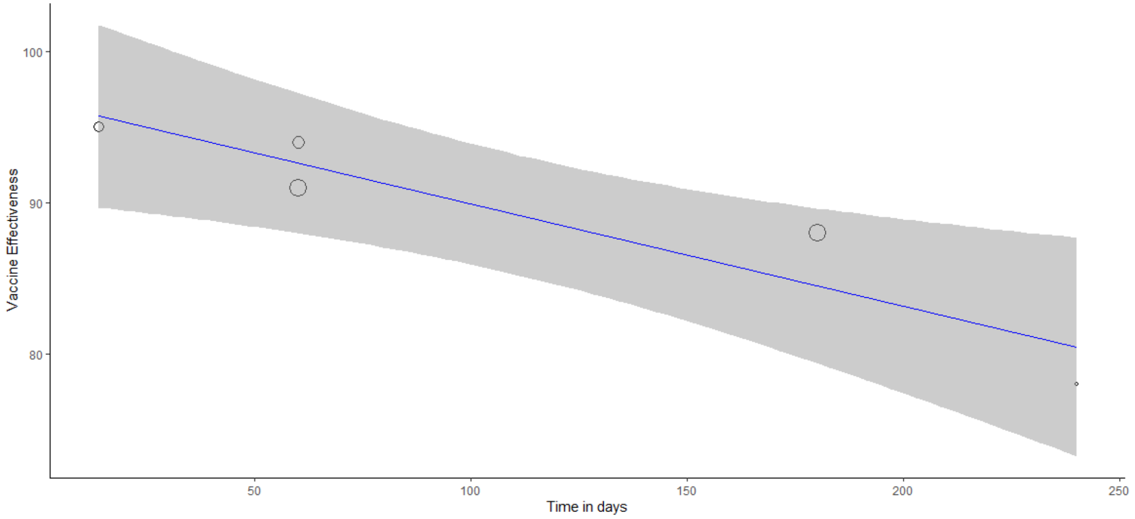 | |
| Variable | mRNA-1273 | |
| Results | VE: 99.2% (76.3% to 100%) | VE estimate (%): 99.2% (76.3% to 100%)<br>VE reduction estimate (% per month): N/A |
| Reg Plot | N/A | |

Cell is denoted as **green** if meta-regression is performed (i.e., if k > 3). Cell is denoted as **yellow** if the result is calculated from meta-analysis of two report (k = 2). Cell is denoted as **orange** if the result is from a report (k = 1) which stays the same as what has been reported by the original study. Cell is denoted as **red** if the result has not been reported by any studies (k = 0).

Table S14. Detailed results of VE estimates and VE reduction of booster dose for symptomatic infection outcome

| Variable | overall | |
|---|---|---|
| Results | Mixed-Effects Model (k = 32; tau^2 estimator: REML)<br><br>logLik deviance AIC BIC AICc<br>-129.3988 258.7975 264.7975 269.0011 265.7206<br><br>tau^2 (estimated amount of residual heterogeneity): 297.8935 (SE = 84.8558)<br>tau (square root of estimated tau^2 value): 17.2596<br>I^2 (residual heterogeneity / unaccounted variability): 99.81%<br>H^2 (unaccounted variability / sampling variability): 517.13<br>R^2 (amount of heterogeneity accounted for): 15.68%<br><br>Test for Residual Heterogeneity:<br>QE(df = 30) = 6592.3798, p-val < .0001<br><br>Test of Moderators (coefficient 2):<br>F(df1 = 1, df2 = 30) = 8.9309, p-val = 0.0055<br><br>Model Results:<br><br>estimate se tval df pval ci.lb ci.ub<br>intrcpt 65.0327 5.1151 12.7138 30 <.0001 54.5863 75.4791 ***<br>Days -0.1286 0.0430 -2.9885 30 0.0055 -0.2164 -0.0407 ** | R^2 = 15.68%<br>VE estimate (%):<br>65.03 (54.59 to 75.48), p:<.0001<br>VE reduction estimate (% per month):<br>-3.858 (-6.492 to -1.221), p:0.0055<br><br>VE (%) = 65.0327 - 0.1286 day |

| | | |
|---|---|---|
| Reg Plot | 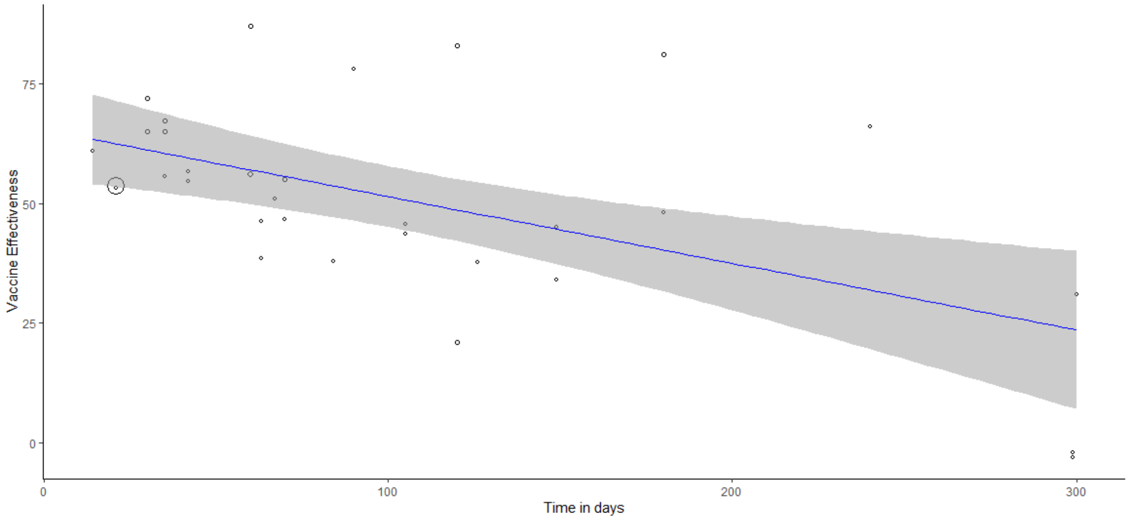 | |
| Variable | Ad26.COV2.S | |
| Results | N/A | |
| Reg Plot | N/A | |
| Variable | BNT162b2 | |
| Results | Mixed-Effects Model (k = 17; tau^2 estimator: REML)<br><br>logLik deviance AIC BIC AICc<br>-57.6048 115.2096 121.2096 123.3337 123.3914<br><br>tau^2 (estimated amount of residual heterogeneity): 103.7006 (SE = 47.6416)<br>tau (square root of estimated tau^2 value): 10.1834<br>I^2 (residual heterogeneity / unaccounted variability): 99.13%<br>H^2 (unaccounted variability / sampling variability): 115.43<br>R^2 (amount of heterogeneity accounted for): 68.20%<br><br>Test for Residual Heterogeneity:<br>QE(df = 15) = 1755.9344, p-val < .0001 | R^2 = 68.2%<br>VE estimate (%):<br>68.79 (59.21 to 78.38), p:<.0001<br>VE reduction estimate (% per month):<br>-6.423 (-8.898 to -3.948), p:<.0001<br><br>VE (%) = 68.7937 - 0.2141 day |

| | | |
|---|---|---|
| | Test of Moderators (coefficient 2):<br>F(df1 = 1, df2 = 15) = 30.6190, p-val < .0001<br><br>Model Results:<br><br>estimate se tval df pval ci.lb ci.ub<br>intrcpt 68.7937 4.4963 15.3002 15 <.0001 59.2101 78.3773 ***<br>Days -0.2141 0.0387 -5.5334 15 <.0001 -0.2966 -0.1316 *** | |
| Reg Plot | 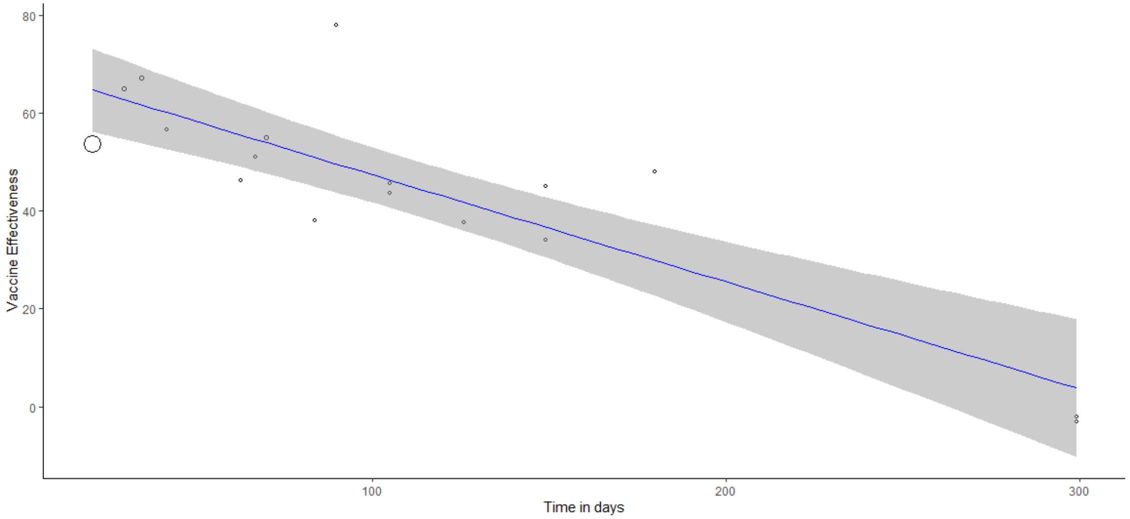  | |
| Variable | ChAdOx1 nCov-19 | |
| Results | Number of studies combined: k = 2<br><br>VE 95%-CI t p-value<br>Random effects model 51.4480 [-4.9677; 107.8637] 11.59 0.0548<br><br>Quantifying heterogeneity:<br>tau^2 = 9.6042; tau = 3.0991; I^2 = 24.2%; H = 1.15 | I^2 = 24.2%<br>VE estimate (%):<br>51.4480 (-4.9677to 107.8637),<br>p:0.0548<br>VE reduction estimate (% per month):<br>N/A |

| | | |
|---|---|---|
| | Test of heterogeneity:<br>Q d.f. p-value<br>1.32 1 0.2506 | |
| Reg Plot | N/A | |
| Variable | mixture | |
| Results | Mixed-Effects Model (k = 3; tau^2 estimator: REML)<br><br>logLik deviance AIC BIC AICc<br>-5.1995 10.3990 16.3990 10.3990 40.3990<br><br>tau^2 (estimated amount of residual heterogeneity): 1921.3492 (SE = 2718.1185)<br>tau (square root of estimated tau^2 value): 43.8332<br>I^2 (residual heterogeneity / unaccounted variability): 99.97%<br>H^2 (unaccounted variability / sampling variability): 2953.42<br>R^2 (amount of heterogeneity accounted for): 0.00%<br><br>Test for Residual Heterogeneity:<br>QE(df = 1) = 2953.4221, p-val < .0001<br><br>Test of Moderators (coefficient 2):<br>F(df1 = 1, df2 = 1) = 0.0055, p-val = 0.9527<br><br>Model Results:<br><br>estimate se tval df pval ci.lb ci.ub<br>intrcpt 59.9937 92.9874 0.6452 1 0.6352 -1121.5233 1241.5107<br>Days -0.0666 0.8948 -0.0744 1 0.9527 -11.4361 11.3030 | R^2 = 0%<br>VE estimate (%):<br>59.99 (-1121.52 to 1241.51), p:0.6352<br>VE reduction estimate (% per month):<br>-1.998 (-343.083 to 339.09), p:0.9527<br><br>VE (%) = 59.9937 - 0.0666 day |

| | | |
|---|---|---|
| Reg Plot | 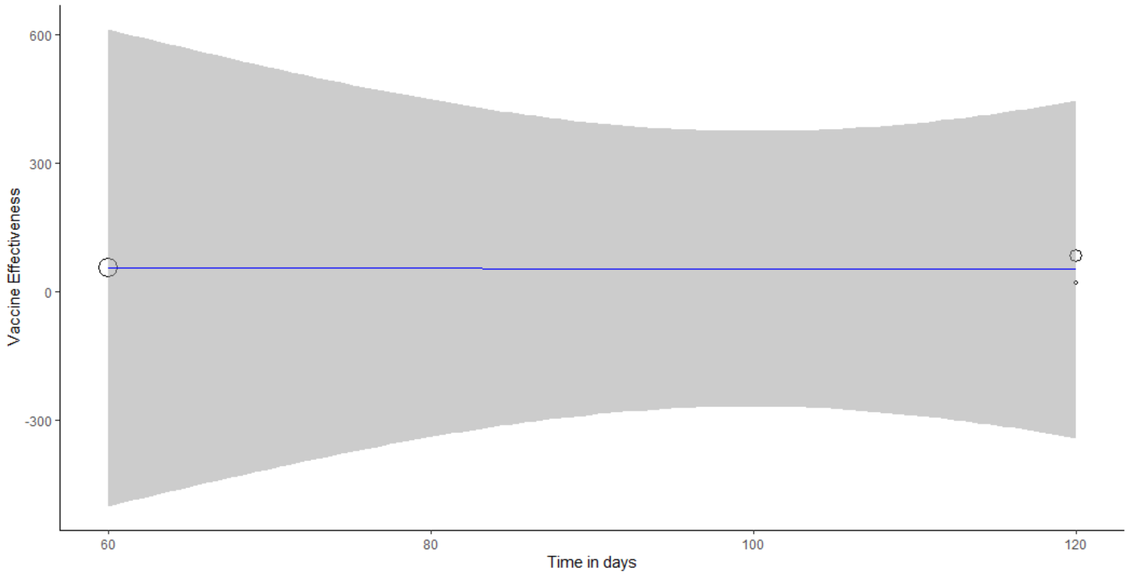 | |
| Variable | Mixture of mRNA | |
| Results | Mixed-Effects Model (k = 5; tau^2 estimator: REML)<br><br>logLik deviance AIC BIC AICc<br>-13.0663 26.1326 32.1326 29.4284 56.1326<br><br>tau^2 (estimated amount of residual heterogeneity): 236.0327 (SE = 231.3246)<br>tau (square root of estimated tau^2 value): 15.3634<br>I^2 (residual heterogeneity / unaccounted variability): 98.40%<br>H^2 (unaccounted variability / sampling variability): 62.33<br>R^2 (amount of heterogeneity accounted for): 0.00%<br><br>Test for Residual Heterogeneity:<br>QE(df = 3) = 147.6486, p-val < .0001<br><br>Test of Moderators (coefficient 2):<br>F(df1 = 1, df2 = 3) = 0.2119, p-val = 0.6766 | R^2 = 0%<br>VE estimate (%):<br>76.66 (34.59 to 118.73), p:0.0102<br>VE reduction estimate (% per month):<br>-1.119 (-8.862 to 6.621), p:0.6766<br><br>VE (%) = 76.6635 - 0.0373 day |

| | | |
|---|---|---|
| | Model Results:<br><br>estimate se tval df pval ci.lb ci.ub<br>intrcpt 76.6635 13.2197 5.7992 3 0.0102 34.5924 118.7346 *<br>Days -0.0373 0.0811 -0.4604 3 0.6766 -0.2954 0.2207 | |
| Reg Plot | 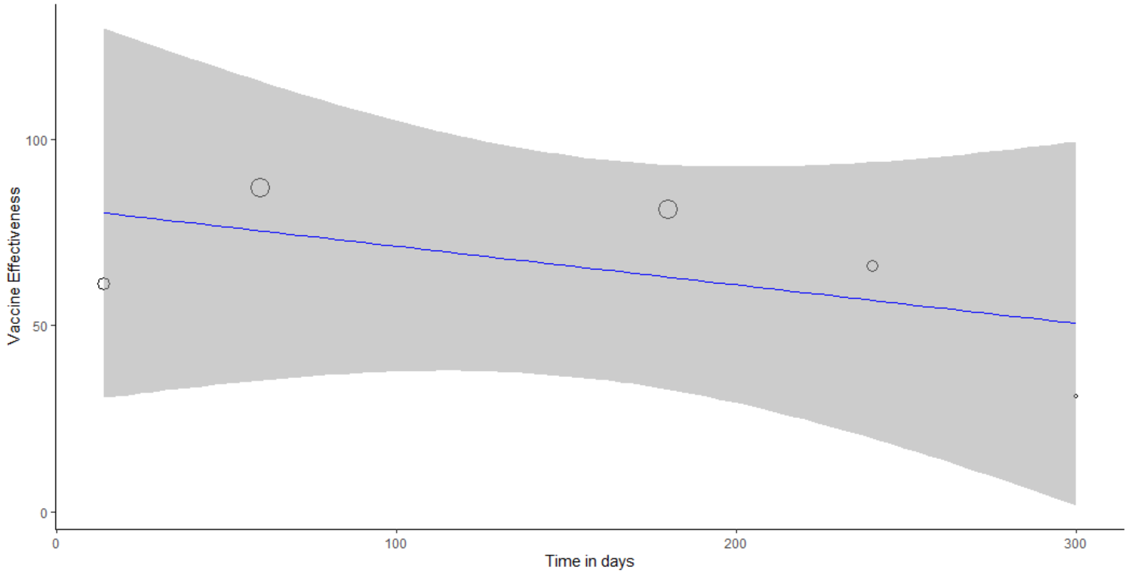 | |
| Variable | mRNA-1273 | |
| Results | Mixed-Effects Model (k = 5; tau^2 estimator: REML)<br><br>logLik deviance AIC BIC AICc<br>-11.3853 22.7707 28.7707 26.0665 52.7707<br><br>tau^2 (estimated amount of residual heterogeneity): 95.7588 (SE = 92.1843)<br>tau (square root of estimated tau^2 value): 9.7856<br>I^2 (residual heterogeneity / unaccounted variability): 94.39%<br>H^2 (unaccounted variability / sampling variability): 17.84<br>R^2 (amount of heterogeneity accounted for): 20.38% | R^2 = 20.38%<br>VE estimate (%):<br>78.44 (30.98 to 125.9), p:0.0134<br>VE reduction estimate (% per month):<br>-16.152 (-53.139 to 20.835), p:0.2588<br><br>VE (%) = 78.4407 - 0.5384 day |

| | |
|---|---|
| | Test for Residual Heterogeneity:<br>QE(df = 3) = 22.5681, p-val < .0001<br><br>Test of Moderators (coefficient 2):<br>F(df1 = 1, df2 = 3) = 1.9314, p-val = 0.2588<br><br>Model Results:<br><br>estimate se tval df pval ci.lb ci.ub<br>intrcpt 78.4407 14.9141 5.2595 3 0.0134 30.9773 125.9041 *<br>Days -0.5384 0.3874 -1.3898 3 0.2588 -1.7713 0.6945 |
| Reg Plot | 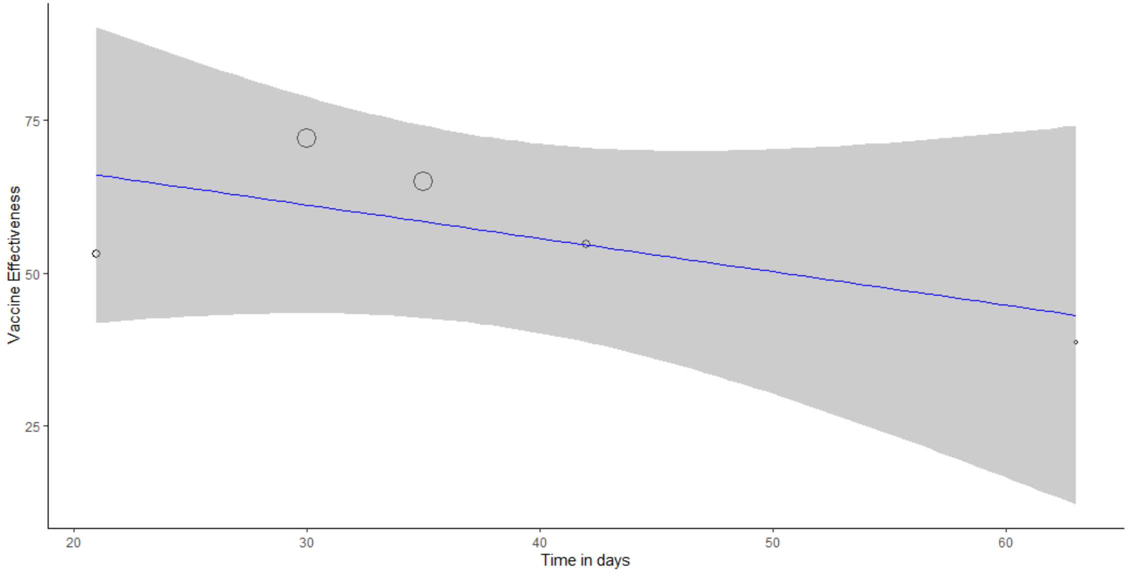 |

Cell is denoted as **green** if meta-regression is performed (i.e., if k > 3). Cell is denoted as **yellow** if the result is calculated from meta-analysis of two report (k = 2). Cell is denoted as **red** if the result has not been reported by any studies (k = 0).

#### S.10. Detailed Results of Meta-analyses for the 'Within 3 Months' Model

##### Table S15. Detailed Results of meta-analyses for within 3 months model

| Variable | k | VED (95%CI) | p-value of VED | Tau2 | I2 (95%CI) | p-value of I2 | Chi^2 | df | p-value of Egger's test |
|---|---|---|---|---|---|---|---|---|---|
| **Any Infection** | | | | | | | | | |
| The Overall Result | 22 | 22% (15% to 29%) | <0,0001 | 0.0207 | 98.3% (97.9% to 98.6%) | <0,0001 | 1214.69 | 21 | 0.203724 |
| *Study Design Subgroup:* | | | | | | | | | |
| Cohort | 6 | 35% (14% to 57%) | <0,0001 | 0.0678 | 99.6% (99.4% to 99.7%) | <0,0001 | 1148.18 | 5 | 0.245683 |
| Case-control | 16 | 17% (13% to 21%) | <0,0001 | 0.0041 | 76.1% (61.4% to 85.3%) | <0,0001 | 62.88 | 15 | 0.811881 |
| *Vaccine Subgroup:* | | | | | | | | | |
| Ad26.COV2 | 0 | N/A | N/A | N/A | N/A | N/A | N/A | N/A | N/A |
| BNT162b2 | 8 | 25% (8% to 41%) | <0,0001 | 0.0485 | 98.9% (98.5% to 99.1%) | <0,0001 | 616.65 | 7 | 0.856062 |
| ChAdOx1 nCov-19 | 1 | 16% (2% to 30%) | N/A | N/A | N/A | N/A | N/A | N/A | N/A |
| Mixture | 2 | 27% (-1% to 55%) | <0.0001 | 0.0407 | 96.9% (91.8% to 98.8%) | <0.0001 | 32.17 | 1 | N/A |
| Mixture of mRNA | 7 | 15% (6% to 24%) | <0.0001 | 0.0105 | 79.9% (59% to 90.2%) | <0.0001 | 29.88 | 6 | 0.845442 |
| mRNA-1273 | 4 | 25% (4% to 47%) | <0,0001 | 0.0423 | 96.8% (94.3% to 98.2%) | <0,0001 | 94.28 | 3 | 0.889649 |
| **Severe Infection** | | | | | | | | | |
| The Overall Result | 7 | 20% (8% to 32%) | <0,0001 | 0.0193 | 81.1% (61.9% to 90.6%) | <0,0001 | 31.8 | 6 | 0.919516 |
| *Study Design Subgroup:* | | | | | | | | | |
| Cohort | 2 | 34% (16% to 52%) | 0.0525 | 0.0126 | 73.4% (0% to 94%) | 0.0525 | 3.76 | 1 | N/A |
| Case-control | 5 | 13% (5% to 20%) | 0.2038 | 0.0024 | 32.6% (0% to 74.4%) | 0.2038 | 5.94 | 4 | 0.477389 |
| *Vaccine Subgroup:* | | | | | | | | | |
| Ad26.COV2 | 0 | N/A | N/A | N/A | N/A | N/A | N/A | N/A | N/A |
| BNT162b2 | 2 | 22% (9% to 35%) | 0.7444 | 0 | 0% (0% to 0%) | 0.7444 | 0.11 | 1 | N/A |
| ChAdOx1 nCov-19 | 0 | N/A | N/A | N/A | N/A | N/A | N/A | N/A | N/A |
| Mixture | 1 | 42% (32% to 52%) | N/A | N/A | N/A | N/A | N/A | N/A | N/A |
| Mixture of mRNA | 4 | 12% (3% to 21%) | 0.1372 | 0.0036 | 45.7% (0% to 81.9%) | 0.1372 | 5.52 | 3 | 0.65151 |
| mRNA-1273 | 0 | N/A | N/A | N/A | N/A | N/A | N/A | N/A | N/A |

| | | | | | | | | | |
|---|---|---|---|---|---|---|---|---|---|
| **Symptomatic Infection** | | | | | | | | | |
| The Overall Result | 12 | 22% (11% to 34%) | <0,0001 | 0.0371 | 98.5% (98.1% to 98.8%) | <0,0001 | 747 | 11 | 0.989773 |
| *Study Design Subgroup:* | | | | | | | | | |
| Cohort | 2 | 51% (49% to 53%) | 0.5383 | 0 | N/A | 0.5383 | 0.38 | 1 | N/A |
| Case-control | 10 | 17% (12% to 22%) | <0,0001 | 0.0043 | 81.3% (66.7% to 89.5%) | <0,0001 | 48.11 | 9 | 0.844111 |
| *Vaccine Subgroup:* | | | | | | | | | |
| Ad26.COV2 | 0 | N/A | N/A | N/A | N/A | N/A | N/A | N/A | N/A |
| BNT162b2 | 5 | 25% (5% to 46%) | 0.0001 | 0.0504 | 99.4% (99.1% to 99.5%) | 0.0001 | 616.01 | 4 | 0.827264 |
| ChAdOx1 nCov-19 | 1 | 16% (2% to 30%) | N/A | N/A | N/A | N/A | N/A | N/A | N/A |
| Mixture | 0 | N/A | N/A | N/A | N/A | N/A | N/A | N/A | N/A |
| Mixture of mRNA | 3 | 17% (1% to 33%) | <0.0001 | 0.0177 | 90.7% (75.6% to 96.5%) | <0.0001 | 21.51 | 2 | 0.842218 |
| mRNA-1273 | 3 | 23% (-7% to 54%) | <0,0001 | 0.0065 | 97.9% (96% to 98.9%) | <0,0001 | 94.24 | 2 | 0.928712 |

Abbreviation: k, number of analyses; N/A, not available; VED, vaccine effectiveness difference.

### S.11. Detailed Results of Meta-analyses for the 'Within 3 Months or More' Model

#### Table S16. Detailed Results of meta-analyses for within 3 months or more model

| Variable | k | VED (95%CI) | p-value of VED | Tau2 | I2 (95%CI) | p-value of I2 | Chi^2 | df | p-value of Egger's test |
|---|---|---|---|---|---|---|---|---|---|
| **Any Infection** | | | | | | | | | |
| The Overall Result | 28 | 30% (24% to 37%) | <0.0001 | 0.0362 | 97.6% (97% to 98%) | <0.0001 | 1103.33 | 27 | 0.583963 |
| *Study Design Subgroup:* | | | | | | | | | |
| Cohort | 6 | 43% (22% to 64%) | <0.0001 | 0.0661 | 99.3% (99% to 99.5%) | <0,0001 | 765.77 | 5 | 0.52177 |
| Case-control | 22 | 27% (21% to 34%) | <0.0001 | 0.019 | 93.8% (92% to 95.3%) | <0.0001 | 336.14 | 21 | 0.827997 |
| *Vaccine Subgroup:* | | | | | | | | | |
| Ad26.COV2 | 0 | N/A | N/A | N/A | N/A | N/A | N/A | N/A | N/A |
| BNT162b2 | 9 | 40% (28% to 52%) | <0.0001 | 0.0271 | 97.7% (97% to 98.3%) | <0.0001 | 345.71 | 8 | 0.666296 |
| ChAdOx1 nCov-19 | 1 | 27% (14% to 41%) | <0.0001 | N/A | N/A | N/A | N/A | N/A | N/A |
| Mixture | 2 | 29% (-4% to 62%) | 0.0863 | 0.0542 | 96.7% (91% to 98.8%) | <0.0001 | 30.12 | 1 | N/A |
| Mixture of mRNA | 10 | 21% (14% to 27%) | <0.0001 | 0.0067 | 79% (62% to 88.4%) | <0.0001 | 42.93 | 9 | 0.796208 |
| mRNA-1273 | 6 | 36% (22% to 50%) | <0.0001 | 0.0241 | 93.3% (88% to 96.2%) | <0.0001 | 74.36 | 5 | 0.291389 |
| **Severe Infection** | | | | | | | | | |
| The Overall Result | 9 | 18% (13% to 23%) | <0.0001 | 0.0043 | 88.8% (81% to 93.4%) | <0,0001 | 71.34 | 8 | 0.876591 |
| *Study Design Subgroup:* | | | | | | | | | |
| Cohort | 2 | 35% (13% to 57%) | 0.0016 | 0.0202 | 80% (14% to 95.3%) | 0.0255 | 4.99 | 1 | N/A |
| Case-control | 7 | 11% (5% to 17%) | 0.0004 | 0.0014 | 21.1% (0% to 64.4%) | 0.2688 | 7.6 | 6 | 0.305242 |
| *Vaccine Subgroup:* | | | | | | | | | |
| Ad26.COV2 | 0 | N/A | N/A | N/A | N/A | N/A | N/A | N/A | N/A |
| BNT162b2 | 2 | 22% (9% to 35%) | 0.0008 | 0 | 0% (0% to 0%) | 0.7444 | 0.11 | 1 | N/A |
| ChAdOx1 nCov-19 | 0 | N/A | N/A | N/A | N/A | N/A | N/A | N/A | N/A |
| Mixture | 1 | 46% (34% to 58%) | <0.0001 | N/A | N/A | N/A | N/A | N/A | N/A |
| Mixture of mRNA | 5 | 11% (3% to 18%) | 0.005 | 0.0028 | 41.8% (0% to 78.6%) | 0.1425 | 6.88 | 4 | 0.483804 |
| mRNA-1273 | 1 | 15% (-24% to 54%) | 0.4578 | N/A | N/A | N/A | N/A | N/A | N/A |

| | | | | | | | | | |
|---|---|---|---|---|---|---|---|---|---|
| **Symptomatic Infection** | | | | | | | | | |
| The Overall Result | 14 | 37% (29% to 46%) | <0.0001 | 0.0226 | 97.1% (96% to 97.8%) | <0.0001 | 451.64 | 13 | 0.755126 |
| *Study Design Subgroup:* | | | | | | | | | |
| Cohort | 2 | 51% (49% to 53%) | 0 | 0 | 0% (0% to 0%) | 0.5383 | 0.38 | 1 | N/A |
| Case-control | 12 | 34% (25% to 44%) | <0.0001 | 0.023 | 95.5% (94% to 96.8%) | <0,0001 | 243.29 | 11 | 0.444867 |
| *Vaccine Subgroup:* | | | | | | | | | |
| Ad26.COV2 | 0 | N/A | N/A | N/A | N/A | N/A | N/A | N/A | N/A |
| BNT162b2 | 6 | 40% (26% to 55%) | <0.0001 | 0.027 | 98.5% (98% to 98.9%) | <0.0001 | 329.82 | 5 | 0.652238 |
| ChAdOx1 nCov-19 | 1 | 27% (14% to 41%) | <0.0001 | N/A | N/A | N/A | N/A | N/A | N/A |
| Mixture | 0 | N/A | N/A | N/A | N/A | N/A | N/A | N/A | N/A |
| Mixture of mRNA | 3 | 28% (20% to 36%) | <0.0001 | 0.0031 | 61.1% (0% to 88.9%) | 0.0767 | 5.14 | 2 | 0.183352 |
| mRNA-1273 | 4 | 41% (25% to 56%) | <0.0001 | 0.0226 | 94.4% (89% to 97.2%) | <0,0001 | 53.56 | 3 | 0.470046 |

*The Egger's test has p-value <0.05 but cannot be an indication of publication bias due to k<10.
Abbreviation: k, number of analyses; N/A, not available; VE, vaccine effectiveness.

#### S.12. Sensitivity Analyses for the ‘Within 3 Months’ Model

Figure S1. The result of leave-one-out sensitivity analyses for any infection endpoint in the ‘within 3 months’ model

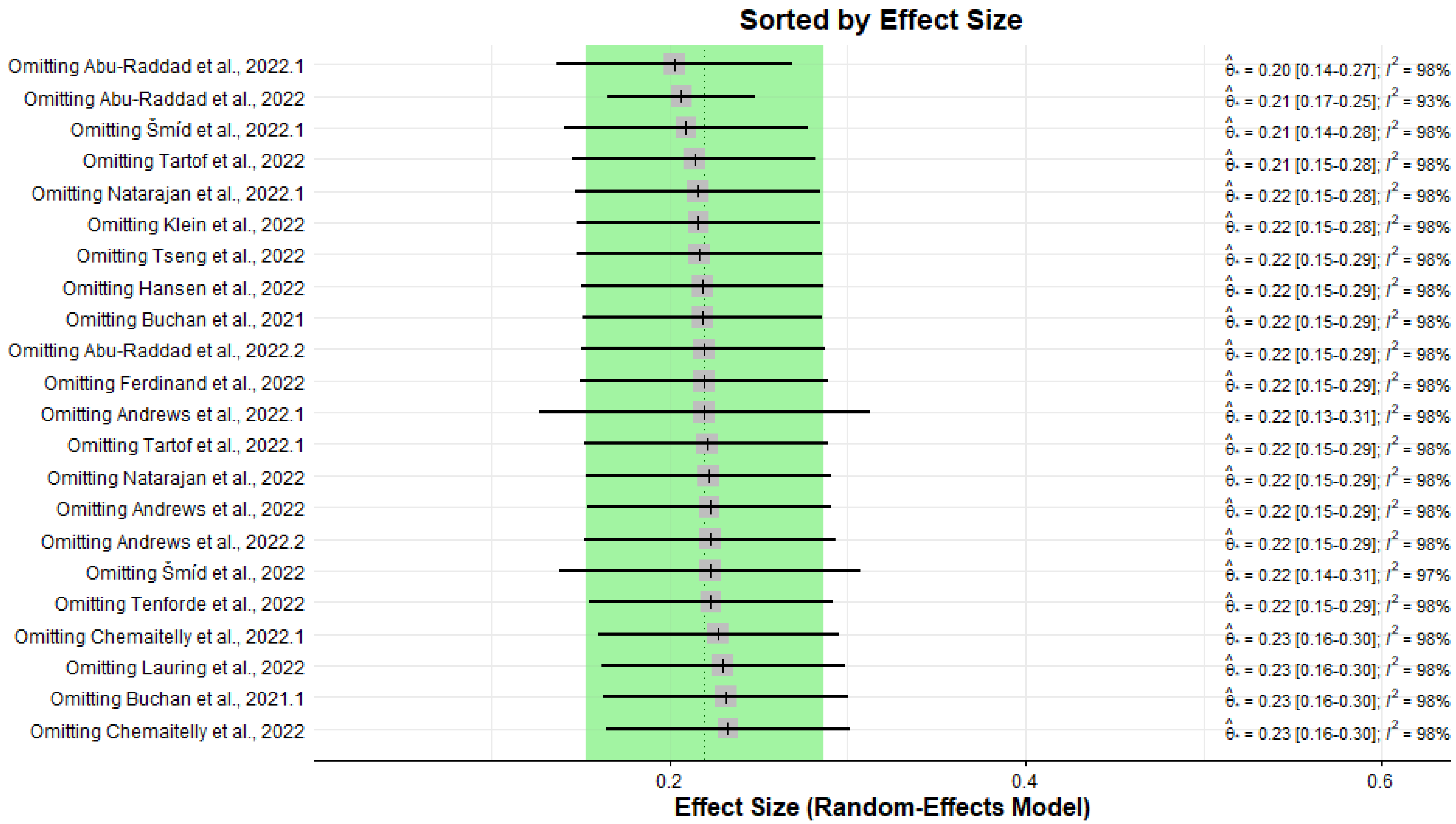

Figure S2. The result of leave-one-out sensitivity analyses for severe infection endpoint in the ‘within 3 months’ model

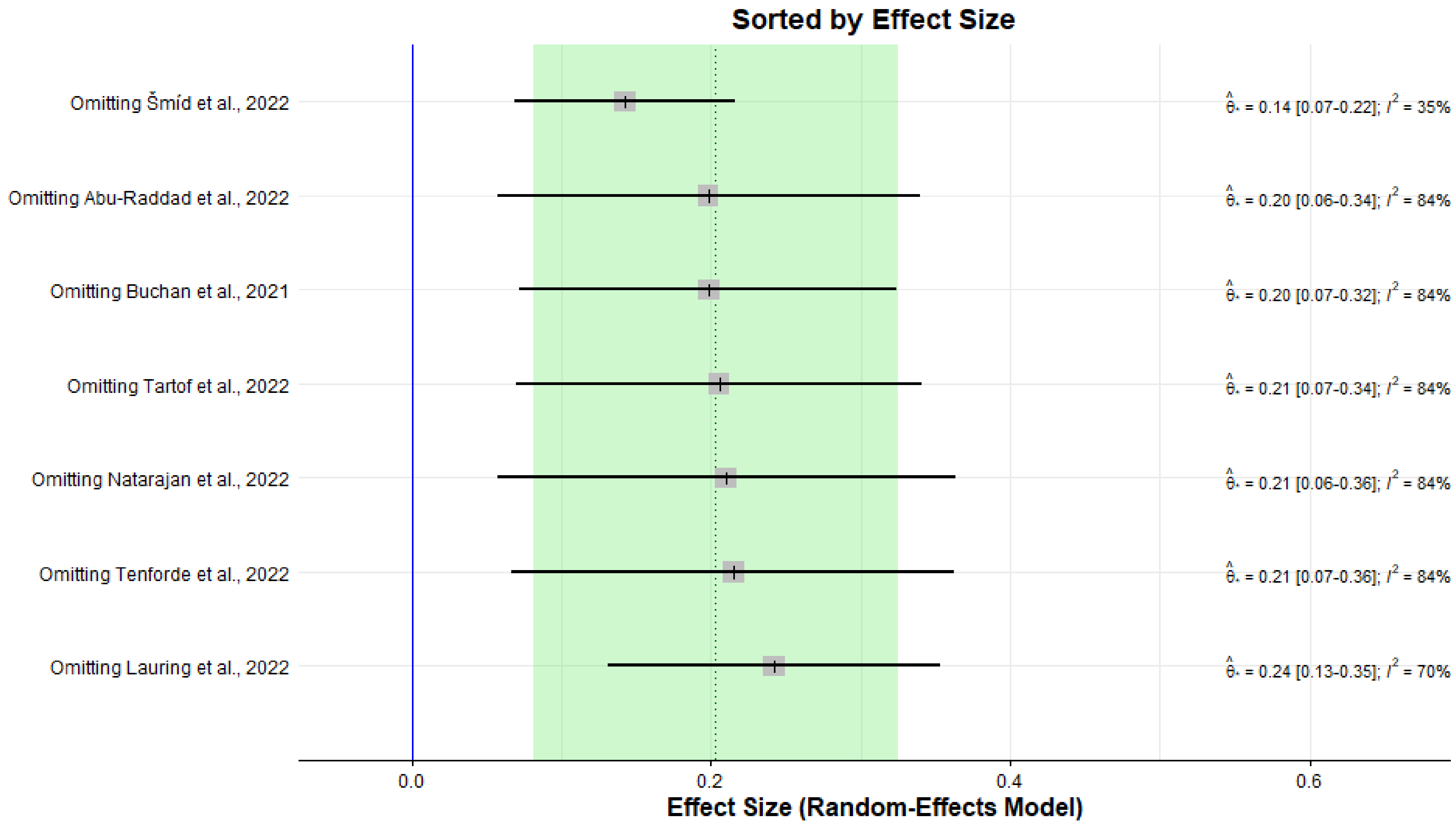

Figure S3. The result of leave-one-out sensitivity analyses for symptomatic infection endpoint in the ‘within 3 months’ model

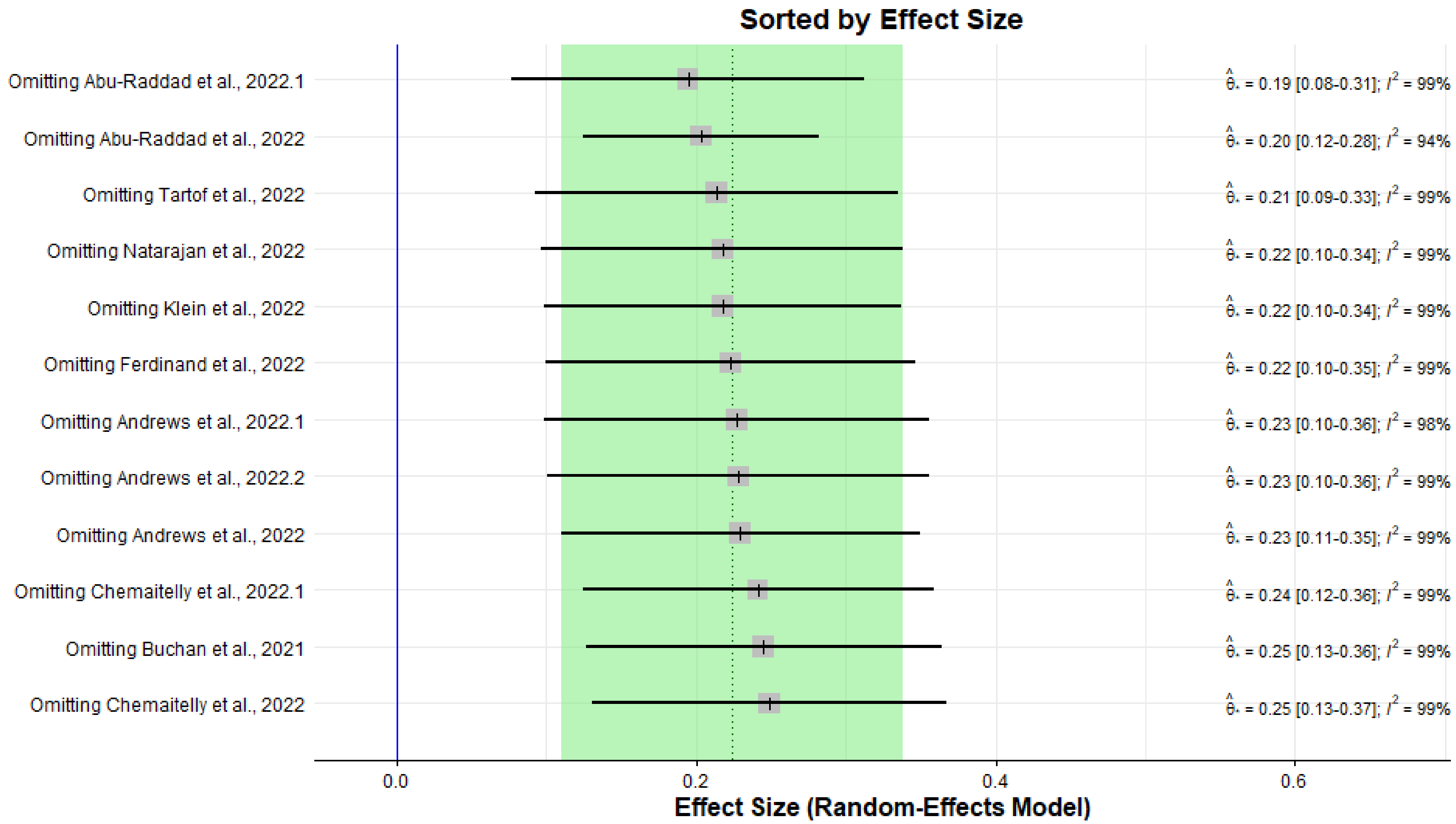

#### S.13. Sensitivity Analyses for the 'Within 3 Months or More' Model

Figure S4. The result of leave-one-out sensitivity analyses for any infection endpoint in the 'within 3 months or more' model

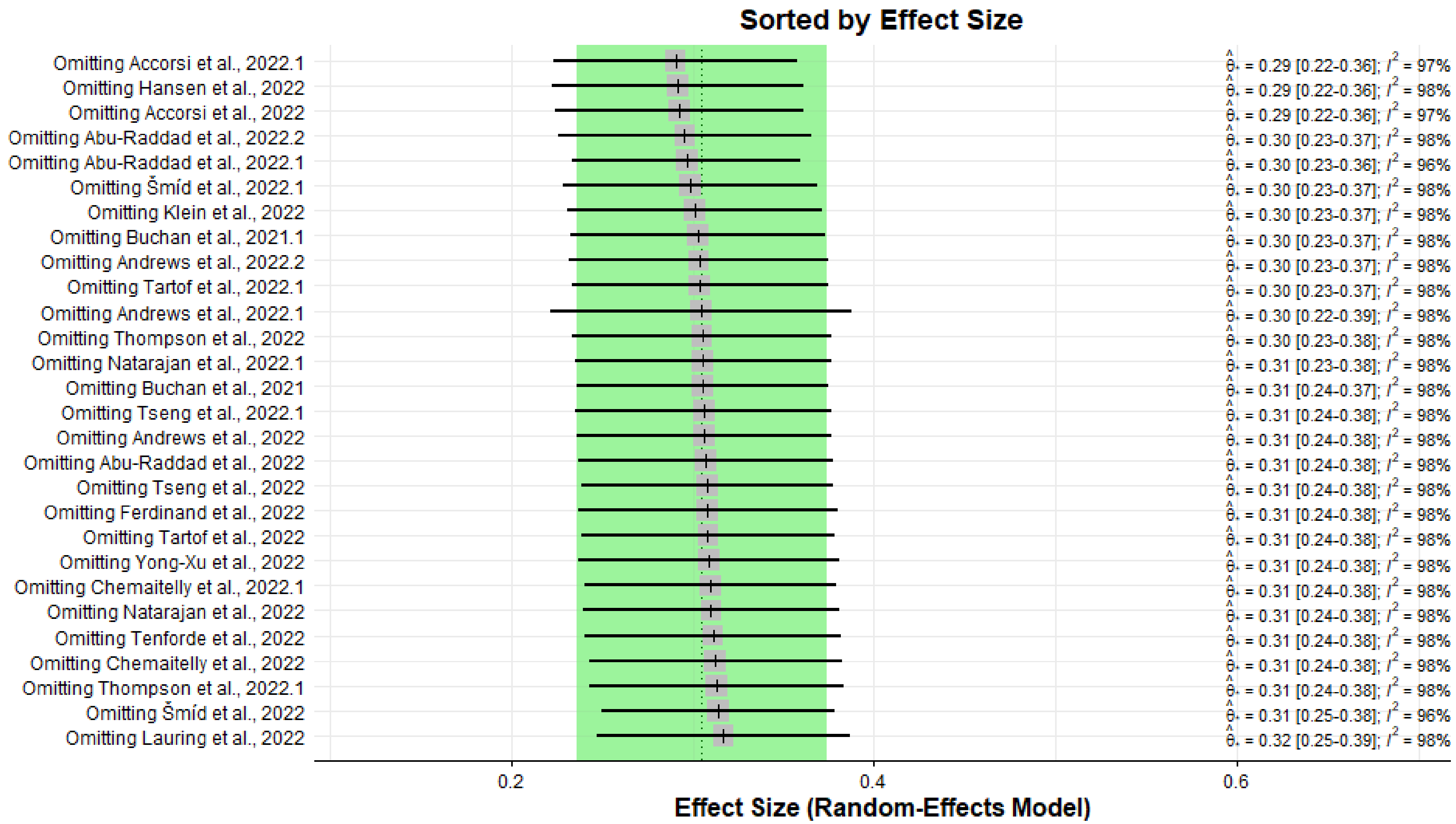

Figure S5. The result of leave-one-out sensitivity analyses for severe infection endpoint in the ‘within 3 months or more’ model

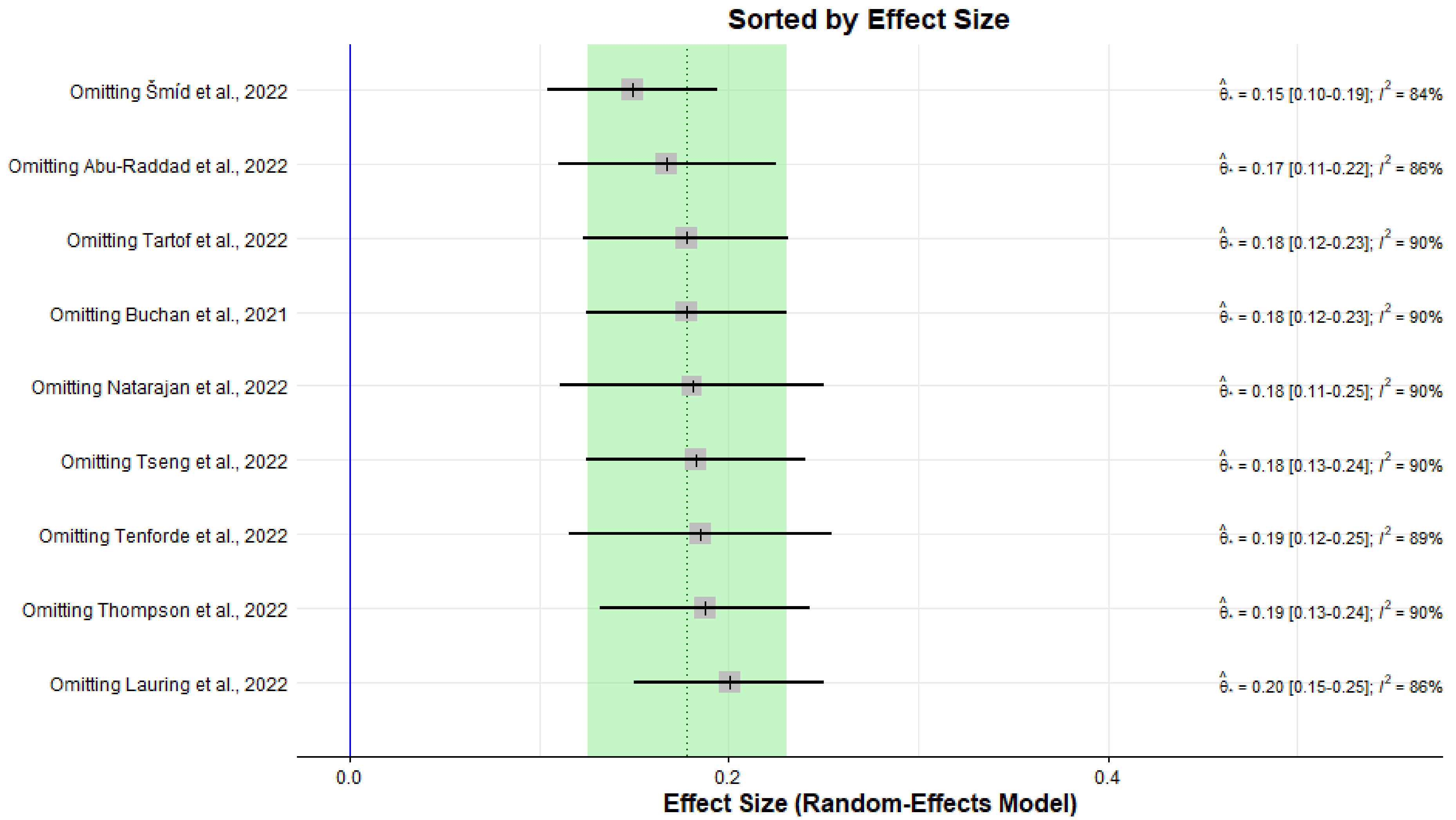

Figure S6. The result of leave-one-out sensitivity analyses for symptomatic infection endpoint in the 'within 3 months or more' model

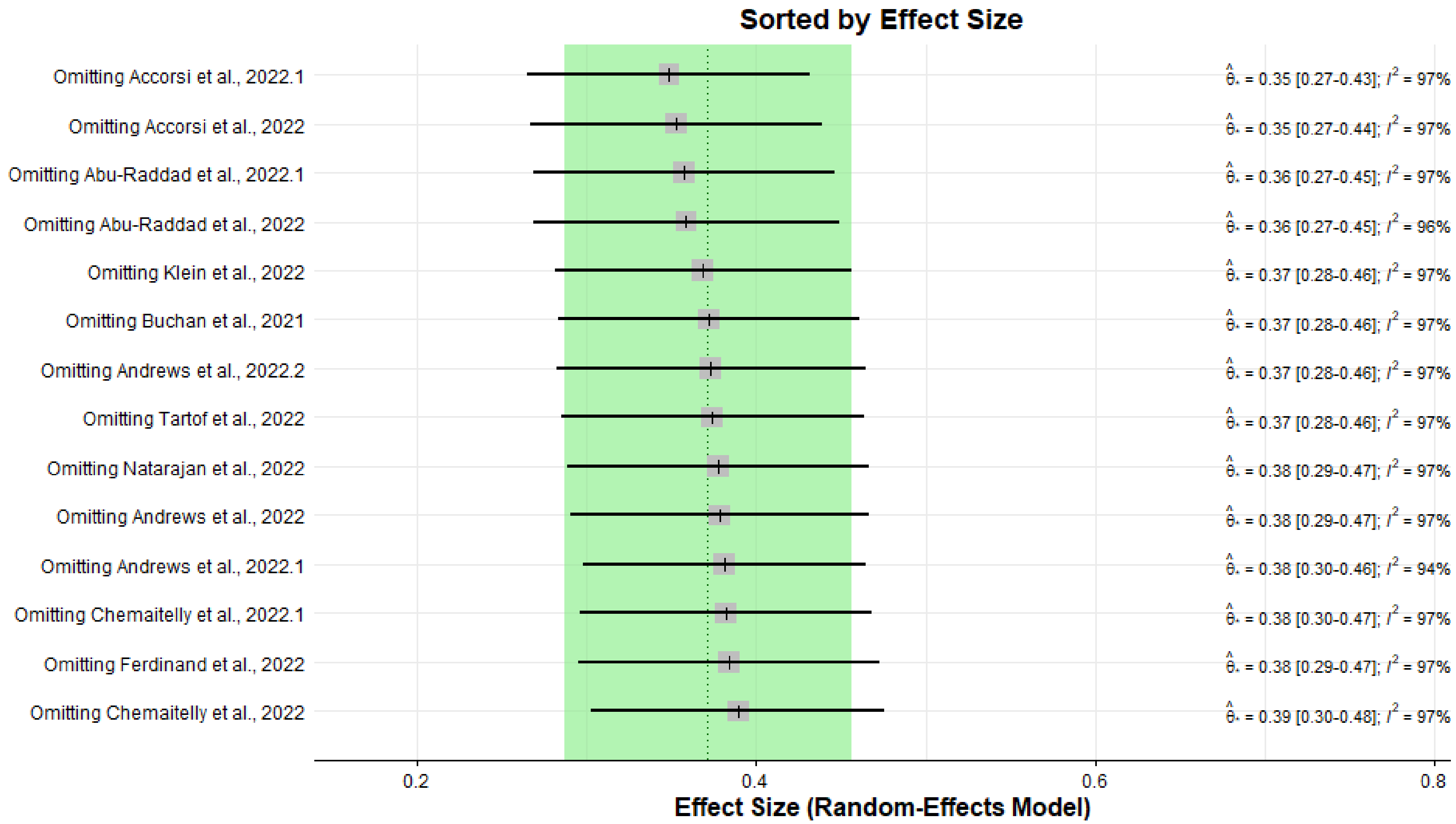
